# A Phase II Study Integrating a Single-Blind Safety Phase with a Double-Blind, Placebo-Controlled Randomized Phase, Assessing Single-Dose Intramuscular or Intranasal Administration to Evaluate the Safety and Immunogenicity of the Recombinant Vaccine Against COVID-19 (AVX/COVID-12 “Patria”) Based on an Active Newcastle Disease Viral Vector as a Heterologous Booster in Subjects with Evidence of Previous Immunity to SARS-CoV-2

**DOI:** 10.1101/2024.02.11.24302594

**Authors:** Constantino López-Macías, Martha Torres, Brenda Armenta-Copca, Niels Wacher, Laura Castro-Castrezana, Andrea Alicia Colli-Domínguez, Tania Rivera-Hernández, Alejandro Torres-Flores, Luis Ramírez-Martínez, Georgina Paz-De la Rosa, Oscar Rojas-Martínez, Alejandro Suárez-Martínez, Gustavo Peralta-Sánchez, Claudia Carranza, Esmeralda Juárez, Horacio Zamudio-Meza, Laura E. Carreto-Binaghi, Mercedes Viettri, Damaris Romero-Rodríguez, Andrea Palencia, Edgar Reyna-Rosas, José E. Márquez-García, David Sarfati-Mizrahi, Weina Sun, Héctor Elías Chagoya-Cortés, Felipa Castro-Peralta, Peter Palese, Florian Krammer, Adolfo García-Sastre, Bernardo Lozano-Dubernard

## Abstract

**Background:** The global inequity in coronavirus disease 2019 (COVID-19) vaccine distribution, primarily affecting low- and middle-income countries (LMICs), highlights the urgent need for innovative and cost-effective vaccine technologies to address availability disparities. This is crucial for achieving and sustaining widespread immunity and protecting vulnerable populations during future booster campaigns.

**Methods:** To address this need, we conducted a phase II clinical trial evaluating the safety and immunogenicity of the AVX/COVID-12 “Patria” vaccine as a booster dose. The vaccine was administered through both intramuscular (IM) and intranasal (IN) routes to participants who had previously received severe acute respiratory syndrome coronavirus 2 (SARS-CoV-2) vaccines based on adenoviral technology, inactivated virus, or mRNA technology. The inclusion criterion involved individuals with initial anti-spike IgG titers below 1,200 U/mL, allowing observation of the booster effect induced by vaccination.

**Results:** Immunization with AVX/COVID-12 resulted in a significant (>2.5 times) increase in neutralizing antibodies against the original Wuhan strain and variants of concern (VOCs) such as Alpha, Beta, Delta, and Omicron (BA.2 and BA.5). This immune response was accompanied by cellular interferon-gamma (IFN-γ) production, indicating a robust and multifaceted reaction.

**Conclusions:** The administration of AVX/COVID-12 as a booster dose, whether through IM or IN routes, was safe and well-tolerated. The vaccine extended immune responses not only against the original Wuhan-1 strain but also against various VOCs. Its ability to enhance preexisting immune responses suggests a potential contribution to expanding and sustaining herd immunity within the population.

## Introduction

Since 2020, SARS-CoV-2 and its variants have led to widespread infections and millions of deaths worldwide (1). The unequal distribution of vaccines globally, particularly impacting low- and middle-income countries (LMICs), underscores the necessity for developing new vaccine platforms to address distribution disparities (2). Ensuring a high vaccination rate globally is crucial for achieving widespread immunity. There is a strong possibility that administering periodic booster doses in the future will be necessary to prevent subsequent waves of COVID-19. Consequently, the global vaccine supply will continue to be a problem that needs to be addressed.

Population protection against COVID-19 is achieved by immunity, which has been extensively developed through a combination of infection and vaccination, including the use of vaccines that employ the spike protein from the original Wuhan-1 variant of SARS-CoV-2 as an antigen. This protection is referred to as “hybrid immunity.” Various scientific articles have demonstrated that hybrid immunity is highly effective in primarily preventing hospitalization and death caused by variants of concern, including Omicron and its predominant subvariants, even though protection against infection may have diminished (3,4). In other words, first-generation vaccines remain valuable for strengthening and expanding acquired SARS-CoV-2 immunity, leading the World Health Organization (WHO) to continue recommending their use when vaccines targeting more recent variants are unavailable (5).

Supporting this point, it is noteworthy that hospitalization and mortality have significantly decreased since the introduction of vaccination with first-generation vaccines. Some studies report a 25% reduction in the risk of hospitalization and a 96% reduction in mortality for each additional dose administered (6). In Mexico, the mortality rate before vaccination was 1.04 per 1,000 inhabitants; after the implementation of vaccination and the development of hybrid immunity, a mortality rate of 0.01 per 1,000 inhabitants has been achieved in the sixth wave, despite being caused by different VOCs, including Omicron (7).

Unfortunately, the mutation rate of SARS-CoV-2 has been until now faster than our capacity to generate updated vaccines. Nevertheless, it is desirable to develop vaccines as close as possible to the circulating strains, aiming to protect not only against hospitalization and death but also against symptomatic disease. Furthermore, there is a need for the development of vaccines that prevent transmission. One possibility is the creation of vaccines that induce mucosal immunity to avert the initial viral infection at the site of entrance, such as intranasally administered vaccines.

The AVX/COVID-12 vaccine candidate “Patria” utilizes a vector based on the Newcastle disease virus LaSota (NDV-LaSota), expressing a stabilized prefusion conformation version of the spike (S) protein of the SARS-CoV-2, known as HexaPro S (8,9). This vector presents distinct advantages, including a standardized production process in embryonated chicken eggs, a method commonly used in influenza vaccine manufacturing. This approach allows for efficient dose generation and provides the added benefit of intranasal administration, further enhancing its usability. The IN administration feature is particularly advantageous, offering an alternative and potentially more accessible method of delivery compared to traditional IM routes. This characteristic is desirable to expand vaccination coverage, especially in areas with low immunization rates or incomplete vaccination schedules.

The NDV-LaSota vector expressing HexaPro S elicited a strong immune response with IgA, IgG2a antibodies, and IFN-γ-producing T cells when used in mice and hamsters, providing complete protection against SARS-CoV-2 challenge. Trials in pigs assessed AVX/COVID-12 safety and immunogenicity via IM and IN administration, revealing no inflammation, fibrosis, or abscesses at the application site. The study demonstrated robust serum-neutralizing antibodies, cross-reactivity to spike protein epitopes in VOCs, and overall safety without signs of viral infection (10,11).

In a phase I clinical trial (NCT04871737) that included 91 volunteers, AVX/COVID-12’s safety and immunogenicity were assessed with various prime-boost regimens administered IM and IN. Three 50% embryo infective doses (EID50%) (Low: 10^7.0^ EID50%, Middle: 10^7.5^ EID50% or High: 10^8.0^ EID50%) were well-tolerated, with minimal reactogenicity. No serious adverse events were associated with any route or dose, confirming the vaccine’s safety and immunogenic profile (12). The study laid the foundation for subsequent clinical development.

In this study, we assessed the safety and immunogenicity of the AVX/COVID-12 vaccine in a phase II clinical trial. Administered as a booster dose (both IM and IN), AVX/COVID-12 was given to participants who had previously received SARS-CoV-2 vaccines based on adenoviral technology, inactivated virus, or mRNA technology, primarily with initial anti-S IgG titers below 1,200 U/mL. These titers were considered low enough to observe the booster effect induced by the IM and IN administration of AVX/COVID-12.

## Material and methods

### Study design

A phase II study (ClinicalTrials.gov #NCT05205746) was conducted to assess the safety phase under single-blind conditions, followed by randomized, double-blind, placebo-controlled, single-dose administration via IM or IN routes, in subjects with evidence of prior immunity to SARS-CoV-2. This was followed by an assessment phase to evaluate the booster response, involving an IM dose of the COVID-19 vaccine (recombinant AZ/ChAdOx-1-S) in subjects originally randomized to the placebo arm.

The protocol was formulated by iLS Clinical Research, S.C. in collaboration with the Mexican Institute for Social Security (IMSS) and Laboratorio Avi-Mex, S.A. de C.V. (Avimex®), which was the sponsor. Approval for the study was granted by the Federal Commission for the Protection against Sanitary Risks (COFEPRIS) in Mexico, assigned the number RNEC2021-AVXSARSCoV2VAC002. Ethics approval was obtained from the institutional ethics committee at each research site participating in the study. The research was conducted in complete compliance with Mexican regulations and by the principles outlined in the Declaration of Helsinki and Good Clinical Practice. Immunological assay samples were processed at the National Institute for Respiratory Diseases (INER) in Mexico City.

### Study groups

The study groups included adults aged 18 years or older of both genders who signed informed consent and had not experienced respiratory issues in the 21 days preceding the administration of the single dose. Participants committed to maintaining appropriate preventive measures to avoid SARS-CoV-2 infection throughout their study participation, particularly during the initial 14 days following the baseline visit. Additionally, eligibility criteria involved having detectable anti-spike IgG titers in serum during the screening visit, with titers levels less than 1,200 U/mL measured in a chemiluminescence test. Furthermore, participants were required to provide evidence of vaccination, with a minimum of 4 months having passed since the last vaccination. Due to the nature of the study, recruited subjects were enrolled in one of three subgroups based on the vaccine technology used during the primary immunization against SARS-CoV-2: subjects with a history of prior immunization with vaccines based on adenoviral technology, inactivated virus, or mRNA technology.

The exclusion criteria encompassed hypersensitivity or allergy to any component of the vaccine, a history of severe anaphylactic reactions, a history of seizures, uncontrolled chronic diseases, chronic conditions requiring treatment with immunosuppressive agents or immune response modulators (e.g., systemic corticosteroids, cyclosporine, rituximab, among others), the development of oncological diseases, active participation in or involvement within the last three months in any other clinical or experimental intervention study, use of any drug or herbal supplement in the 30 days preceding the screening evaluation, or alternative medicine (e.g., chlorine dioxide, etc.) aimed at treating or preventing complications or SARS-CoV-2 infection, or any other condition, fever at the time of the screening visit, and having received any vaccine (experimental or approved) in the 60 days prior to the screening visit, except for the influenza vaccine. Exclusion criteria also included having received a blood or blood component transfusion, or being a plasma donor within the 4 months prior to the screening visit, undergoing dialysis or hemodialysis procedures in the last year before the screening visit, working on poultry or gamecock farms, and having a history of substance abuse problems that, in the investigator’s judgment, could interfere with the subject’s ability to comply adequately with the protocol guidelines.

The randomization of volunteers was conducted using a computer-based random assignment system, ensuring blinding throughout the study. Upon signing the informed consent form, each participant received a patient number, encoding all their information in a pseudo-anonymized manner during data collection and becoming fully anonymized during the analysis.

The stratification for subject randomization was generated as follows:

Safety phase: Involved single-blinded assignment (blinded to the research subject) to the arm receiving the AVX/COVID-12 vaccine for the first three subjects of each specific vaccine recruited in each branch, IM and IN. For this purpose, eight blocks of six vaccines were generated, and to each block, three subjects with the AVX/COVID-12 vaccine and three subjects with placebo were randomly assigned.

Double-blind, placebo-controlled randomized phase: subjects with the corresponding vaccination history were randomized to receive a placebo or the AVX/COVID-12 vaccine in either of the two administration modalities (IM or IN). Starting from block 9, blocks of 4 were generated, and each block was randomly assigned 1 subject with IM AVX/COVID-12 vaccine, 1 subject with IN AVX/COVID-12 vaccine, 1 subject with IM placebo, and 1 subject with IN placebo.

After 14 days, following the administration of the AVX/COVID-12 vaccine or placebo, the blinding was revealed for subjects who received a placebo. These subjects received an additional dose of the COVID-19 vaccine (AZ/ChAdOx-1-S) IM.

### AVX/COVID-12 vaccine preparation

The AVX/COVID-12 vaccine was manufactured in a purpose-built good manufacturing practice (GMP) facility. The recombinant virus was cultivated in 10-day-old specific-pathogen-free (SPF) chicken embryos through inoculation via the allantoic cavity with 10^3.3^ EID50/0.1 mL of the production seed. The embryos were incubated for a period of 72 hours (h) at an average temperature of 37 °C and a relative humidity range of 60% to 70%. After this period, the embryos were refrigerated for at least 12 h to sacrifice them. Harvesting of the allantoic fluid (AF) was conducted under aseptic conditions using a vacuum. The AF was clarified through filters with a pore size of 0.8 to 8.0 μm, concentrated by a factor of 10X in 300-kDa cassettes, and subjected to diafiltration in 20 volumes of phosphate-buffered saline (PBS). The AF was stored frozen at −70 °C. The AF yielded a titer of 10^8.67^ EID50/mL, from which the frozen active vaccines were prepared. The corresponding volumes of purified AF were aseptically mixed with the stabilizer trehalose, monobasic potassium phosphate, dibasic sodium phosphate and monosodium glutamate and homogeneously mixed for 5 minutes using a Teflon bar and a magnetic stirrer. Finally, they were packed in high-density polyethylene bottles. The vaccines were stored at −80 °C.

### Safety

Safety was assessed by comparing adverse events (AEs) between placebo and AVX/COVID-12 vaccine groups, considering their severity and relationship to the intervention. While all participants completed the same visits, those assigned to specific groups underwent additional sample collections during visits corresponding to days 0, 14, 42, 90, 180 and 365 of the study. The primary endpoints in the phase II aspects of this trial included solicited local and systemic AEs. The classification of potential AEs and their association with vaccine exposure followed a two-period analysis. The first period included the initial seven days following vaccination, during which AEs were deemed related to the vaccine components. Conversely, reactions occurring after seven days, typically in response to the development of an immune response, constituted the second period of analysis (13).

### Sample collection

Venous blood samples were obtained from participants during scheduled site visits for serology tests and cellular response assessments. Using standard phlebotomy procedures, the samples were collected into two sodium heparin tubes and one separator tube (SST BD vacutainer tubes, Franklin Lakes, NJ, USA). Following collection, the samples were transported at room temperature to the INER for immediate processing. All blood samples and products were handled in a BSL-2 laboratory, using appropriate personal protective equipment and safety precautions, adhering to processing protocols approved by the Institutional Biosafety Committee. Peripheral blood samples from each volunteer were centrifuged at 2,260 x *g* at room-temperature for 10 minutes and transported to a biological safety cabinet. Then, for each sample, the serum was recovered, divided into six aliquots of 200 μL, stored in pre-labeled 1.5 mL cryovials according to the established procedure, and frozen at -20 °C until use. One aliquot of serum was thawed for each antibody determination, and the remaining portion of each thawed aliquot was discarded as infectious biological material (IBM).

### Peripheral blood mononuclear cell (PBMC) isolation

PBMCs were isolated from venous blood collected in sodium heparin tubes (BD vacutainer tubes, Franklin Lakes, NJ, USA). Within four hours of collection, PBMC isolation was conducted by density-gradient sedimentation of whole blood diluted at a 1:2 ratio in Roswell Park Memorial Institute medium (RPMI) 1640 (Lonza, Basel, Switzerland) at room-temperature. The diluted blood was layered over an appropriate volume of Lymphoprep (Axis-Shield, Dundee, UK) at room-temperature. Then, the PBMC were then recovered, cryopreserved in a medium consisting of 10% dimethyl sulfoxide (DMSO; Sigma Aldrich, St. Louis, MO, USA) and 90% heat-inactivated fetal bovine serum (FBS; GIBCO, California, USA), and stored at -80 °C until use. For T-cell assays, the thawing process involved warming frozen cryovials at 37 °C in a water bath, and cells were resuspended in 5 mL of CTS OpTmizer (GIBCO, California, USA). After a wash with OpTmizer, cells were counted using a Neubauer chamber, and the viability was assessed by trypan blue; cells were resuspended in OpTmizer medium to a density of 4 x 10^6^ cells/mL.

The subunit 1 of the spike protein of SARS-CoV-2 (RayBiotech, Peachtree Corners, Georgia, USA) was selected as the primary antigen due to its display of the most immunogenic epitopes of the spike protein. Additionally, it contains the receptor binding domain (RBD) fragment of the virus, which mediates binding to the host receptor angiotensin-converting enzyme 2 (ACE2) in lung cells.

### SARS-CoV-2 detection

Sample processing was conducted through nucleic acid extraction, followed by one-step real-time RT-PCR amplification and detection for SARS-CoV-2. A Roche Magnapure instrument (Roche Diagnostics, Basel, Switzerland) was used for viral RNA extraction, and the following ModularDx kits (TIB Molbiol, Berlin, Germany) were employed:

- 53-0775-96 Sarbeco N-gene detects SARS and SARS-CoV-2
- 53-0776-96 Sarbeco E-gene detects SARS and SARS-CoV-2
- 53-0777-96 SARS-CoV-2 RdRP detects SARS-CoV-2

### Determination of IgG anti-SARS-CoV-2 binding antibodies

For the determination of total anti-SARS-CoV-2 serum antibodies, a semi-quantitative enzyme-linked immunosorbent assay (ELISA) was employed to detect IgG antibodies to the spike protein subunit 1 domain (ELISA anti-SARS-CoV-2 (IgG); Catalog # 2606-9601 G; EUROIMMUN, Lübeck, Germany). Samples were processed according to the manufacturer’s instructions and analyzed on the EUROIMMUN Analyzer I (EUROIMMUN, Lübeck, Germany). Results are reported semi-quantitatively in ELISA ratio, representing a measure of the relative antibody concentration in participants’ serum. For the calculation of the relative concentration, the ratio of the extinction value of the control or sample to the extinction value of the calibrator was determined using the following formula: relative concentration (ratio) = (extinction of control or volunteer sample) / (extinction of calibrator).

Interpretation of results, in terms of positivity, was conducted according to the manufacturer’s instructions:

- Ratio <0.80: Negative
- Ratio >0.8 to <1.1: Uncertain
- Ratio >1.1: Positive

To estimate the geometric mean titers (GMT) of the IgG anti-spike subunit 1 antibodies in participants’ serum, serial dilutions (1:100, 1:300, 1:900, 1:2700, 1:5400) were performed for all serum samples with ratio values equal to or greater than 1.1 at the 1:100 dilution. For sera with ratio values < 0.8 at the 1:100 dilution, a titer of 1:20 was assigned for simplified analysis.

### Pseudovirus neutralization assay

The presence of neutralizing antibodies was assessed using a microneutralization assay. The assay relies on the inhibition capacity of infection in an *in vitro* model present in serum samples and employed recombinant vesicular stomatitis virus (rVSV) pseudovirus particles expressing the protein from the ancestral Wuhan-1 strain, Alpha, Delta, Beta, and the Omicron subvariants BA.2 (clone 1) and BA.5 (clone 4) (VSV-eGFP-SARS2) kindly provided by Dr. Sean Whelan (Washington University, USA). The cellular model used to assess the inhibition capacity was the Vero E6 cell line (The American Type Culture Collection (ATCC) CRL-1586, Manassas, VA, USA), which is susceptible to infection as it expresses the recognition receptors for the SARS-CoV-2 spike protein.

Before performing the assay (previously validated and established as an analytical method), all serum samples were inactivated at 56 °C for 30 minutes. In 96-well plates, 1.3 x 10^4^ Vero E6 cells (ATCC, Manassas, VA, USA) per well were seeded and incubated for 24 hours at 37 °C, 5% CO2, allowing them to reach 85% confluence. Next, serum samples were serially diluted 1:2 in FBS-free Eagle’s minimal essential medium (EMEM; Rahway, New Jersey, USA) in duplicate, starting with a 1:20 dilution up to 1:1280. Diluted samples were mixed with the concentration of particles of VSV-SARS-CoV-2 S pseudoviruses from the ancestral strain and the aforementioned variants, capable of causing cellular damage at 50% (50% tissue culture infectious dose, TCID50). The mixture was incubated for 60 minutes at 37 °C, 5% CO2. Then, each serum dilution was transferred to a parallel plate containing Vero E6 cells and incubated for 72 h at 37 °C, 5% CO2.

After the incubation period, 3.7% formaldehyde was added and incubated for 30-40 minutes at room temperature. The plates were washed twice with PBS, and the recording and analysis of the plates was carried out by optical microscopy, documenting the presence or absence of damage to the monolayer, known as cytopathic effect, at each dilution of the different serum samples. These values were used to calculate the 50% protection level by applying the Spearman-Karber Method, obtaining the 50% inhibitory dilution (ID50) and the protective titer of each sample. The upper and lower detection limits for neutralizing activity are ID50 > 1.6 and a titer > 1:40 (initial dilution). Each assay included negative and positive infection controls (uninfected cells and infection control inoculated with the reporter vector VSV-eGFP-SARS2 and the subvariants in a suspension containing 100 TCID50). Titers < 1:40 (initial dilution) are reported with a value of 20 for statistical analysis.

### Evaluation of cytokines in cell culture supernatants

PBMCs were resuspended at a concentration of 2 x 10^6^ cells/mL in CTS OpTmizer medium (GIBCO, California, USA). Duplicate samples of 2 x 10^5^ cells/well were plated in 96-well plates and stimulated with 5 µg/mL of the spike protein subunit 1 (RayBiotech, Peachtree Corners, GA), using culture medium as a negative control. After 18 hours of culture, the supernatant was collected, aliquoted, and stored at -20 °C until use for cytokine determination. Subsequently, the supernatants were thawed and centrifuged at 14,000 rpm for 10 minutes, and cytokine concentrations were measured using the Th1/Th2 Bio-Plex kit (BIO-RAD, California, USA). This kit allows the quantification using the Bio-Plex 200 system and Bio-Plex Manager 6.1 software (BIO-RAD, California, USA). Values were extrapolated from a concentration curve and expressed in pg/mL.

### Hemagglutination inhibition antibody assay for Newcastle disease virus (NDV)

A hemagglutination inhibition assay was performed to assess the presence of inhibitory antibodies against specific hemagglutination of NDV. This method, adapted from the original avian sera procedure, was implemented and validated in our group for human serum analysis. The procedure utilized a system with 1% avian erythrocytes and eight hemagglutinant units (8 HAU/mL) of inactivated NDV antigen (AgNDV, DCV Inc. Lot: 23920103). Briefly, serum samples were inactivated for 30 minutes at 56 °C; 100 µL of each sample underwent pre-absorption with an equal volume of a 5% avian erythrocyte suspension and incubated overnight (16-18 h) at 2-8 °C. Next, the samples were centrifuged at 1,500 x g for 60 seconds, and the supernatants were collected and stored at -20 °C until use.

In 96-well round-bottom plates, 25 µL/well of pre-absorbed serum samples were placed in duplicate, and serial 1:2 dilutions were made (starting with a 1:8 dilution, subsequent dilutions included 1:16, 1:32, 1:64, 1:128, 1:256, 1:512, and 1:1,024). Subsequently, 25 µL/well of AgNDV suspension adjusted to 8 HAU/mL was added, and the samples were incubated (inhibition process) for 30 minutes at room-temperature (19-25 °C). Then, 25 µL of 1% avian erythrocyte suspension was added and incubated for 40 minutes at room temperature (19-25 °C). The interpretation of the plates was performed by placing them at a 45° position to observe the precipitated erythrocytes flowing (teardrop formation). The technique’s limit of detection was a titer of 1:4, and the positive cutoff limit started from the titer of 1:8. Samples that did not show inhibitory activity or whose activity was below the 1:4 dilution were assigned the value of the 1:2 dilution, corresponding to the immediately preceding dilution (15,16).

### Statistical analysis

To outline the demographic characteristics of the study population, measures of central tendency and dispersion were utilized for continuous variables, while proportions were employed for discrete or categorical variables. Continuous variables underwent analysis using one-way ANOVA and students’ t-tests, whereas non-parametric tests were applied to discrete variables.

For safety assessments, the z-test was employed to compare proportions between treatment groups in the context of vaccine versus placebo, and the chi-square test was used for comparisons involving administration routes, vaccine, or placebo.

Individual antibody titers were quantified as the highest dilution titer that elicited a positive signal. Concurrently, the T-cell response was assessed by determining the concentration of cytokine-producing cells after stimulation with the spike subunit 1 protein following an 18-hour incubation period. The population results were characterized by a geometric mean (GM) and a 95% confidence interval.

## Results

From November 23, 2022, to July 24, 2023, 1,221 volunteers underwent eligibility assessments, and the distribution of recruited volunteers is depicted in Figure 1. Various research sites across different states of Mexico participated in volunteer recruitment, including Mexico City (Investigación en Salud (CAIMED), S. A. de C. V., Unidad de Medicina Familiar No. 20 IMSS, CEMDEC S.A de C.V., and Unidad Clínica Farmacológica Bioemagno S.A. de C.V.) and Oaxaca (Oaxaca Site Management Organization (OSMO) S.C.).

**Figure 1.**
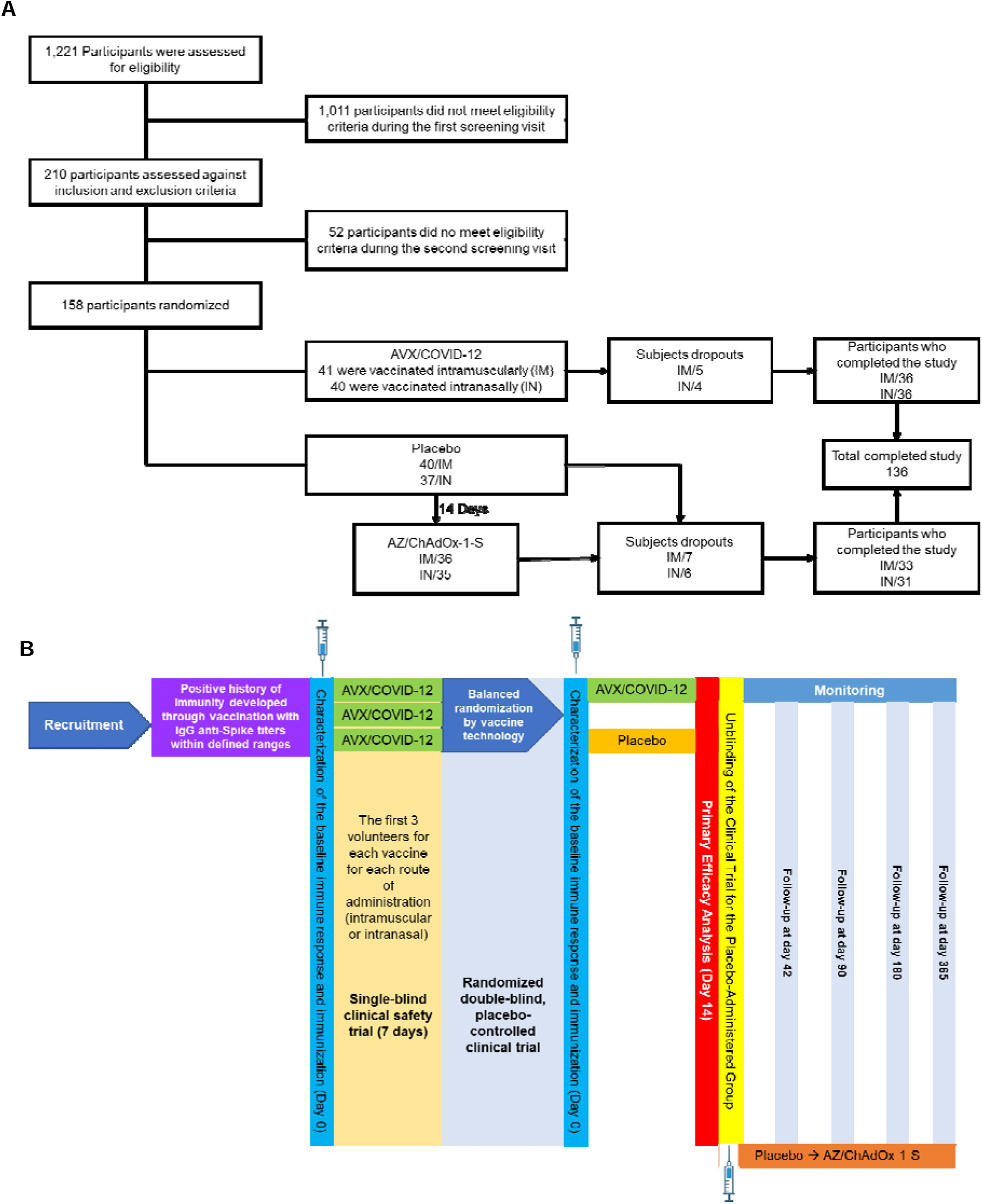
Screening and randomization of the participants. A) Flowchart depicting participant screening, randomization, and distribution across various experimental groups throughout the study. B) Scheme illustrating the distribution of volunteers into groups and post-boosting monitoring.

Of the 1,221 volunteers initially screened, 1,011 participants did not meet the eligibility criteria during their first visit. The remaining 210 participants underwent assessment against both inclusion and exclusion criteria. During the second visit, 52 participants were excluded for not meeting the eligibility and inclusion criteria as described earlier. Therefore, the final cohort of participants comprised 158 individuals who were then randomized to receive either the AVX/COVID-12 vaccine or a placebo. The administration routes included IM and IN (Figure 1A).

In the AVX/COVID-12 vaccine group, 41 participants received IM immunization, while 40 received IN. Five participants from the IM group and four from the IN group withdrew from the study, resulting in 36 participants in each analyzed subgroup. In the placebo group, 40 participants were assigned to IM and 37 to IN administration. Subsequently, participants from the placebo group (36 from the IM group and 35 from the IN group) were inoculated with the AZ/ChAdOx-1-S vaccine 14 days after initially receiving the placebo; 7 participants from the IM group and six from the IN group withdrew from the study. A total of 136 participants completed the study (Figure 1B).

There were no statistically significant differences in age, gender, body mass index, or comorbidities between the IM and IN vaccinated groups and their respective placebo groups, except for race between the AVX/COVID-12 IM vaccinated group and its placebo (p=0.012) (Table 1).

**Table 1.** Demographic characteristics of phase II study volunteers.

|  | IM AVX | IM PL | p-Value | IN AVX | IN PL | p-Value |
| --- | --- | --- | --- | --- | --- | --- |
| **Volunteers (n)** | 41 | 40 |  | 40 | 37 |  |
| **Age, mean (SD)** | 39.8 (11.9) | 33.5 (11.3) | 0.01 | 37.8 (13.5) | 36.6 (13.8) | 0.65 |
| **Age groups (years old)** |  |  |  |  |  |  |
| < 20 | - | - | 0.15 | 1 (2.5) | 1 (2.7) | 0.79 |
| 20 - 29 | 12 (29.27) | 22 (55) |  | 11 (27.5) | 14 (37.84) |  |
| 30 - 39 | 7 (17.07) | 6 (15) |  | 12 (30) | 8 (21.62) |  |
| 40 - 49 | 16 (39.02) | 7 (17.5) |  | 6 (15) | 7 (18.92) |  |
| 50 - 59 | 3 (7.32) | 3 (7.5) |  | 7 (17.5) | 4 (10.81) |  |
| 60 - 69 | 3 (7.32) | 2 (5) |  | 3 (7.5) | 2 (5.41) |  |
| 70 - 79 | - | - |  | - | 1 (2.7) |  |
| **Gender, n (%)** |  |  |  |  |  |  |
| Female | 21 (51.22) | 23 (57.5) | 0.57 | 19 (47.5) | 16 (43.24) | 0.70 |
| Male | 20 (48.78) | 17 (42.5) |  | 21 (52.5) | 21 (56.76) |  |
| **MBI, mean (SD)** | 26.3 (6.0) | 26.5 (5.1) | 0.43 | 26.7 (6.1) | 26.5 | 0.88 |
| **BMI classification** |  |  |  |  |  |  |
| Underweight | 2 (4.88) | 1 (2.5) | 0.68 | 3 (7.5) | 2 (5.41) | 0.64 |
| Normal | 11 (26.83) | 15 (37.5) |  | 14 (35) | 14 (37.84) |  |
| Overweight | 18 (43.9) | 17 (42.5) |  | 10 (25) | 13 (35.14) |  |
| Obesity | 10 (24.39) | 7 (17.5) |  | 13 (32.5) | 8 (21.62) |  |
| **Race, n (%)** |  |  |  |  |  |  |
| White | 3 (7.32) | 2 (5) | 0.012 | 1 (2.5) | 2 (5.41) | 0.14 |
| Amerindian | 10 (24.39) | 2 (5) |  | 10 (25) | 3 (8.11) |  |
| Mexican Mestizo | 27 (65.85) | 28 (70) |  | 26 (65) | 31 (83.78) |  |
| Other | 1 (2.44) | 8 (20) |  | 3 (7.5) | 1 (2.7) |  |
| **Comorbidities, n (%)** |  |  |  |  |  |  |
| 0 | 38 (92.68) | 39 (97.5) | 0.31 | 37 (92.5) | 33 (89) | 0.59 |
| 1 | 3 (7.32) | 1 (2.5) | 0.34 | 3 (7.5) | 4 (10) | 0.69 |
| **Systolic arterial hypertension** | 2 (4.87) | 1 (2.5) | 0.57 | 2 (5) | 2 (5) | 1 |
| **Type 2 Diabetes Mellitus** | - | - |  | 1 (2.5) | - | NA |
| **HIV** | 1 (2.44) | - | NA | - | 2 (5) | NA |

AVX: AVX/COVID-12; PL: placebo; IM: intramuscular; IN: intranasal; SD: Standard Deviation; BMI: Body Mass Index. Mann–Whitney's U test was conducted for median comparisons, and the Chi-Square test was used for categorical variables.

In Mexico, various vaccines, including adenoviral vectors (AstraZeneca, CanSino, Johnson & Johnson, Sputnik V), inactivated viruses (Sinovac), and mRNA (Pfizer-BioNTech), were employed for the national vaccination campaign. Therefore, an additional inclusion criterion during the recruitment phase was completing the primary vaccination series with any of the mentioned vaccines. A comparable number of individuals with a similar history of prior vaccination were enrolled in each group. No statistically significant differences were observed between the groups regarding the history of previous SARS-CoV-2 vaccination, indicating a comparable proportion of subjects vaccinated with various vaccine technologies (Table 2).

**Table 2.** Prior history of SARS-CoV-2 vaccination among phase II participants at baseline.

|  | IM AVX | IM PL | p-Value | IN AVX | IN PL | p-Value |
| --- | --- | --- | --- | --- | --- | --- |
| **Volunteers (n)** | 41 | 40 |  | 40 | 37 |  |
| **Last vaccination schedule prior to the study, n (%)** |  |  |  |  |  |  |
| **Adenoviral Vector** | 31 (75.6) | 33 (82.5) | 0.44 | 31 (77.5) | 33 (89.2) | 0.17 |
| AstraZeneca | 16 (39.02) | 16 (40) | 0.92 | 12 (30) | 14 (37.84) | 0.46 |
| CanSino | 4 (9.76) | 4 (10) | 0.97 | 5 (12.5) | 8 (21.62) | 0.28 |
| Johnson & Johnson | 1 (2.44) | - | N/A | 1 (2.5) | - | N/A |
| Sputnik V | 10 (24.39) | 13 (32.5) | 0.41 | 13 (32.5) | 11 (29.73) | 0.79 |
| **Inactivated Viruses** |  |  |  |  |  |  |
| Sinovac | 6 (14.63) | 4 (10) | 0.52 | 5 (12.5) | 3 (8.11) | 0.52 |
| **mRNA** |  |  |  |  |  |  |
| Pfizer | 4 (9.76) | 3 (7.5) | 0.71 | 4 (10) | 1 (2.7) | 0.19 |

AVX: AVX/COVID-12; PL: placebo; IM: intramuscular; IN: intranasal; SD: Standard Deviation; BMI: Body Mass Index. Mann–Whitney’s U test was used for median comparisons, and the Chi-Square test was used for categorical variables.

### Safety after vaccination with AVX/COVID-12

AEs were documented in subjects receiving IM AVX/COVID-12 (n=41), IM placebo (n=40), IN AVX/COVID-12 (n=40), and IN placebo (n=37) (total subjects 158). At day 28, IM and IN placebo group subjects received the AZ/ChAdOx-1-S vaccine, and AEs were recorded (n=71). Therefore, AEs were documented for 158 subjects receiving IM or IN AVX/COVID-12 vaccine and placebo groups, in addition to 71 subjects from the IM and IN placebo group who received the AZ/ChAdOx-1-S vaccine, making a total of 229 subjects (Tables 1S).

From the 229 subjects analyzed, 186 subjects reported AEs, with 38 belonging to the group that received the AVX/COVID-12 vaccine IM and 31 from the IM placebo group. Furthermore, 36 individuals came from the group that received the AVX/COVID-12 vaccine IN, and 29 subjects received the IN placebo. Considering the number of subjects recruited for each group that received the vaccine IM administration (vaccine or placebo), it was observed that 92.68% of participants who received the AVX/COVID-12 vaccine reported AEs, while the percentage for the IM placebo group was 77.5%. This difference in proportions for both groups was statistically significant, with a higher proportion of affected subjects in the AVX/COVID-12 vaccine group (Figure 2A). Analyzing the groups that received the AVX/COVID-12 vaccine and placebo IN, it was observed that 90% and 78.37% of subjects in these groups reported adverse events, respectively. The difference between these proportions was not statistically significant (Figure 2A).

**Figure 2.**
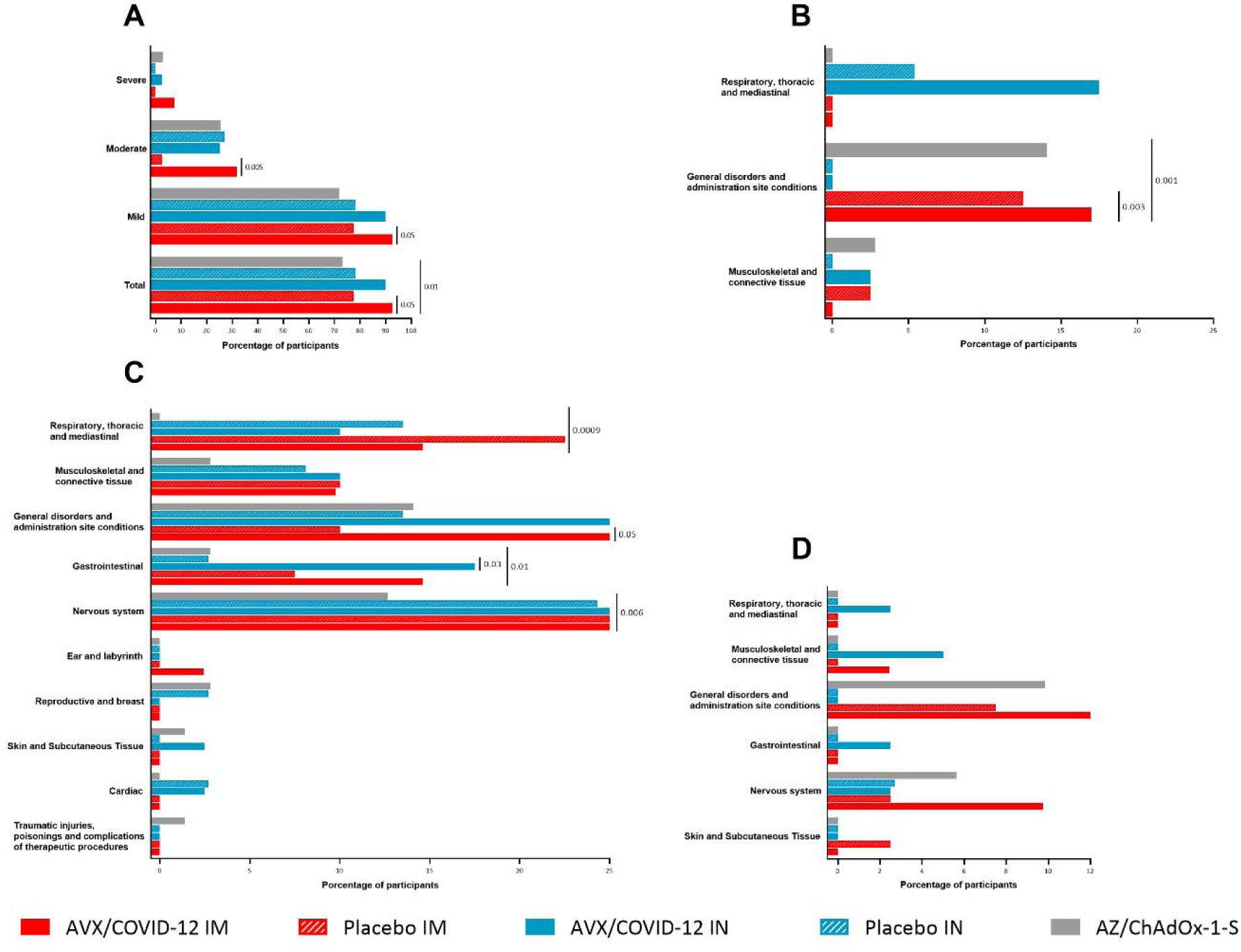
Adverse events 7 days after vaccination with AVX/COVID-12 or AZ/ChAdOx-1-S. Adverse events in participants following vaccination with the AVX/COVID-12 vaccine, compared with the AZ/ChAdOx-1-S vaccinated and placebo group. A) Percentage of participants affected by adverse events classified by severity; B) Local adverse events of special interest; C) Systemic adverse events of special interest; and D) Adverse events potentially associated with the vaccine. p-value calculated from a comparison of proportions (Z-test). Statistically significant differences were observed with p-values: p≤0.05.

After completing the follow-up period and unblinding the study, subjects initially receiving the placebo were administered a booster with the AZ/ChAdOx-1-S vaccine. The proportion of subjects reporting any AEs in the group that received the AVX/COVID-12 vaccine was significantly higher than the group that received the AZ/ChAdOx-1-S vaccine (Figure 2A). No significant differences were observed in the proportions of both groups when AEs were analyzed and grouped by severity.

Special interest local AEs reported at 7 days post-immunization, categorized by the number of affected subjects, were recorded (Table 2S). About, 19.21% of the study participants reported being affected by at least one of these events. Specifically, for each study group, this corresponds to 41.46% for the AVX/COVID-12 IM group, 15% for the IM placebo group, 17.5% for the AVX/COVID-12 IN group, 5.4% for the IN placebo group, and 16.9% for the AZ/ChAdOx-1-S group. When comparing the proportion of subjects affected by the total events (Table 2S), it was observed that AVX/COVID-12 IM had a significantly higher proportion than IM placebo (p=0.008) and AZ/ChAdOx-1S vaccine (p=0.004). This pattern was also observed for mild events, while for moderate and severe events, no statistically significant differences were found among these groups (Table 2S).

We identified statistically significant differences in AEs related to local general disorders and administration site conditions when comparing the group receiving AVX/COVID-12 IM with the AZ/ChAdOx-1-S vaccine group (p=0.001). Additionally, when the AVX/COVID-12 vaccinated group was compared with the placebo group, statistically significant differences were also observed (p=0.003) (Figure 2B). General discomfort included symptoms such as injection site pain (Table 2S).

Similarly, we analyzed systemic AEs of special interest reported within 7 days after the administration of the AVX/COVID-12 vaccine. About, 44.1% of the study participants reported being affected by one of these events. Specifically, 58.53% corresponded to the IM AVX/COVID-12 group, 50% to the IM placebo group, 57.5% to the IN AVX/COVID-12 group, 40.54% to the IN placebo group, and 26.76% to the AZ/ChAdOx-1-S group (Table 3S). When comparing groups, it was observed that the group receiving IM AVX/COVID-12 had a significantly higher proportion of subjects affected by this type of event than the AZ/ChAdOx-1-S group (p=0.0009). Analyzing events according to severity, a significant difference was found for mild events (Table 3S).

Subsequently, an analysis of events classified by system organ class (SOC) and preferred term (PT) for systemic AEs at 7 days post-immunization was performed (Table 3S). This analysis shows that the group receiving IM AVX/COVID-12 had a significantly higher proportion of subjects affected than the AZ/ChAdOx-1-S group for mild disorders of the nervous system, particularly headache (p=0.006); mild gastrointestinal disorders, particularly odynophagia (p=0.01); and mild thoracic and mediastinal respiratory disorders, particularly rhinorrhea (p=0.0009) (Figure 2C and Table 3S).

In the comparison between IM AVX/COVID-12 and placebo, a statistically significant difference was observed in the proportion of subjects affected by general disorders and site administration alterations (p=0.05). When the AVX/COVID-12 vaccine was administered IN, a significantly higher proportion of subjects showed mild gastrointestinal disorders compared to the IN placebo group (p=0.03) (Figure 2C).

Finally, regarding AEs related to vaccination, 9.17% of subjects reported AEs (Table 4S). In particular, 14.63% of subjects in the IM AVX/COVID-12 group, 10% in the IM placebo group, 7.5% in the IN AVX/COVID-12 group, 2.7% in the IN placebo group, and 9.85% in the AZ/ChAdOx-1-S group were reported. When analyzing events by SOC and PT, no statistically significant differences were observed despite variations in the proportion of subjects affected (Figure 2D).

### AVX/COVID-12 induced binding and neutralizing antibody titers against Wuhan-1 and SARS-CoV-2 variants of concern

The evaluation of antibody titers and their neutralizing capacity in sera from AVX/COVID-12 vaccinated volunteers against the spike protein was conducted using ELISA and the pseudovirus neutralization assay at 14 days post-immunization, both for the total number of volunteers vaccinated with AVX/COVID-12 IM and for subjects who did not develop COVID-19 during the initial 14 days of the study (Table 3). Subsequent assessments were carried out over one year in IM-vaccinated subjects (Table 5S and 6S) and IN (Table 7S and 8S).

**Table 3.**
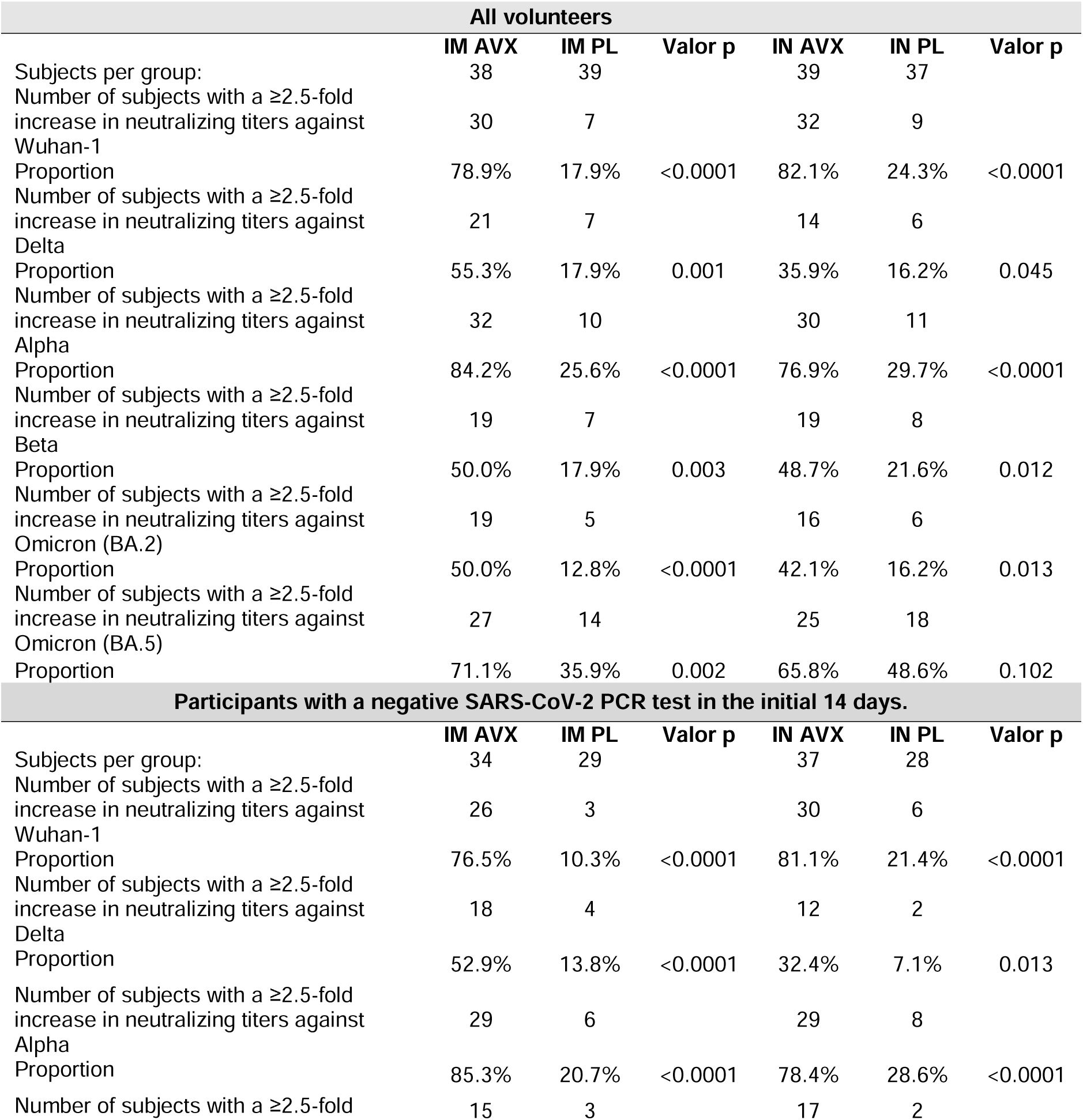
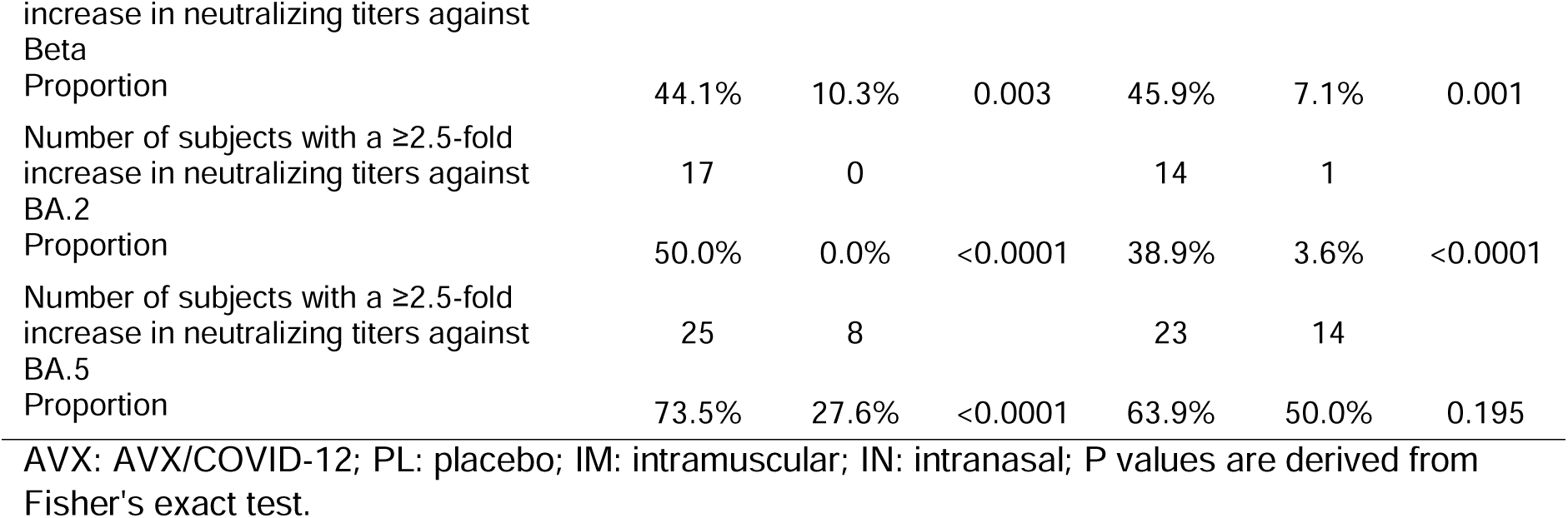
Proportion of subjects with an increase in neutralizing antibody titers against SARS-CoV-2 variants of concern 14 days after receiving AVX/COVID-12 or placebo.

**Table 4.** Proportion of subjects with an increase in neutralizing antibody titers at 14 days post-immunization against different variants of SARS-CoV-2.

|  | IM AVX | IM PL | p-Value | IN AVX | IN PL | p-Value |
| --- | --- | --- | --- | --- | --- | --- |
| Subjects per group | 38 | 39 |  | 39 | 37 |  |
| Number of subjects with $\geq 2.5$-fold increase in neutralizing antibody titers against Wuhan-1 variant | 30 | 7 | | 32 | 9 | |
| Proportion | 78.9% | 17.9% | <0.0001 | 82.1% | 24.3% | <0.0001 |
| Number of subjects with $\geq 2.5$-fold increase in neutralizing antibody titers against Delta variant | 21 | 7 | | 14 | 6 | |
| Proportion | 55.3% | 17.9% | 0.001 | 35.9% | 16.2% | 0.045 |
| Number of subjects with $\geq 2.5$-fold increase in neutralizing antibody titers against Alpha variant | 32 | 10 | | 30 | 11 | |
| Proportion | 84.2% | 25.6% | <0.0001 | 76.9% | 29.7% | <0.0001 |
| Number of subjects with $\geq 2.5$-fold increase in neutralizing antibody titers against Beta variant | 19 | 7 | | 19 | 8 | |
| Proportion | 50.0% | 17.9% | 0.003 | 48.7% | 21.6% | 0.012 |
| Number of subjects with $\geq 2.5$-fold increase in neutralizing antibody titers against BA.2 | 19 | 5 | | 16 | 6 | |
| Proportion | 50.0% | 12.8% | <0.0001 | 42.1% | 16.2% | 0.013 |
| Number of subjects with $\geq 2.5$-fold increase in neutralizing antibody titers against BA.5 | 27 | 14 | | 25 | 18 | |
| Proportion | 71.1% | 35.9% | 0.002 | 65.8% | 48.6% | 0.102 |
| **Subjects without COVID-19 in the first 14 days** |  |  |  |  |  |  |
|  | IM AVX | IM PL | p-Value | IN AVX | IN PL | p-Value |
| Subjects per group | 34 | 29 |  | 37 | 28 |  |
| Number of subjects with $\geq 2.5$-fold increase in neutralizing antibody titers against Wuhan-1 variant | 26 | 3 | | 30 | 6 | |
| Proportion | 76.5% | 10.3% | <0.0001 | 81.1% | 21.4% | <0.0001 |
| Number of subjects with $\geq 2.5$-fold increase in neutralizing antibody titers against Delta variant | 18 | 4 | | 12 | 2 | |
| Proportion | 52.9% | 13.8% | <0.0001 | 32.4% | 7.1% | 0.013 |
| Number of subjects with $\geq 2.5$-fold increase in neutralizing antibody titers against Alpha variant | 29 | 6 | | 29 | 8 | |
| Proportion | 85.3% | 20.7% | <0.0001 | 78.4% | 28.6% | <0.0001 |
| Number of subjects with $\geq 2.5$-fold increase in neutralizing antibody titers against Beta variant | 15 | 3 | | 17 | 2 | |
| Proportion | 44.1% | 10.3% | 0.003 | 45.9% | 7.1% | 0.001 |
| Number of subjects with $\geq 2.5$-fold increase in neutralizing antibody titers against BA.2 | 17 | 0 | | 14 | 1 | |
| Proportion | 50.0% | 0.0% | <0.0001 | 38.9% | 3.6% | <0.0001 |
| Number of subjects with $\geq 2.5$-fold increase in neutralizing antibody titers against BA.5 | 25 | 8 | | 23 | 14 | |
| Proportion | 73.5% | 27.6% | <0.0001 | 63.9% | 50.0% | 0.195 |

AVX: AVX/COVID-12; PL: placebo; IM: intramuscular; IN: intranasal; P values are derived from Fisher's exact test.

The antibody response was analyzed for a year following immunization with the AVX/COVID-12 vaccine via intramuscular and intranasal routes. Significant differences are observed at all time points for anti-spike IgG antibodies in vaccinated participants; titers increased at all analyzed time points compared to baseline levels (Figure 3A), with the highest levels observed on day 14 post-immunization, followed by a gradual decline. Interestingly, a slight increase is observed on day 365, possibly due to SARS-CoV-2 infections during participants’ follow-up. When analyzing the population, excluding those who developed mild COVID-19 during the study period (Figure 3B), a similar antibody kinetics was observed. Additionally, when analyzing IgG anti-S titers in volunteers vaccinated IN, a comparable pattern is observed when excluding subjects positive for COVID-19 (Figure 3B).

**Figure 3.**
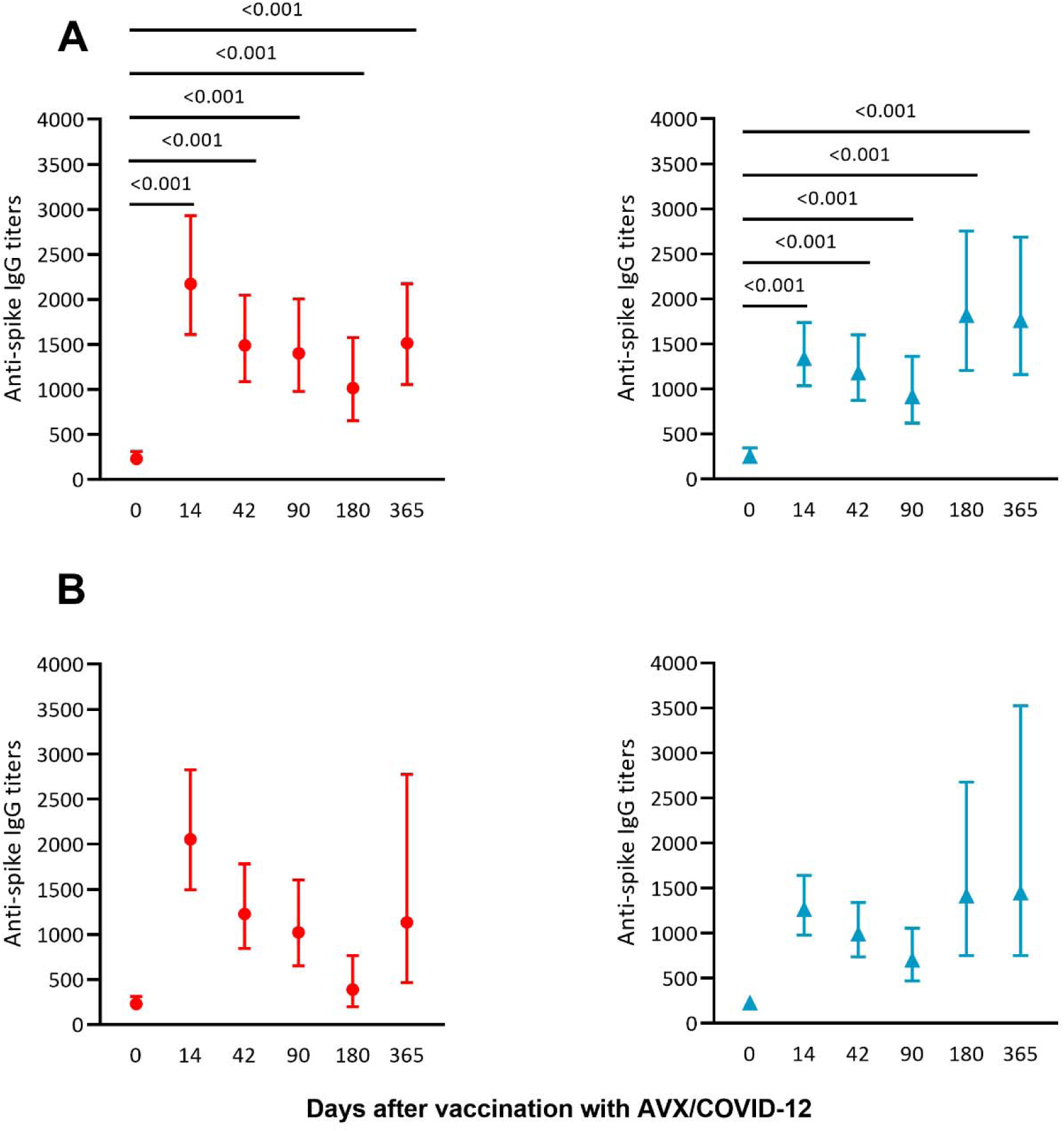
Intramuscular and intranasal administration of the AVX/COVID-12 vaccine resulted in comparable anti-spike IgG antibody titers. Geometric means and confidence intervals of serum antibody titers post-vaccination with the AVX/COVID-12 vaccine were assessed A) intramuscularly (red circles) and intranasally (blue triangles) throughout the study and B) intramuscularly (red circles) and intranasally (blue triangles) in subjects where COVID-19 was not detected during the study follow-up period. P values were calculated by comparing the geometric means using the Wilcoxon signed-rank test with variables presented on a logarithmic scale.

AVX/COVID-12 induced a ≥ 4-fold increase in anti-spike IgG titers in 73.7% of the subjects receiving IM immunization when compared to placebo (10%) (p<0.0001). In contrast, individuals receiving the vaccine IN showed a ≥ 4-fold increase of 51.3%, compared to 16.2% in the placebo group (p=0.001) (Figure 4A). Similarly, 76.5% of subjects who did not experience COVID-19 in the first 14 days post-immunization and received IM AVX/COVID-12 showed a ≥ 4-fold increase compared to 0% in the placebo group (p<0.0001). Meanwhile, 48.6% of volunteers who received the vaccine IN exhibited a ≥ 4-fold increase compared to 7.1% in the placebo group (p<0.0001) (Figure 4B).

**Figure 4.**
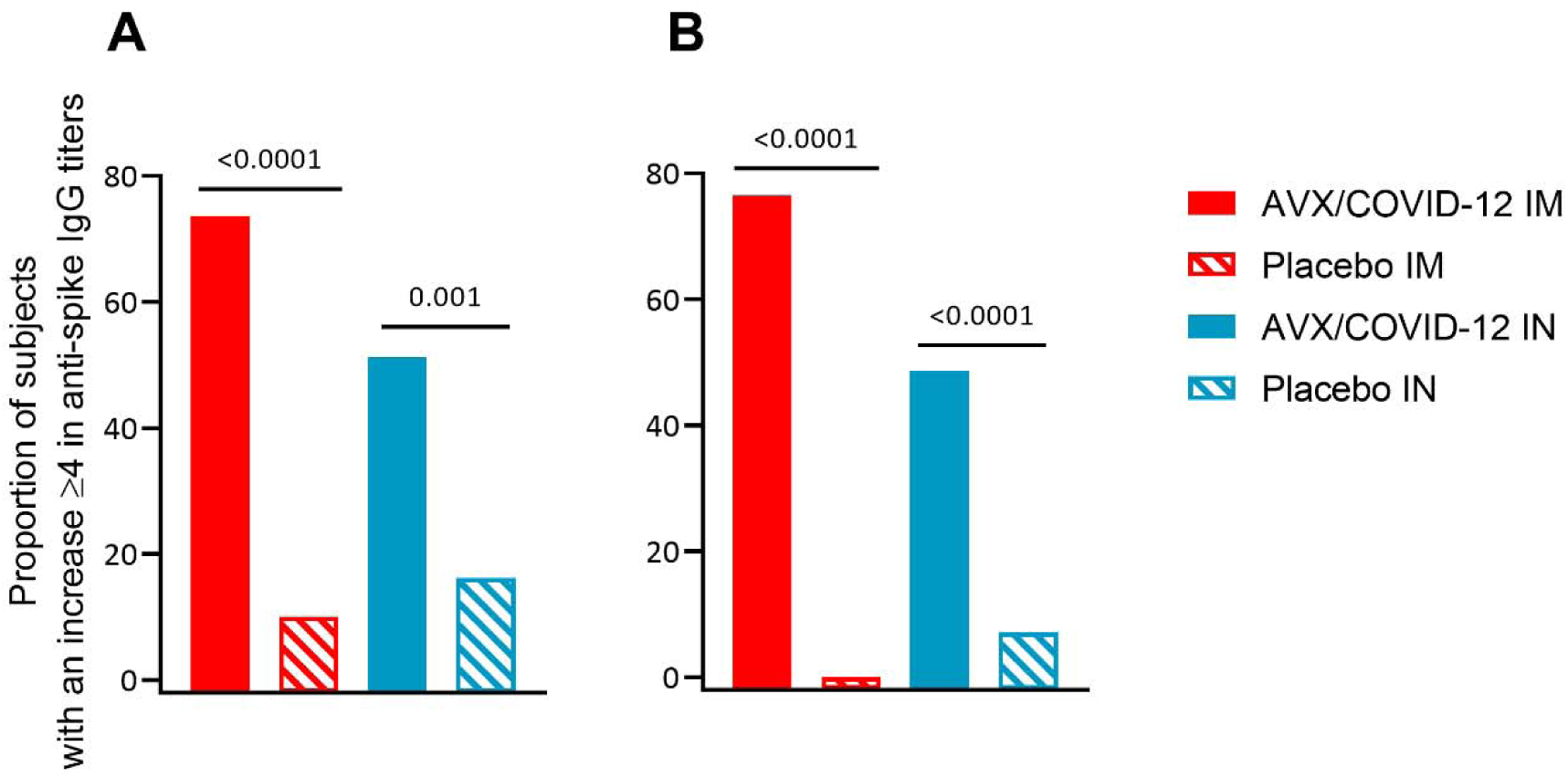
Proportion of subjects with a ≥4-fold increase in anti-spike IgG antibody titers. (A) All enrolled study participants and (B) subjects where SARS-CoV-2 infection was not detected during the 14 days after vaccination. P values were calculated using Fisher’s exact test.

Subsequently, neutralizing antibody titers against SARS-CoV-2 Wuhan-1 and Alpha, Beta, Delta, and Omicron BA.2 and BA.5 VOCs were analyzed. For the IM-administered AVX/COVID-12 vaccine, statistically significant differences in the neutralizing antibody responses were observed when compared to the placebo in all cases (Figure 5 and Table 5S). In contrast, in subjects who received IN immunization with AVX/COVID-12, statistically significant differences in the neutralizing antibody responses were only observed for Wuhan-1 and Alpha (Figure 5). In the case of subjects who did not present COVID-19 in the first 14 days, all groups immunized IM or IN showed statistically significant differences in the neutralizing antibody responses when compared to the placebo group, except for the anti-BA.5 neutralizing responses in the IN immunized group (Figure 5 and Table 6S).

**Figure 5.**
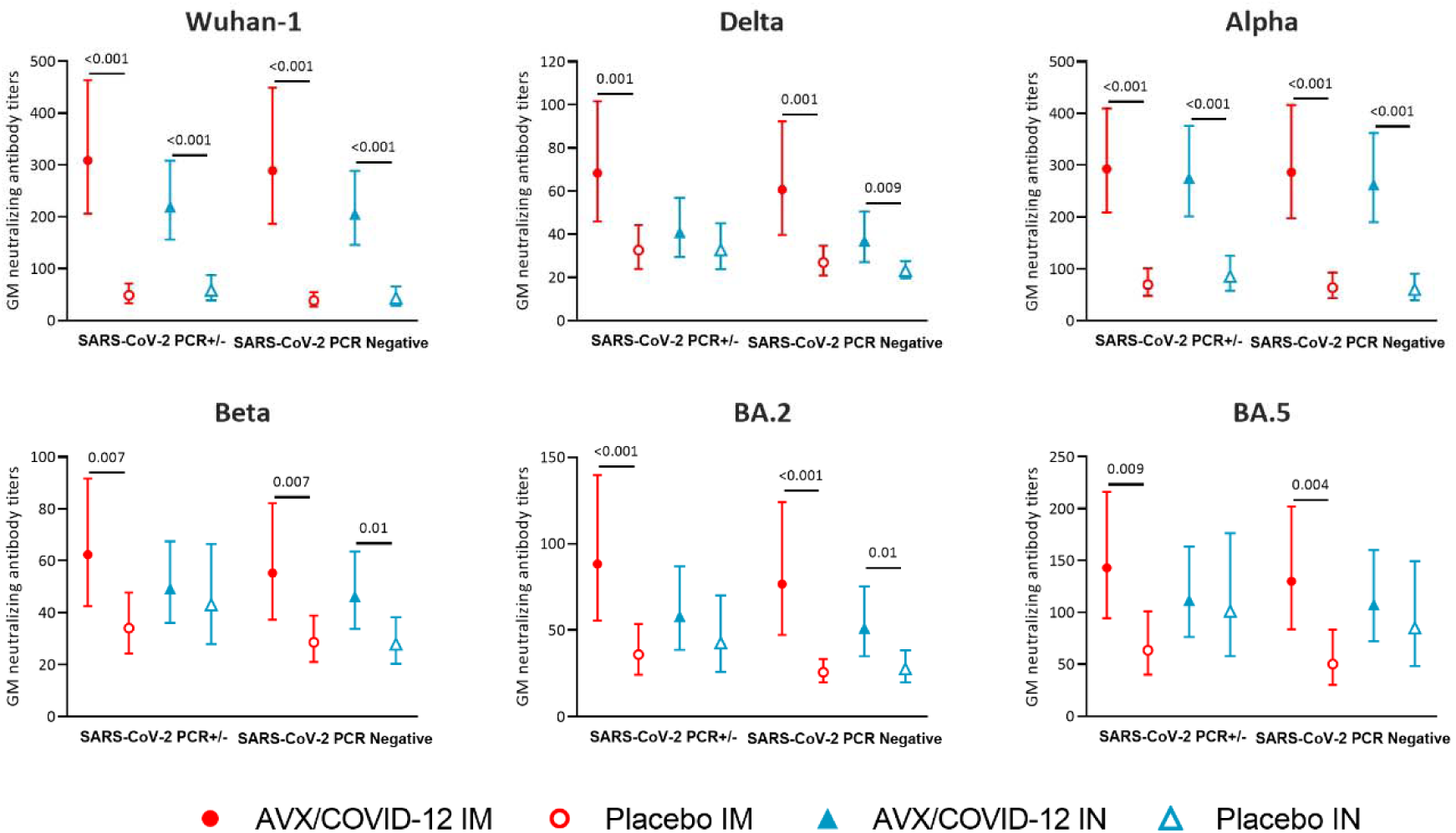
Comparison of neutralizing antibody titers against SARS-CoV-2 variants of concern (VOC) in subjects 14 days after receiving AVX/COVID-12 or placebo. Geometric mean comparison of neutralizing antibody titers against Wuhan, Delta, Alpha, Beta, and Omicron (BA.2 and BA.5) VOCs in volunteers after 14 days of receiving AVX/COVID-12 intramuscular (IM) (closed red circles) or IM placebo (open red circles) and AVX/COVID-12 intranasal (IN) (closed blue triangles) or IN placebo (open blue triangles). Analysis was performed on the total of vaccinated individuals, regardless of SARS-CoV-2 infection confirmation (SARS-CoV-2 PCR+/-), or in subjects who did not test positive for SARS-CoV-2 in a PCR test (SARS-CoV-2 PCR Negatives). Mann–Whitney U test with variables presented on a natural logarithmic scale.

In addition, the proportion of subjects who exhibited a ≥ 2.5-fold increase in neutralizing antibody titers against Wuhan-1 and the aforementioned VOC was determined. For subjects who received AVX/COVID-12 IM immunization, the proportions of those exhibiting a ≥ 2.5-fold increase in neutralizing antibody titers range from 84.2% to 50% (Alpha > Wuhan-1 > Omicron BA.5 > Delta > Omicron BA.2 > Beta), compared with subjects who received IM placebo, where 35.9% to 12.8% exhibited a ≥ 2.5-fold increase in neutralizing antibody titers (p < 0.0001 to 0.003) (Figure 6A). For those vaccinated with AVX/COVID-12 IN, the percentage of individuals who exhibited a ≥ 2.5-fold increase in neutralizing antibody titers ranges from 82.1% to 35.9% (Wuhan-1 > Alpha > Omicron BA.5 > Beta > Omicron BA.2 > Delta) (p > 0.0001 to 0.045), except for the anti-BA.5 neutralizing antibody response, where no statistically significant differences with the placebo were observed (Figure 6A).

**Figure 6.**
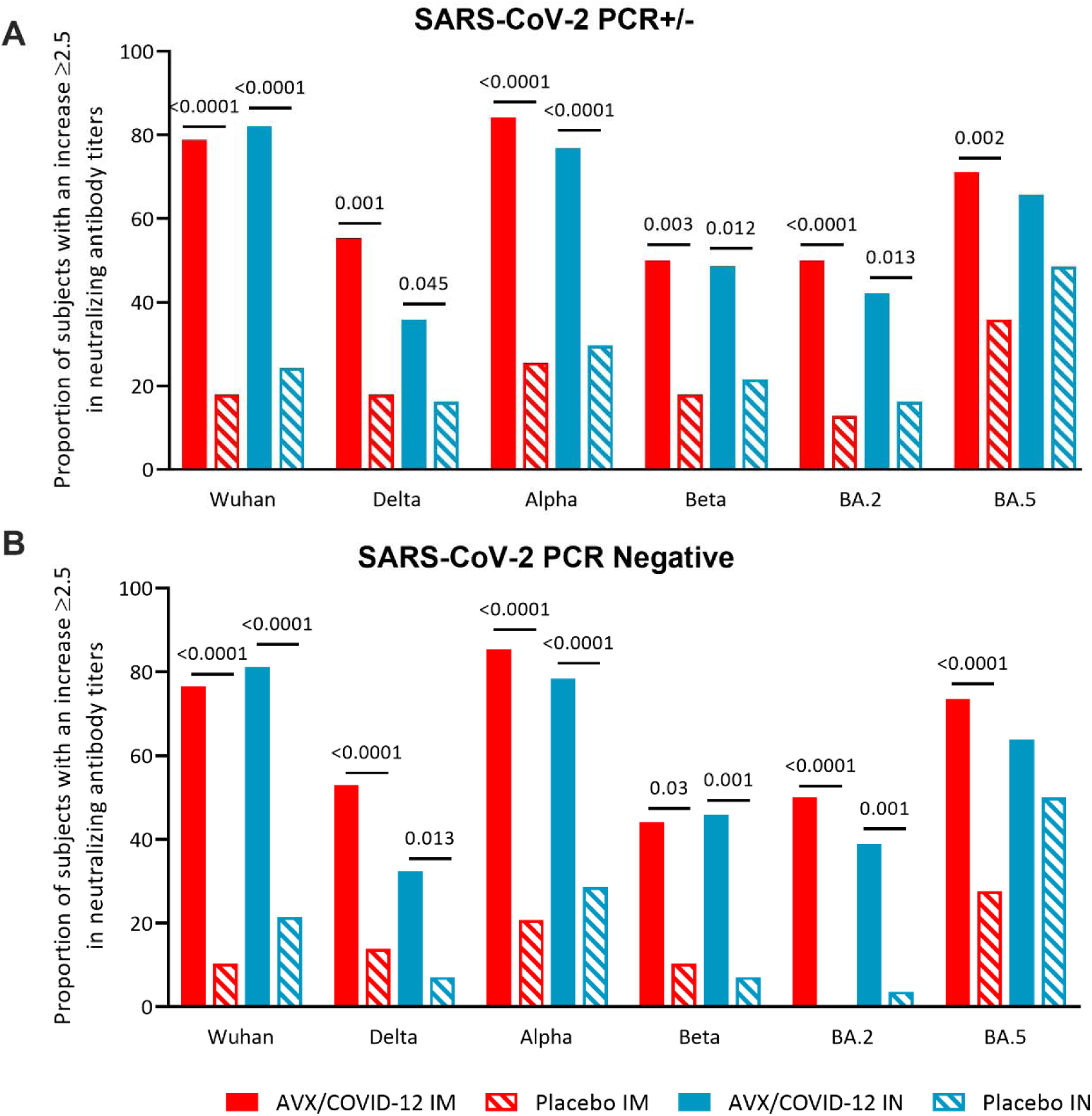
Proportion of subjects with a ≥2.5-fold increase in neutralizing titers against SARS-CoV-2 variants of concern induced by AVX/COVID-12 immunization. Neutralizing antibody titers against Wuhan, Delta, Alpha, Beta, and Omicron (BA.2 and BA.5) variants of concern were measured 14 days after intramuscular (IM) (red closed bars) or intranasal (IN) (blue closed bars) AVX/COVID-12 administration, and IM (striped red bars) or IN (striped blue bars) placebo administration. A) In the total of vaccinated individuals, regardless of SARS-CoV-2 infection confirmation (SARS-CoV-2 PCR +/-) or B) in subjects who did not test positive for SARS-CoV-2 in a PCR test (SARS-CoV-2 PCR Negatives). P values were calculated using Fisher’s exact test.

In the sub-analysis, only subjects who did not present with COVID-19 in the first 14 days post-immunization were considered. For subjects who received AVX/COVID-12 IM, the percentage of those exhibiting a ≥ 2.5-fold increase in neutralizing antibody titers ranged from 85.3% to 44.1% (Alpha > Wuhan-1 > Omicron BA.5 > Delta > Omicron BA.2 > Beta), compared with volunteers administered a placebo, ranging from 35.9% to 12.8% (p > 0.0001 to 0.03). Among the group vaccinated IN, the percentage of individuals who exhibited a ≥2.5-fold increase in neutralizing antibody titers ranged from 81.1% to 32.4% (Wuhan-1 > Alpha > Omicron BA.5 > Beta > Omicron BA.2 > Delta). However, except for of the anti-BA.5 neutralizing antibody responses, no statistically significant difference was observed when compared with the placebo group (Figure 6B). Nevertheless, it should also be noted that the IN placebo control group presented an unusually higher rise of anti-BA.5 specific antibodies as compared to the other VOCs, perhaps indicative of a recent infection.

### Comparable IFN-γ positive cellular responses were identified among participants immunized with AVX/COVID-12, regardless of the administration route

IFN-γ production was measured from supernatants of PBMCs obtained from volunteers vaccinated IM or IN (Tables 5 and 6) with the AVX/COVID-12 vaccine. Cells were collected on days 0, 14, 180, and 365 (Table 7) and stimulated *in vitro* with purified SARS-CoV-2 spike protein subunit 1 (Wuhan-1). In the total vaccinated individuals, regardless of SARS-CoV-2 infection and/or COVID-19 confirmation (SARS-CoV-2 PCR+/- and/or COVID-19+/-) (Table 7), statistically significant differences in IFN-γ production were observed at day 365 for IM-vaccinated individuals (p < 0.01) and at days 180 (p < 0.03) and 365 (p < 0.01) in IN-vaccinated individuals (Figure 7A).

**Table 5.** IFN-γ secreted (pg/mL) by T lymphocytes, comparison on day 14 between AVX/COVID-12 and placebo.

| All volunteers |  |  |  |  |  |  |
| --- | --- | --- | --- | --- | --- | --- |
|  | IM AVX | IM PL | p-Value | IN AVX | IN PL | p-Value |
| Subjects | 20 | 35 |  | 20 | 33 |  |
| GM | 324.2 | 258.6 |  | 193.7 | 406.7 |  |
| CI | 166.5 - 631.4 | 123.1 - 543.2 | 0.97 | 73 - 513.4 | 262 - 631.4 | 0.31 |
| Volunteers without COVID-19 in the first 14 days. |  |  |  |  |  |  |
|  | IM AVX | IM PL | p-Value | IN AVX | IN PL | p-Value |
| Subjects | 17 | 27 |  | 19 | 27 |  |
| GM | 280 | 168.6 |  | 175.5 | 358 |  |
| CI | 133.3 - 588.1 | 71.2 - 399.2 | 0.71 | 64 - 481 | 220 - 581 | 0.33 |

AVX: AVX/COVID-12; PL: placebo; IM: intramuscular; IN: intranasal; GM: geometric mean; CI: confidence interval. Comparisons of GM at day 14 between groups, p values were calculated using the Mann-Whitney's U test.

**Table 6.**
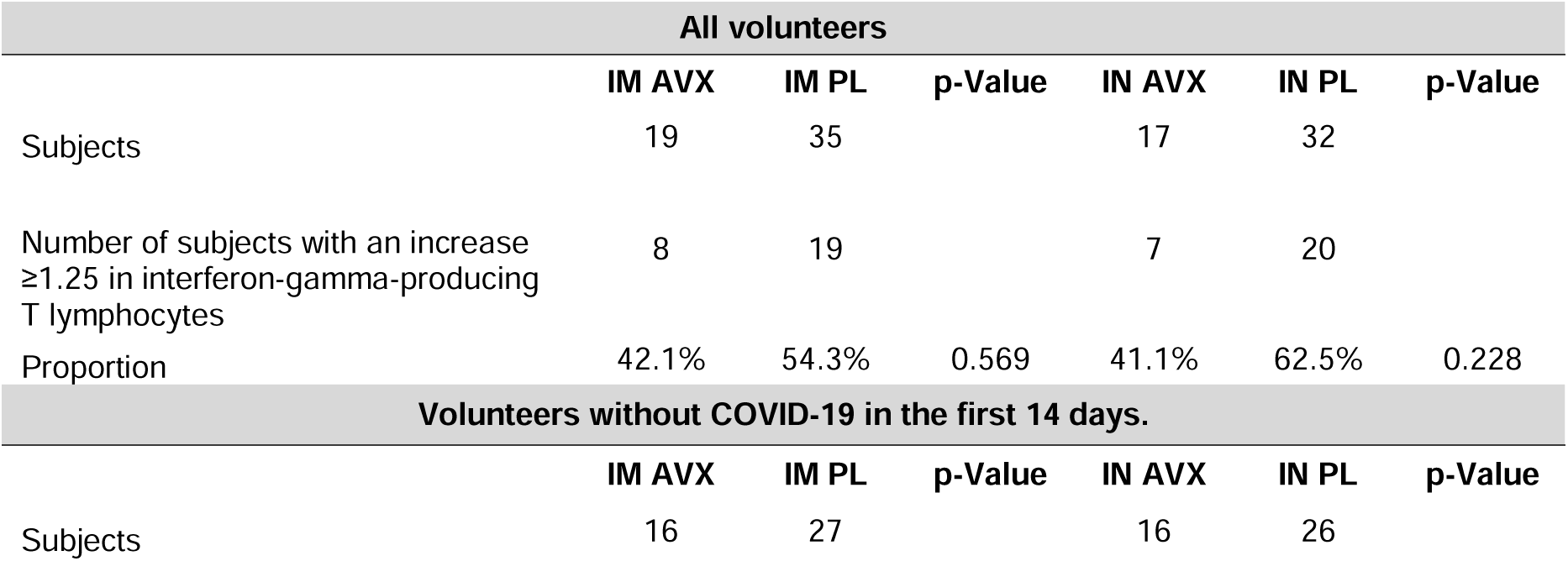
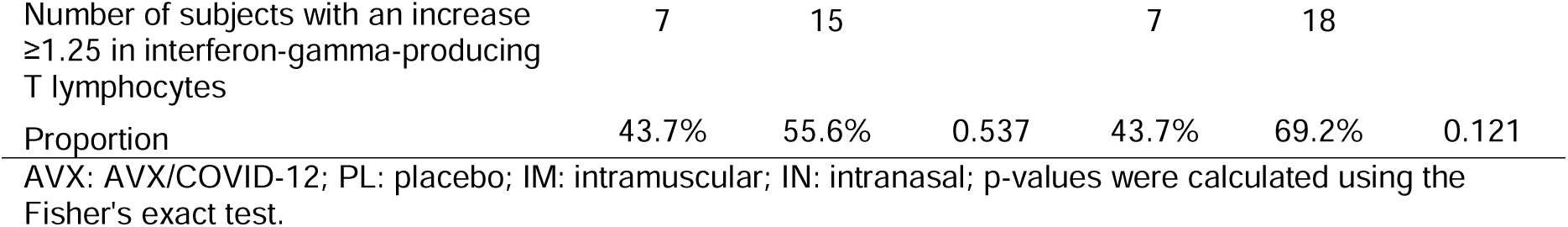
Proportion of subjects with an increase in IFN-γ secretion by peripheral blood T lymphocytes in response to stimulation with the S protein of SARS-CoV-2.

**Table 7.** IFN-γ secreted (pg/mL) by T lymphocytes in subjects vaccinated with AVX/COVID-12 throughout the study.

| All volunteers |  |  |  |  |
| --- | --- | --- | --- | --- |
| AVX IM |  |  |  |  |
|  | Baseline | Day 14 | Day 180 | Day 365 |
| Subjects | 24 | 20 | 39 | 21 |
| GM | 235.8 | 324.2 | 867.1 | 1160.1 |
| CI | 107.1 - 519.2 | 166.5 - 631.4 | 602 - 1249.1 | 746- 1804.1 |
| p-Value |  | 0.99 | 0.09 | 0.01 |
| AVX IN |  |  |  |  |
|  | Baseline | Day 14 | Day 180 | Day 365 |
| Subjects | 20 | 20 | 34 | 18 |
| GM | 241.4 | 193.7 | 951.2 | 1011.3 |
| CI | 81.75248 713.1 | 73 - 513.4 | 648.5 - 1395.3 | 642.2 - 1592.4 |
| p-Value |  | 0.83 | 0.03 | 0.01 |

AVX: AVX/COVID-12; IM: intramuscular; IN: intranasal; GM: geometric mean; CI: confidence interval. Comparison of GM using the Wilcoxon signed-rank test.

**Figure 7.**
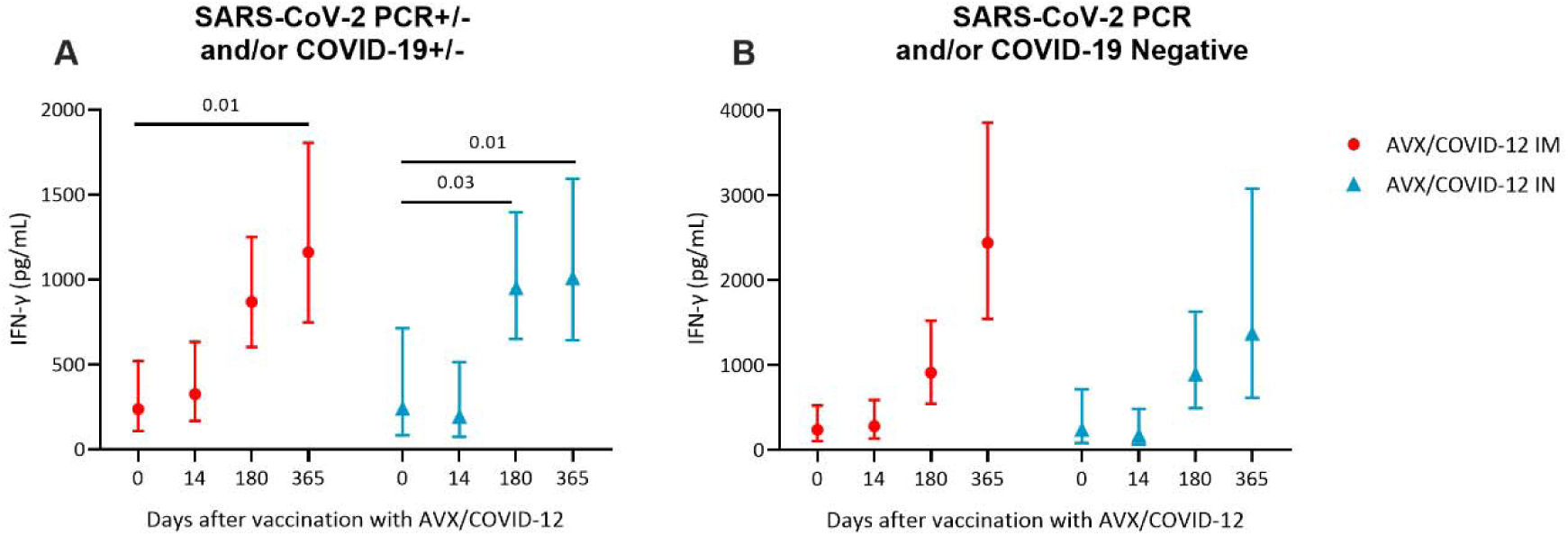
Cellular responses induced by the AVX/COVID-12 Vaccine. IFN-γ production was measured from peripheral blood mononuclear cells (PBMCs) of volunteers immunized intramuscularly (IM) with red circles or intranasally (IN) with blue triangles using the AVX/COVID-12 vaccine at the indicated time points. Volunteers’ PBMCs were stimulated for 18 hours with purified SARS-CoV-2 S1 protein (Wuhan-1), and IFN-γ production was measured from cell culture supernatants using BioPlex. A) In the total vaccinated individuals, regardless of SARS-CoV-2 infection and/or COVID-19 confirmation (SARS-CoV-2 PCR+/- and/or COVID-19+/-), or B) in subjects who did not test positive for SARS-CoV-2 in a PCR test and/or COVID-19 confirmation (SARS-CoV-2 PCR and/or COVID-19 Negative). Significant differences were calculated with the Wilcoxon signed-rank test.

Meanwhile, in subjects who did not test positive for SARS-CoV-2 in a PCR test and/or COVID-19 confirmation (SARS-CoV-2 PCR and/or COVID-19 negative) (Table 8), the response observed at day 365 was higher compared to the SARS-CoV-2 PCR+/- and/or COVID-19+/- group for both IM and IN immunized individuals. A trend towards increased IFN-γ concentration was evident in both IM and IN immunized groups with the AVX/COVID-12 vaccine; however, no statistically significant differences were observed (Figure 7B).

**Table 8.** IFN-γ secreted (pg/mL) by T lymphocytes in subjects without COVID-19 vaccinated with AVX/COVID-12 throughout the study.

| Volunteers without COVID-19 |  |  |  |
| --- | --- | --- | --- |
| AVX IM |  |  |  |
|  | Día 14 | Día 180 | Día 365 |
| Subjects | 3 | 24 | 17 |
| GM | 744.6 | 842.6 | 974 |
| CI | 40.4 - 13715.6 | 496.9 - 1428.9 | 583.9 - 1624.7 |
| AVX IN |  |  |  |
|  | Día 14 | Día 180 | Día 365 |
| Subjects | 19 | 18 | 8 |
| GM | 175.5 | 895.7 | 1374.6 |
| CI | 64 - 481 | 493.1 - 1626.9 | 614.3 - 3075.7 |

AVX: AVX/COVID-12; IM: intramuscular; IN: intranasal; GM: geometric mean; CI: confidence interval.

### Immunization with the AVX/COVID-12 vaccine induces notably low specific antibody titers against the NDV vector

To assess the immune response elicited against the NDV vaccine vector, we evaluated the specific antibody titers exhibiting hemagglutination inhibition (HAI) activity in serum samples from individuals participating in phase I and II studies.

No antibodies with specific HAI against NDV were detected in all serum samples from individuals who participated in the phase II study on day 0 of recruitment, before the administration of the AVX/COVID-12 vaccine as a booster dose. Samples above the cut-off threshold of 8 HAI titers were considered positive. On day 14 following the initial administration of the AVX/COVID-12 vaccine, only two samples displayed titers above 8, indicating seroconversion. Meanwhile, on days 42 and 90, 4 and 3 out of 144 sera exhibited titers equal to or greater than 8, respectively. These results demonstrate that the administration of the AVX/COVID-12 vaccine at a dose of 10^8.0^ EID50/dose via the IM route induces a low specific seroconversion to NDV (Figure 8).

**Figure 8.**
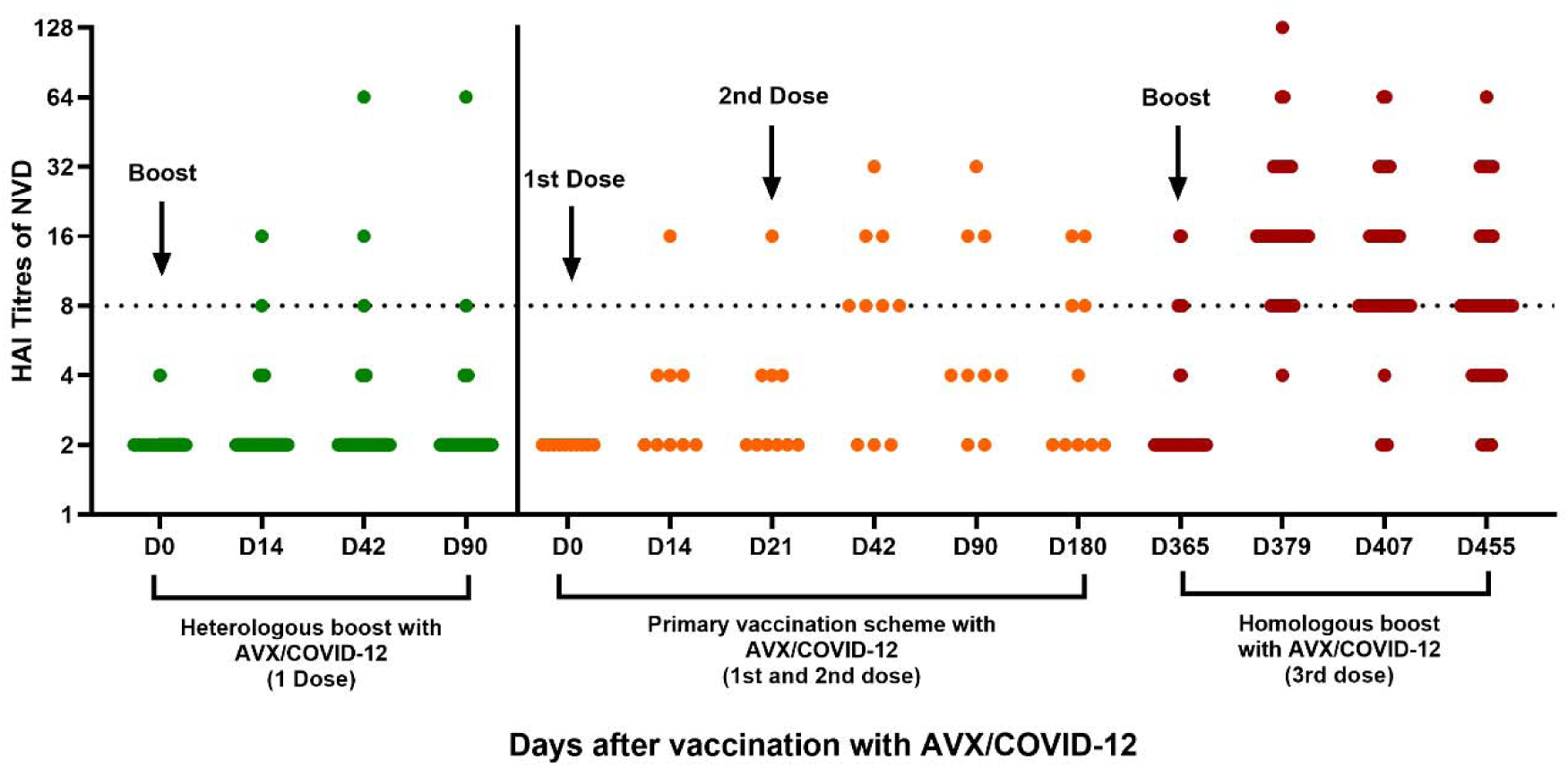
Immunization with the AVX/COVID-12 vaccine induces notably low specific antibody titers against the NDV vector. Analysis of hemagglutination inhibition antibody (HAI) titers specific to the Newcastle disease virus (NDV) vector in sera from individuals who received the AVX/COVID-12 vaccine as a booster dose in the phase II clinical trial (green circles, n=154) or as part of the primary immunization scheme with two doses on day 0 and a booster on day 21 (orange circles, n=10), and after a homologous booster dose on day 365 in the phase I clinical trial (red circles, n=43). Samples above the cut-off threshold of 8 HAI titers were considered positive (doted line).

In addition, serum samples from individuals participating in phase I, who received two doses of 10^8.0^ EID50/dose of the AVX/COVID-12 vaccine, were analyzed. It was observed that all serum samples were negative for specific antibodies to the NDV vector before the first immunization via IN or IM administration. On day 14 after immunization, only 1 sample was positive. Meanwhile, titers higher than 8 were detected after the second application of the AVX/COVID-12 vaccine (Day 42). On the 90th day of follow-up, only 4 out of 10 sera showed titers higher than 8. After one year, volunteers received a third dose as a boost. Before boosting, only 6 volunteers had antibody titers equal to or greater than 8. After boosting, 42 out of 43 samples were positive, most showing low titers against NDV. Seroconversion slowly faded, and on day 90 after the boost, only 28 samples showed low antibody titers (Figure 8). Collectively, the data reveal that the induction of NDV-specific antibodies is exceedingly low (less than 0.2%) in the studied population after a single dose of the AVX/COVID-12 vaccine. Following the second and third immunizations, seroconversion increased; nevertheless, antibody titers remained at a low level. It is important to emphasize that the technique developed with highly sensitive.

## Discussion

SARS-CoV-2 and its VOCs have had a staggering effect on human health worldwide. Despite the development and rapid deployment of vaccines against SARS-CoV-2, many countries, particularly low and middle-income countries, have not yet met their vaccination needs. This underscores the urgency of establishing vaccine development and production capabilities in these regions to contribute to a quicker onset of global community immunity against severe disease.

The periodic administration of booster doses emerges as an essential measure to sustain community immunity against severe disease, reducing the magnitude of future waves of COVID-19 and safeguarding vulnerable populations, including the elderly, immunocompromised individuals, and those with comorbidities (17).

In this phase II clinical trial, we assessed the safety and immunogenicity of the AVX/COVID-12 vaccine as a booster dose in volunteers who had previously received COVID-19 vaccines based on adenoviral technology, inactivated virus, or mRNA technology and exhibited antibody titers lower than 1,200 U/mL. These titers were considered sufficiently low to observe the booster effect induced by the IM or IN administration of AVX/COVID-12. Recruitment presented particular challenges due to the prevailing infections with Delta and Omicron variants (BA.1 to BA.5) in Mexico at the time of the trial, resulting in a high prevalence of antibody titers in the population (Figure 1S).

After screening 1,221 volunteers, we ultimately enrolled 158 subjects who met the inclusion criteria (Figure 1). The IM or IN booster immunization with AVX/COVID-12 was safe and well-tolerated for all individuals analyzed; most of the AEs were mainly mild, and fewer were of moderate extent (Figure 2A). The vaccine exhibited appropriate tolerability, and its safety profile did not raise any concerns, irrespective of the volunteers’ initial vaccination history (Figure 2B-C). AEs potentially associated with the vaccine were also mild and occurred in a small number of the volunteers (Figure 2D). Additionally, the safety profile of AVX/COVID-12 was similar to that observed for AZ/ChAdOx-1-S (Figure 2), the most widely used vaccine in Mexico and worldwide (18,19).

We observed a substantial increase in spike binding (Figures 3 and 4) and neutralizing antibody titers (Figure 5) among all volunteers following booster doses of AVX/COVID-12 administered through IM and IN routes. Specific anti-spike IgG slowly decayed during the following six months, but titers remained high and increased one year after vaccination, perhaps due to viral exposure (Figure 3). As described above, the trial was performed during a pandemic infection wave; therefore, some volunteers were infected (no hospitalizations or deaths were observed). The antibody titer increase held true irrespective of the volunteers’ initial vaccination history, involving vaccines based on adenoviral technology, inactivated virus, or mRNA technology.

In the case of influenza vaccines, a ≥ 4-fold increase in HAI titers is considered a successful seroconversion (20), and neutralizing antibody titers have also been correlated with protection for SARS-CoV-2 (21). Here, we observed a high proportion of subjects who received the AVX/COVID-12 vaccine showing a ≥ 4-fold increase in anti-S IgG titers. As anticipated, the IM route of immunization proved to be more efficient than the IN route in inducing serum antibody titers (Figure 4).

Real-world evidence indicates that first-generation vaccines, utilizing the spike protein from the original Wuhan-1 strain, exhibit high effectiveness against the severity of infection (intensive care unit (ICU) admission or death) caused by various subvariants of Omicron. For instance, a full vaccination scheme with a booster from Moderna shows 95% effectiveness, Pfizer demonstrates 95-100% (22,23), while AstraZeneca or Janssen exhibits 85-90% effectiveness (23). Additionally, the mRNA vaccine BNT162b2 has been reported to be 84.9% effective in preventing intensive care unit admission for COVID-19 in children and 95% in adolescents (24). Other studies indicate a 90.9% effectiveness of this vaccine against hospitalization and death caused by the Omicron BA.1 and BA.2 variants of SARS-CoV-2, while the AZ/ChAdOx-1-S vaccine showed 82.3% effectiveness (25). Moreover, COVID-19 vaccines available in Mexico have been reported to significantly reduce the risk of severe disease during the Omicron wave in the country (26).

Our results are in concordance with these observations: AVX/COVID-12 induced neutralizing antibody responses not only against the Wuhan-1 original strain but also against the Alpha, Beta, Delta, and Omicron (BA.2 and BA.5) variants (Figure 5). It was particularly surprising to observe a higher proportion of individuals with a ≥ 2.5 increase in neutralizing titers against the BA.5 compared to BA.2, despite recruitment being performed during the BA.1-4 and BA.5 pandemic waves (Figure 1S). These responses might be explained by the generation of hybrid immunity induced by the combination of infection and vaccination or vice versa (3,4). In addition, a significant cross-reactive response of both antibodies and T cells generated by infection or vaccination has been identified; in the case of hybrid immunity, these cross-reactive responses are heightened and broadened (3,4,27–29). These findings align with the description of a large set of conserved epitopes in the spike protein for both humoral and cellular responses among SARS-CoV-2 Wuhan-1 and VOCs, as well as SARS-CoV, MERS-CoV, and seasonal coronaviruses, and even within all those in the *Orthocoronavirinae* group (30).

Across all study groups, an increase in IFN-γ levels at later time points is observed, suggesting a potential response to infections during the study (Figure 6). This phenomenon could be explained by the development of hybrid immunity, where the cellular response, through constant restimulations, may enhance both the magnitude of effector capacity (cytotoxicity and cytokine production) and the long-term memory response (31,32).

We observed a tendency toward a more pronounced IFN-γ response in individuals who recovered from the infection (Figure 7). This response can be attributed to the development of antigen-specific T cells and memory cells targeting viral antigens of the virus, as the antigen-specific stimulation utilizes the spike protein of the virus. We have found that these cells can persist in circulation despite decreasing antibody titers. Our study shows lower antibody titers at 180 and 365 days compared to those observed 14 days after the first immunization (Figures 3 and 7). This increase might also be attributed to continuous, asymptomatic contact with SARS-CoV-2, implying the observed re-stimulation at day 365 (Figures 3 and 7). It is crucial to note the study’s limitation in quantifying IFN-γ using BioPlex, which analyzes IFN-γ production derived from PBMCs stimulated with the spike protein, including natural killer cells (NK), CD4+ and CD8+ T cells, and monocytes.

A significant concern when using vector vaccines is the potential development of immune responses against the vector that could hinder its use in further immunizations. In this sense, it is essential to note that seroconversion against the NDV vector was almost absent after one or two immunizations, and the three-dose immunization scheme induced low and transient antibody titers (Figure 8). This suggests that the AVX/COVID-12 vaccine could be used repeatedly without losing efficacy.

## Conclusions

The administration of the AVX/COVID-12 “Patria” vaccine as a booster dose, either IM or IN, was deemed safe and well-tolerated. This resulted in the extension of immune responses, not only against the Wuhan-1 original strain but also against other VOCs. The vaccine effectively enhanced preexisting immune responses, supporting its use for boosting vaccination programs to expand and sustain community immunity against severe outcomes in the population. These compelling findings strongly advocate incorporating AVX/COVID-12 as an additional booster dose for the general population.

## Data Availability

The protocol was registered in the National Registry of Clinical Studies under number RNEC2021-AVXSARSCoV2VAC002 and published under NCT05205746. Individual de-identified participant data will not be shared beyond the limits permitted by the informed consent and Mexican law. Specifically, this includes the sharing of the study protocol, statistical analysis plan, informed consent form, and approved clinical study report. Additionally, other de-identified data allowed under the informed consent and Mexican law may be shared. The data will be made available immediately upon publication and for 12 months thereafter. Access to the data will be granted solely to investigators with methodologically sound proposals, subject to authorization by an independent review committee and the ethics committees involved in approving the protocol. If required by law, authorization from the Federal Commission for the Protection against Sanitary Risks (COFEPRIS) in Mexico will also be obtained. Any use of the data must strictly adhere to the authorized purposes outlined during the approval process.

## Acknowledgments

From CONAHCYT, we acknowledge María Elena Álvarez-Buylla Roces and Delia Aideé Orozco Hernández for their role as an inter-institutional liaison and overall facilitation of proceedings, committee evaluations, sanitary setup, and follow-up of trial design approvals, evaluations, and progress. We additionally acknowledge the broader support from various teams within all the clinical research sites and iLS Clinical Research S. C., particularly Gabriela Rosas, Daniela Castro, Claudia Aguilar, Roman Jarosch, and Alejandro Arias. From Laboratorio Avi-Mex, S. A. de C. V., we acknowledge the following people for their operative support: Bernardo Lozano Alcántara, Alejandro Ruiz, Rosalba Rodríguez, Leticia Espinosa Gervasio, Rodrigo Yebra Reyes, Vanessa Escamilla Jiménez, Jaime Becerra Jiménez, Lorena Juárez Pedraza, Sandra Yuridia Ang Tinajero, Avelia Ariadna Cuevas Cifuentes, Juan Pablo Robles Álvarez, Avirán Almazán Gutiérrez, Aurora Betsabé Gutiérrez Balderas, Merlenne Rubio Díaz, and Guadalupe Aguilar Rafael. From INER, we acknowledge the following people for their technical support: Liliana Figueroa Hernández, Francisco Cruz Flores, Claudia Ivett Hernández Lázaro, María Angelica Velázquez González, Jessica Romero Rodríguez, Dulce Cinthia Soriano Hernández, Milton Nieto Ponce, Edgar Reyna, Itzel Corona, José E. Márquez, Angelica Moncada, and Pablo Franco-Mendoza. We acknowledge Sean Whelan from the Icahn School of Medicine at Mount Sinai. From Instituto Politécnico Nacional, Escuela Nacional de Ciencias Biológicas, we acknowledge Sonia Mayra Pérez Tapia, Alexis Gabriel Suárez Gómez, and Sandra Comparán Alarcón.

## Ethical responsibilities

The protocol was approved by the ethics, research, and biosafety committees of each clinical research site, National Committee for Science, Technology and Innovation in Public Health of the National Council for Humanities, Science and Technology (CONAHCYT), and also by the Federal Commission for the Protection against Sanitary Risks (COFEPRIS, Mexico) as a regulatory agency.

## Protection of humans and animals

No animals were used in this study. The protocol was designed to comply with the ethical principles of the Helsinki Declaration, Good Clinical Practices, and the applicable Mexican law.

## Privacy rights and informed consent

Informed Consent formats were provided and signed by all participants under Good Clinical Practices and Mexican law. Personal data is confidential, and subjects are not identified personally. Their data was analyzed and duly de-identified.

---

**Supplementary Figure 1.**
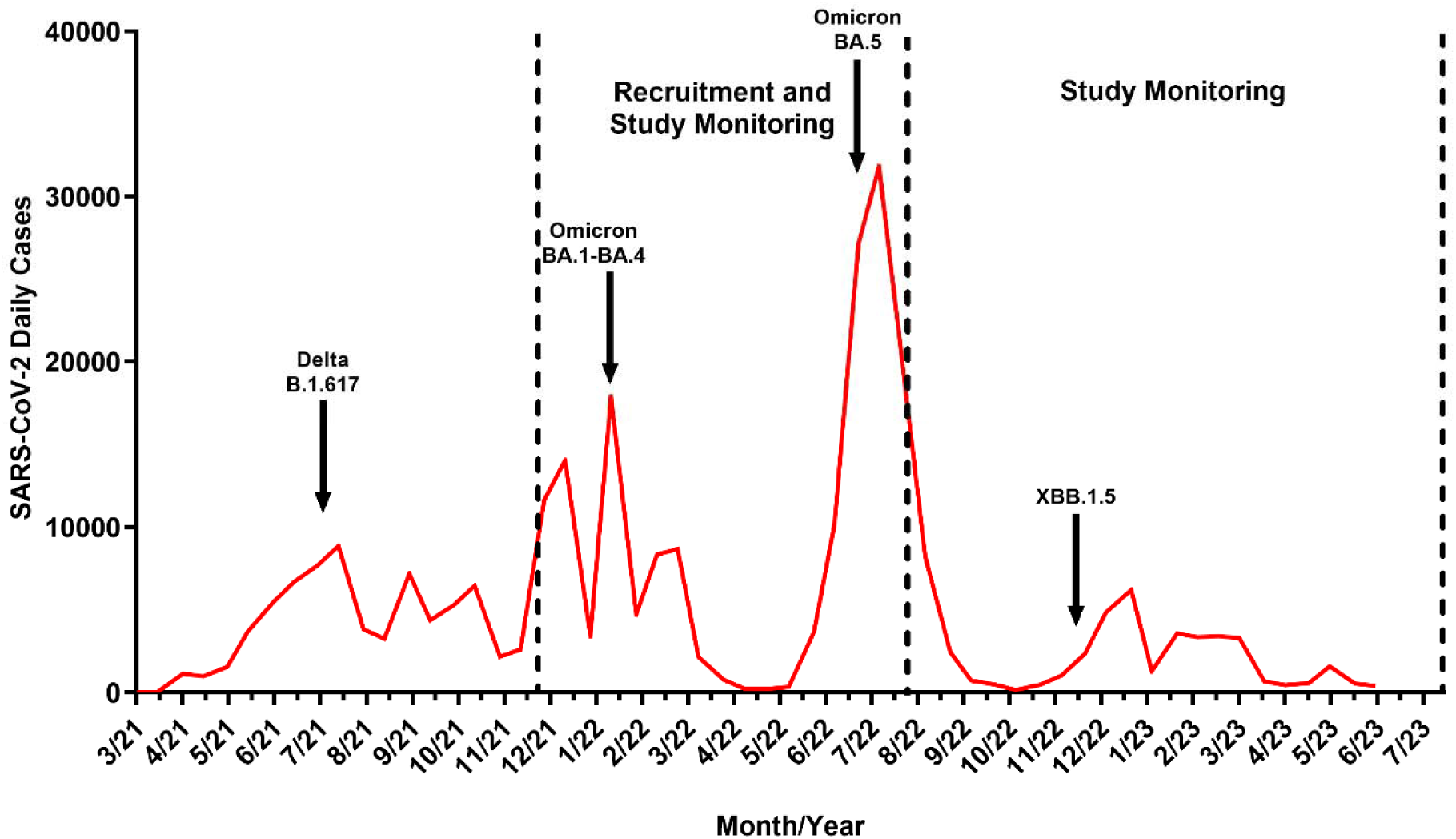
COVID-19 incidence in Mexico during the recruitment and monitoring of the study participants.

**Table 1S.** Proportion of affected subject by adverse events at 7 days post-immunization.

| SOC<br>PT | SEV | IM<br>AVX | % | IM PL | % | IN<br>AVX | % | IN PL | % | IM AZ | % | Total | % | IM<br>AVX-PL<br>p-value | IN<br>AVX-PL<br>p-value | AVX-AZ<br>p-value |
| --- | --- | --- | --- | --- | --- | --- | --- | --- | --- | --- | --- | --- | --- | --- | --- | --- |
| Subjects |  | 41 |  | 40 |  | 40 |  | 37 |  | 71 |  | 229 |  |  |  |  |
| Any AE | Total | 38 | (92.68) | 31 | (77.5) | 36 | (90) | 29 | (78.37) | 52 | (73.23) | 186 | (81.22) | 0.05 | 0.16 | 0.01 |
|  | MI | 38 | (92.68) | 31 | (77.5) | 36 | (90) | 29 | (78.37) | 51 | (71.83) | 185 | (80.78) | 0.05 | 0.16 | 0.008 |
|  | Mo | 13 | (31.7) | 1 | (2.5) | 10 | (25) | 10 | (27.02) | 18 | (25.35) | 52 | (22.7) | 0.0005 | 0.83 | 0.46 |
|  | S | 3 | (7.31) | 0 | (0) | 1 | (2.5) | 0 | (0) | 2 | (2.81) | 6 | (2.62) | 0.08 | 0.33 | 0.26 |
| Pregnancy, puerperium, and perinatal diseases | Total | 0 | (0) | 0 | (0) | 1 | (2.5) | 0 | (0) | 1 | (1.4) | 2 | (0.87) |  | 0.33 | 0.44 |
|  | Mi | 0 | (0) | 0 | (0) | 1 | (2.5) | 0 | (0) | 1 | (1.4) | 2 | (0.87) |  | 0.33 | 0.44 |
|  | Mo | 0 | (0) | 0 | (0) | 0 | (0) | 0 | (0) | 0 | (0) | 0 | (0) |  |  |  |
|  | S | 0 | (0) | 0 | (0) | 0 | (0) | 0 | (0) | 0 | (0) | 0 | (0) |  |  |  |
| Abortion | Mi | 0 | (0) | 0 | (0) | 0 | (0) | 0 | (0) | 1 | (1.4) | 1 | (0.43) |  |  | 0.44 |
| Pregnancy | Mi | 0 | (0) | 0 | (0) | 1 | (2.5) | 0 | (0) | 0 | (0) | 1 | (0.43) |  | 0.33 |  |
| Supplementary examinations | Total | 4 | (9.75) | 0 | (0) | 2 | (5) | 1 | (2.7) | 0 | (0) | 7 | (3.05) | 0.04 | 0.60 | 0.007 |
|  | MI | 4 | (9.75) | 0 | (0) | 2 | (5) | 1 | (2.7) | 0 | (0) | 7 | (3.05) | 0.04 | 0.60 | 0.007 |
|  | Mo | 0 | (0) | 0 | (0) | 0 | (0) | 0 | (0) | 0 | (0) | 0 | (0) |  |  |  |
|  | S | 0 | (0) | 0 | (0) | 0 | (0) | 0 | (0) | 0 | (0) | 0 | (0) |  |  |  |
| Decrease in Glomerular Filtration Rate | Mi | 1 | (2.43) | 0 | (0) | 0 | (0) | 0 | (0) | 0 | (0) | 1 | (0.43) | 0.32 |  | 0.18 |
| Elevation of Aspartate Aminotransferase | Mi | 0 | (0) | 0 | (0) | 1 | (2.5) | 0 | (0) | 0 | (0) | 1 | (0.43) |  | 0.33 |  |
| Elevation of Cystatin C | Mi | 1 | (2.43) | 0 | (0) | 0 | (0) | 0 | (0) | 0 | (0) | 1 | (0.43) | 0.32 |  | 0.18 |
| Elevation of Human Chorionic Gonadotropin | Mi | 1 | (2.43) | 0 | (0) | 0 | (0) | 0 | (0) | 0 | (0) | 1 | (0.43) | 0.32 |  | 0.18 |
| Elevation of Lactate Dehydrogenase | Mi | 1 | (2.43) | 0 | (0) | 0 | (0) | 0 | (0) | 0 | (0) | 1 | (0.43) | 0.32 |  | 0.18 |
| Elevation of Blood Pressure | Mi | 0 | (0) | 0 | (0) | 0 | (0) | 1 | (2.7) | 0 | (0) | 1 | (0.43) |  | 0.29 |  |
| Elevation of Systolic Pressure | Mi | 0 | (0) | 0 | (0) | 1 | (2.5) | 0 | (0) | 0 | (0) | 1 | (0.43) |  | 0.33 |  |
| Elevation of Alanine Aminotransferase | Mi | 1 | (2.43) | 0 | (0) | 0 | (0) | 0 | (0) | 0 | (0) | 1 | (0.43) | 0.32 |  | 0.18 |
| Infections and infestations | Total | 36 | (87.8) | 15 | (37.5) | 29 | (72.5) | 13 | (35.13) | 37 | (52.11) | 130 | (56.76) | 0.0000 | 0.001 | 0.0001 |
|  | Mi | 35 | (85.36) | 15 | (37.5) | 28 | (70) | 13 | (35.13) | 32 | (45.07) | 123 | (53.71) | 0.0000 | 0.002 | 0.0000 |
|  | Mo | 4 | (9.75) | 0 | (0) | 5 | (12.5) | 2 | (5.4) | 7 | (9.85) | 18 | (7.86) | 0.04 | 0.27 | 0.98 |
|  | S | 0 | (0) | 0 | (0) | 0 | (0) | 0 | (0) | 0 | (0) | 0 | (0) |  |  |  |
| Dental Abscess | Mi | 1 | (2.43) | 0 | (0) | 0 | (0) | 0 | (0) | 0 | (0) | 1 | (0.43) | 0.32 |  | 0.18 |
| Bronchitis | Mi | 2 | (4.87) | 0 | (0) | 1 | (2.5) | 0 | (0) | 1 | (1.4) | 4 | (1.74) | 0.15 | 0.33 | 0.27 |
| COVID-19 | Mi | 26 | (63.41) | 9 | (22.5) | 18 | (45) | 11 | (29.72) | 19 | (26.76) | 83 | (36.24) | 0.0002 | 0.16 | 0.0001 |
| COVID-19 | Mo | 1 | (2.43) | 0 | (0) | 3 | (7.5) | 0 | (0) | 1 | (1.4) | 5 | (2.18) | 0.32 | 0.08 | 0.69 |
| Toothache | Mi | 1 | (2.43) | 0 | (0) | 0 | (0) | 1 | (2.7) | 0 | (0) | 2 | (0.87) | 0.32 | 0.29 | 0.18 |
| Gastroenteritis | Mi | 0 | (0) | 0 | (0) | 3 | (7.5) | 1 | (2.7) | 2 | (2.81) | 6 | (2.62) |  | 0.34 | 0.27 |
|  | Mo | 0 | (0) | 0 | (0) | 1 | (2.5) | 1 | (2.7) | 0 | (0) | 2 | (0.87) |  | 0.95 |  |
| Gingivitis | Mi | 0 | (0) | 1 | (2.5) | 0 | (0) | 0 | (0) | 0 | (0) | 1 | (0.43) | 0.30 |  |  |
| Hepatitis A | Mo | 0 | (0) | 0 | (0) | 0 | (0) | 0 | (0) | 1 | (1.4) | 1 | (0.43) |  |  | 0.44 |
| Herpes Zoster | Mi | 0 | (0) | 0 | (0) | 1 | (2.5) | 0 | (0) | 0 | (0) | 1 | (0.43) |  | 0.33 | 0.44 |
| Bacterial Infection of<br>Respiratory Tract | Mi | 0 | (0) | 0 | (0) | 1 | (2.5) | 0 | (0) | 1 | (1.4) | 2 | (0.87) |  | 0.33 | 0.44 |
| Respiratory Tract Infection | Mi | 9 | (21.95) | 0 | (0) | 5 | (12.5) | 0 | (0) | 4 | (5.63) | 18 | (7.86) | 0.001 | 0.02 | 0.009 |
| Upper Respiratory Tract<br>Infection | Mi | 5 | (12.19) | 4 | (10) | 4 | (10) | 0 | (0) | 11 | (15.49) | 24 | (10.48) | 0.75 | 0.04 | 0.63 |
| Upper Respiratory Tract<br>Infection | Mo | 2 | (4.87) | 0 | (0) | 2 | (5) | 1 | (2.7) | 3 | (4.22) | 8 | (3.49) | 0.15 | 0.60 | 0.87 |
| Urinary Tract Infection | Mo | 1 | (2.43) | 0 | (0) | 1 | (2.5) | 0 | (0) | 0 | (0) | 2 | (0.87) | 0.32 | 0.33 | 0.18 |
| Urinary Tract Infection | Mi | 1 | (2.43) | 0 | (0) | 1 | (2.5) | 0 | (0) | 0 | (0) | 2 | (0.87) | 0.32 | 0.33 | 0.18 |
| Dental Infection | Mi | 0 | (0) | 1 | (2.5) | 0 | (0) | 0 | (0) | 0 | (0) | 1 | (0.43) | 0.30 |  |  |
| Dental Infection | Mo | 0 | (0) | 0 | (0) | 1 | (2.5) | 0 | (0) | 0 | (0) | 1 | (0.43) |  | 0.33 |  |
| Vaginal Infection | Mi | 0 | (0) | 0 | (0) | 0 | (0) | 0 | (0) | 1 | (1.4) | 1 | (0.43) |  |  | 0.44 |
| Influenza | Mi | 1 | (2.43) | 0 | (0) | 0 | (0) | 0 | (0) | 0 | (0) | 1 | (0.43) | 0.32 |  | 0.18 |
| Pharyngitis | Mi | 0 | (0) | 0 | (0) | 1 | (2.5) | 0 | (0) | 0 | (0) | 1 | (0.43) |  | 0.33 |  |
| Onychomycosis | Mi | 0 | (0) | 0 | (0) | 0 | (0) | 0 | (0) | 2 | (2.81) | 2 | (0.87) |  |  | 0.27 |
| Osteomyelitis | Mi | 0 | (0) | 0 | (0) | 1 | (2.5) | 0 | (0) | 0 | (0) | 1 | (0.43) |  | 0.33 |  |

| SOC | PT | SEV | IM<br>AVX | % | IM PL | % | IN<br>AVX | % | IN PL | % | IM AZ | % | Total | % | IM<br>AVX-PL<br>p-value | IN<br>AVX-PL<br>p-value | AVX-AZ<br>p-value |
| --- | --- | --- | --- | --- | --- | --- | --- | --- | --- | --- | --- | --- | --- | --- | --- | --- | --- |
|  | Acute Otitis Media | Mi | 0 | (0) | 0 | (0) | 1 | (2.5) | 0 | (0) | 0 | (0) | 1 | (0.43) |  | 0.33 |  |
|  | Acute Otitis Media | Mo | 0 | (0) | 0 | (0) | 0 | (0) | 0 | (0) | 1 | (1.4) | 1 | (0.43) |  |  | 0.44 |
|  | Rhinitis | Mi | 0 | (0) | 0 | (0) | 1 | (2.5) | 0 | (0) | 1 | (1.4) | 2 | (0.87) |  | 0.33 | 0.44 |
|  | Rhinitis | Mo | 0 | (0) | 0 | (0) | 0 | (0) | 0 | (0) | 1 | (1.4) | 1 | (0.43) |  |  | 0.44 |
|  | Post-Acute COVID-19 Syndrome | Mi | 0 | (0) | 0 | (0) | 0 | (0) | 0 | (0) | 1 | (1.4) | 1 | (0.43) |  |  | 0.44 |
|  | American Trypanosomiasis | Mi | 1 | (2.43) | 0 | (0) | 0 | (0) | 0 | (0) | 0 | (0) | 1 | (0.43) | 0.32 |  | 0.18 |
|  | Bacterial Vaginosis | Mi | 1 | (2.43) | 0 | (0) | 0 | (0) | 0 | (0) | 0 | (0) | 1 | (0.43) | 0.32 |  | 0.18 |
| Benign, Malignant, and Unspecified Neoplasms (Cysts and Polyps) |  | Total | 0 | (0) | 0 | (0) | 0 | (0) | 0 | (0) | 1 | (1.4) | 1 | (0.43) |  |  | 0.44 |
|  |  | Mi | 0 | (0) | 0 | (0) | 0 | (0) | 0 | (0) | 1 | (1.4) | 1 | (0.43) |  |  | 0.44 |
|  |  | Mo | 0 | (0) | 0 | (0) | 0 | (0) | 0 | (0) | 0 | (0) | 0 | (0) |  |  |  |
|  |  | S | 0 | (0) | 0 | (0) | 0 | (0) | 0 | (0) | 0 | (0) | 0 | (0) |  |  |  |
|  | Lipoma | Mi | 0 | (0) | 0 | (0) | 0 | (0) | 0 | (0) | 1 | (1.4) | 1 | (0.43) |  |  | 0.44 |
| Traumatic injuries, poisonings, and complications of therapeutic procedures |  | Total | 3 | (7.31) | 4 | (10) | 3 | (7.5) | 1 | (2.7) | 5 | (7.04) | 16 | (6.98) | 0.66 | 0.34 | 0.99 |
|  |  | Mi | 2 | (4.87) | 4 | (10) | 3 | (7.5) | 1 | (2.7) | 4 | (5.63) | 14 | (6.11) | 0.37 | 0.34 | 0.86 |
|  |  | Mo | 1 | (2.43) | 0 | (0) | 0 | (0) | 0 | (0) | 1 | (1.4) | 2 | (0.87) | 0.32 |  | 0.69 |
|  |  | S | 0 | (0) | 0 | (0) | 0 | (0) | 0 | (0) | 0 | (0) | 0 | (0) |  |  |  |
|  | Epicondylitis | Mo | 0 | (0) | 0 | (0) | 0 | (0) | 0 | (0) | 1 | (1.4) | 1 | (0.43) |  |  | 0.44 |
|  | Dental Fracture | Mi | 1 | (2.43) | 0 | (0) | 0 | (0) | 0 | (0) | 1 | (1.4) | 2 | (0.87) | 0.32 |  | 0.69 |
|  | Sunstroke | Mi | 0 | (0) | 0 | (0) | 1 | (2.5) | 0 | (0) | 0 | (0) | 1 | (0.43) |  | 0.33 |  |
|  | Injury | Mi | 2 | (4.87) | 1 | (2.5) | 2 | (5) | 1 | (2.7) | 0 | (0) | 6 | (2.62) | 0.57 | 0.60 | 0.06 |
|  | Traumatic Skin Injury | Mi | 0 | (0) | 0 | (0) | 0 | (0) | 0 | (0) | 1 | (1.4) | 1 | (0.43) |  |  | 0.44 |
|  | Facial Traumatic Injury | Mi | 0 | (0) | 1 | (2.5) | 0 | (0) | 0 | (0) | 0 | (0) | 1 | (0.43) | 0.30 |  |  |
|  | Animal Bite | Mo | 1 | (2.43) | 0 | (0) | 0 | (0) | 0 | (0) | 0 | (0) | 1 | (0.43) | 0.32 |  | 0.18 |
|  | Chemical Burn | Mi | 0 | (0) | 0 | (0) | 0 | (0) | 0 | (0) | 1 | (1.4) | 1 | (0.43) |  |  | 0.44 |
|  | Chemical Burn | Mi | 0 | (0) | 1 | (2.5) | 0 | (0) | 0 | (0) | 0 | (0) | 1 | (0.43) | 0.30 |  |  |
|  | Thermal Burn | Mi | 0 | (0) | 1 | (2.5) | 0 | (0) | 0 | (0) | 0 | (0) | 1 | (0.43) | 0.30 |  |  |

| SOC PT | SEV | IM AVX | % | IM PL | % | IN AVX | % | IN PL | % | IM AZ | % | Total | % | IM AVX-PL p-value | IN AVX-PL p-value | AVX-AZ p-value |
| --- | --- | --- | --- | --- | --- | --- | --- | --- | --- | --- | --- | --- | --- | --- | --- | --- |
| Injection-Related Reaction | Mi | 0 | (0) | 0 | (0) | 1 | (2.5) | 0 | (0) | 1 | (1.4) | 2 | (0.87) |  | 0.33 | 0.44 |
| Medical and Surgical Procedures | Total | 1 | (2.43) | 0 | (0) | 3 | (7.5) | 0 | (0) | 3 | (4.22) | 7 | (3.05) | 0.32 | 0.08 | 0.62 |
|  | Mi | 1 | (2.43) | 0 | (0) | 3 | (7.5) | 0 | (0) | 3 | (4.22) | 7 | (3.05) | 0.32 | 0.08 | 0.62 |
|  | Mo | 0 | (0) | 0 | (0) | 0 | (0) | 0 | (0) | 0 | (0) | 0 | (0) |  |  |  |
|  | S | 0 | (0) | 0 | (0) | 0 | (0) | 0 | (0) | 0 | (0) | 0 | (0) |  |  |  |
| Dental Care | Mi | 0 | (0) | 0 | (0) | 0 | (0) | 0 | (0) | 1 | (1.4) | 1 | (0.43) |  |  | 0.44 |
| Periodontal Surgery | Mi | 1 | (2.43) | 0 | (0) | 0 | (0) | 0 | (0) | 0 | (0) | 1 | (0.43) | 0.32 |  | 0.18 |
| Tooth Extraction | Mi | 0 | (0) | 0 | (0) | 2 | (5) | 0 | (0) | 1 | (1.4) | 3 | (1.31) |  | 0.16 | 0.44 |
| Hysterectomy | Mi | 0 | (0) | 0 | (0) | 1 | (2.5) | 0 | (0) | 0 | (0) | 1 | (0.43) |  | 0.33 |  |
| Removal of Intrauterine Contraceptive Device | Mi | 0 | (0) | 0 | (0) | 0 | (0) | 0 | (0) | 1 | (1.4) | 1 | (0.43) |  |  | 0.44 |
| Cardiac Disorders | Total | 0 | (0) | 0 | (0) | 1 | (2.5) | 1 | (2.7) | 2 | (2.81) | 4 | (1.74) |  | 0.95 | 0.27 |
|  | Mi | 0 | (0) | 0 | (0) | 1 | (2.5) | 1 | (2.7) | 0 | (0) | 2 | (0.87) |  | 0.95 |  |
|  | Mo | 0 | (0) | 0 | (0) | 0 | (0) | 0 | (0) | 2 | (2.81) | 2 | (0.87) |  |  | 0.27 |
|  | S | 0 | (0) | 0 | (0) | 0 | (0) | 0 | (0) | 0 | (0) | 0 | (0) |  |  |  |
| Arrhythmia | Mo | 0 | (0) | 0 | (0) | 0 | (0) | 0 | (0) | 1 | (1.4) | 1 | (0.43) |  |  | 0.44 |
| Chest Pain | Mo | 0 | (0) | 0 | (0) | 0 | (0) | 0 | (0) | 1 | (1.4) | 1 | (0.43) |  |  | 0.44 |
| Palpitations | Mi | 0 | (0) | 0 | (0) | 1 | (2.5) | 0 | (0) | 0 | (0) | 1 | (0.43) |  | 0.33 |  |
| Tachycardia | Mi | 0 | (0) | 0 | (0) | 0 | (0) | 1 | (2.7) | 0 | (0) | 1 | (0.43) |  | 0.29 |  |
| Skin and Subcutaneous Tissue Disorders | Total | 0 | (0) | 5 | (12.5) | 3 | (7.5) | 0 | (0) | 8 | (11.26) | 16 | (6.98) | 0.01 | 0.08 | 0.03 |
|  | Mi | 0 | (0) | 5 | (12.5) | 3 | (7.5) | 0 | (0) | 7 | (9.85) | 15 | (6.55) | 0.01 | 0.08 | 0.04 |
|  | Mo | 0 | (0) | 0 | (0) | 0 | (0) | 0 | (0) | 1 | (1.4) | 1 | (0.43) |  |  | 0.44 |
|  | S | 0 | (0) | 0 | (0) | 0 | (0) | 0 | (0) | 0 | (0) | 0 | (0) |  |  |  |
| Acne | Mi | 0 | (0) | 0 | (0) | 0 | (0) | 0 | (0) | 1 | (1.4) | 1 | (0.43) |  |  | 0.44 |
| Dermatitis | Mi | 0 | (0) | 2 | (5) | 2 | (5) | 0 | (0) | 2 | (2.81) | 6 | (2.62) | 0.14 | 0.16 | 0.27 |
| Dermatitis | Mo | 0 | (0) | 0 | (0) | 0 | (0) | 0 | (0) | 1 | (1.4) | 1 | (0.43) |  |  | 0.44 |
| Bruising | Mi | 0 | (0) | 1 | (2.5) | 1 | (2.5) | 0 | (0) | 0 | (0) | 2 | (0.87) | 0.30 | 0.33 |  |
| Erythema | Mi | 0 | (0) | 0 | (0) | 1 | (2.5) | 0 | (0) | 0 | (0) | 1 | (0.43) |  | 0.33 |  |

| SOC | PT | SEV | IM<br>AVX | % | IM PL | % | IN<br>AVX | % | IN PL | % | IM AZ | % | Total | % | IM<br>AVX-PL<br>p-value | IN<br>AVX-PL<br>p-value | AVX-AZ<br>p-value |
| --- | --- | --- | --- | --- | --- | --- | --- | --- | --- | --- | --- | --- | --- | --- | --- | --- | --- |
|  | Granuloma | Mi | 0 | (0) | 0 | (0) | 0 | (0) | 0 | (0) | 1 | (1.4) | 1 | (0.43) |  |  | 0.44 |
|  | Hyperhidrosis | Mi | 0 | (0) | 0 | (0) | 1 | (2.5) | 0 | (0) | 0 | (0) | 1 | (0.43) |  | 0.33 |  |
|  | Petechiae | Mi | 0 | (0) | 1 | (2.5) | 0 | (0) | 0 | (0) | 0 | (0) | 1 | (0.43) | 0.30 |  |  |
|  | Pruritus | Mi | 0 | (0) | 1 | (2.5) | 1 | (2.5) | 0 | (0) | 2 | (2.81) | 4 | (1.74) | 0.30 | 0.33 | 0.27 |
| Disorders of the blood and lymphatic system |  | Total | 2 | (4.87) | 3 | (7.5) | 3 | (7.5) | 3 | (8.1) | 0 | (0) | 11 | (4.8) | 0.62 | 0.92 | 0.06 |
|  |  | Mi | 2 | (4.87) | 3 | (7.5) | 3 | (7.5) | 2 | (5.4) | 0 | (0) | 10 | (4.36) | 0.62 | 0.70 | 0.06 |
|  |  | Mo | 0 | (0) | 0 | (0) | 0 | (0) | 1 | (2.7) | 0 | (0) | 1 | (0.43) |  | 0.29 |  |
|  |  | S | 0 | (0) | 0 | (0) | 0 | (0) | 0 | (0) | 0 | (0) | 0 | (0) |  |  |  |
|  | Hypochromic anemia | Mi | 1 | (2.43) | 0 | (0) | 0 | (0) | 0 | (0) | 0 | (0) | 1 | (0.43) | 0.32 |  | 0.18 |
|  | Basophilia | Mi | 0 | (0) | 0 | (0) | 0 | (0) | 1 | (2.7) | 0 | (0) | 1 | (0.43) |  | 0.29 |  |
|  | Coagulopathy | Mi | 0 | (0) | 1 | (2.5) | 0 | (0) | 0 | (0) | 0 | (0) | 1 | (0.43) | 0.30 |  |  |
|  | Eosinophilia | Mi | 0 | (0) | 0 | (0) | 0 | (0) | 1 | (2.7) | 0 | (0) | 1 | (0.43) |  | 0.29 |  |
|  | Leucopenia | Mi | 0 | (0) | 0 | (0) | 1 | (2.5) | 0 | (0) | 0 | (0) | 1 | (0.43) |  | 0.33 |  |
|  | Lymphopenia | Mi | 0 | (0) | 0 | (0) | 1 | (2.5) | 0 | (0) | 0 | (0) | 1 | (0.43) |  | 0.33 |  |
|  | Neutrophilia | Mi | 0 | (0) | 1 | (2.5) | 0 | (0) | 0 | (0) | 0 | (0) | 1 | (0.43) | 0.30 |  |  |
|  | Neutropenia | Mi | 0 | (0) | 0 | (0) | 1 | (2.5) | 1 | (2.7) | 0 | (0) | 2 | (0.87) |  | 0.95 |  |
|  | Thrombocytosis | Mi | 2 | (4.87) | 1 | (2.5) | 0 | (0) | 1 | (2.7) | 0 | (0) | 4 | (1.74) | 0.57 | 0.29 | 0.06 |
|  | Thrombocytopenia | Mo | 0 | (0) | 0 | (0) | 0 | (0) | 1 | (2.7) | 0 | (0) | 1 | (0.43) |  | 0.29 |  |
| Reproductive and breast disorders |  | Total | 5 | (12.19) | 2 | (5) | 1 | (2.5) | 3 | (8.1) | 7 | (9.85) | 18 | (7.86) | 0.24 | 0.26 | 0.70 |
|  |  | Mi | 3 | (7.31) | 2 | (5) | 1 | (2.5) | 1 | (2.7) | 6 | (8.45) | 13 | (5.67) | 0.66 | 0.95 | 0.83 |
|  |  | Mo | 1 | (2.43) | 0 | (0) | 0 | (0) | 2 | (5.4) | 2 | (2.81) | 5 | (2.18) | 0.32 | 0.13 | 0.90 |
|  |  | S | 1 | (2.43) | 0 | (0) | 0 | (0) | 0 | (0) | 0 | (0) | 1 | (0.43) | 0.32 |  | 0.18 |
|  | Dysmenorrhea | Mi | 0 | (0) | 1 | (2.5) | 0 | (0) | 1 | (2.7) | 1 | (1.4) | 3 | (1.31) | 0.30 | 0.29 | 0.44 |
|  | Dysmenorrhea | Mo | 0 | (0) | 0 | (0) | 0 | (0) | 1 | (2.7) | 0 | (0) | 1 | (0.43) |  | 0.29 |  |
|  | Vaginal bleeding | Mi | 1 | (2.43) | 0 | (0) | 0 | (0) | 0 | (0) | 0 | (0) | 1 | (0.43) | 0.32 |  | 0.18 |
|  | Polycystic ovaries | Mi | 1 | (2.43) | 0 | (0) | 0 | (0) | 0 | (0) | 1 | (1.4) | 2 | (0.87) | 0.32 |  | 0.69 |
|  | Breast cyst | Mi | 0 | (0) | 0 | (0) | 1 | (2.5) | 0 | (0) | 0 | (0) | 1 | (0.43) |  | 0.33 |  |
|  | Prostatitis | Mo | 1 | (2.43) | 0 | (0) | 0 | (0) | 0 | (0) | 0 | (0) | 1 | (0.43) | 0.32 |  | 0.18 |
|  | Delayed menstruation | Mi | 0 | (0) | 0 | (0) | 0 | (0) | 0 | (0) | 1 | (1.4) | 1 | (0.43) |  |  | 0.44 |
|  | Heavy menstrual bleeding | Mi | 1 | (2.43) | 0 | (0) | 0 | (0) | 0 | (0) | 0 | (0) | 1 | (0.43) | 0.32 |  | 0.18 |
|  | Menstrual disorders | Mi | 1 | (2.43) | 1 | (2.5) | 0 | (0) | 0 | (0) | 3 | (4.22) | 5 | (2.18) | 0.98 |  | 0.62 |
|  | Menstrual disorders | Mo | 0 | (0) | 0 | (0) | 0 | (0) | 1 | (2.7) | 1 | (1.4) | 2 | (0.87) |  | 0.29 | 0.44 |
|  | Ovarian cyst | S | 1 | (2.43) | 0 | (0) | 0 | (0) | 0 | (0) | 0 | (0) | 1 | (0.43) | 0.32 |  | 0.18 |
|  | Varicocele | Mo | 0 | (0) | 0 | (0) | 0 | (0) | 0 | (0) | 1 | (1.4) | 1 | (0.43) |  |  | 0.44 |
| Metabolic and Nutritional Disorders |  | Total | 10 | (24.39) | 7 | (17.5) | 6 | (15) | 6 | (16.21) | 1 | (1.4) | 30 | (13.1) | 0.44 | 0.88 | 0.001 |
|  |  | Mi | 10 | (24.39) | 7 | (17.5) | 6 | (15) | 6 | (16.21) | 1 | (1.4) | 30 | (13.1) | 0.44 | 0.88 | 0.001 |
|  |  | M | 0 | (0) | 0 | (0) | 0 | (0) | 0 | (0) | 0 | (0) | 0 | (0) |  |  |  |
|  |  | S | 0 | (0) | 0 | (0) | 0 | (0) | 0 | (0) | 0 | (0) | 0 | (0) |  |  |  |
|  | Vitamin B12 Deficiency | Mi | 1 | (2.43) | 0 | (0) | 0 | (0) | 0 | (0) | 0 | (0) | 1 | (0.43) | 0.32 |  | 0.18 |
|  | Vitamin D Deficiency | Mi | 0 | (0) | 0 | (0) | 1 | (2.5) | 0 | (0) | 0 | (0) | 1 | (0.43) |  | 0.33 |  |
|  | Dyslipidemia | Mi | 2 | (4.87) | 0 | (0) | 2 | (5) | 2 | (5.4) | 0 | (0) | 6 | (2.62) | 0.15 | 0.93 | 0.06 |
|  | Hypercholesterolemia | Mi | 2 | (4.87) | 2 | (5) | 3 | (7.5) | 1 | (2.7) | 0 | (0) | 8 | (3.49) | 0.97 | 0.34 | 0.06 |
|  | Hyperglycemia | Mi | 1 | (2.43) | 0 | (0) | 0 | (0) | 0 | (0) | 0 | (0) | 1 | (0.43) | 0.32 |  | 0.18 |
|  | Hypertriglyceridemia | Mi | 3 | (7.31) | 4 | (10) | 1 | (2.5) | 3 | (8.1) | 0 | (0) | 11 | (4.8) | 0.66 | 0.26 | 0.02 |
|  | Obesity | Mi | 0 | (0) | 1 | (2.5) | 1 | (2.5) | 0 | (0) | 0 | (0) | 2 | (0.87) | 0.30 | 0.33 |  |
|  | Insulin Resistance | Mi | 2 | (4.87) | 0 | (0) | 0 | (0) | 0 | (0) | 0 | (0) | 2 | (0.87) | 0.15 |  | 0.06 |
|  | Metabolic Syndrome | Mi | 1 | (2.43) | 0 | (0) | 0 | (0) | 0 | (0) | 0 | (0) | 1 | (0.43) | 0.32 |  | 0.18 |
|  | Overweight | Mi | 0 | (0) | 0 | (0) | 0 | (0) | 0 | (0) | 1 | (1.4) | 1 | (0.43) |  |  | 0.44 |
| Disorders of the ear and labyrinth |  | Total | 1 | (2.43) | 0 | (0) | 2 | (5) | 0 | (0) | 0 | (0) | 3 | (1.31) | 0.32 | 0.16 | 0.18 |
|  |  | Mi | 1 | (2.43) | 0 | (0) | 2 | (5) | 0 | (0) | 0 | (0) | 3 | (1.31) | 0.32 | 0.16 | 0.18 |
|  |  | Mo | 0 | (0) | 0 | (0) | 0 | (0) | 0 | (0) | 0 | (0) | 0 | (0) |  |  |  |
|  |  | S | 0 | (0) | 0 | (0) | 0 | (0) | 0 | (0) | 0 | (0) | 0 | (0) |  |  |  |
|  | Ear pain | Mi | 0 | (0) | 0 | (0) | 1 | (2.5) | 0 | (0) | 0 | (0) | 1 | (0.43) |  | 0.33 |  |
|  | Ear disorder | Mi | 0 | (0) | 0 | (0) | 1 | (2.5) | 0 | (0) | 0 | (0) | 1 | (0.43) |  | 0.33 |  |
|  | Vertigo | Mi | 1 | (2.43) | 0 | (0) | 0 | (0) | 0 | (0) | 0 | (0) | 1 | (0.43) | 0.32 |  | 0.18 |

| SOC PT | SEV | IM AVX | % | IM PL | % | IN AVX | % | IN PL | % | IM AZ | % | Total | % | IM AVX-PL p-value | IN AVX-PL p-value | AVX-AZ p-value |
| --- | --- | --- | --- | --- | --- | --- | --- | --- | --- | --- | --- | --- | --- | --- | --- | --- |
| Disorders of the immune system | Total | 1 | (2.43) | 0 | (0) | 0 | (0) | 0 | (0) | 0 | (0) | 1 | (0.43) | 0.32 |  | 0.18 |
|  | Mi | 1 | (2.43) | 0 | (0) | 0 | (0) | 0 | (0) | 0 | (0) | 1 | (0.43) | 0.32 |  | 0.18 |
|  | Mo | 0 | (0) | 0 | (0) | 0 | (0) | 0 | (0) | 0 | (0) | 0 | (0) |  |  |  |
|  | S | 0 | (0) | 0 | (0) | 0 | (0) | 0 | (0) | 0 | (0) | 0 | (0) |  |  |  |
| Hypersensitivity | Mi | 1 | (2.43) | 0 | (0) | 0 | (0) | 0 | (0) | 0 | (0) | 1 | (0.43) | 0.32 |  | 0.18 |
| Disorders of the nervous system | Total | 17 | (41.46) | 16 | (40) | 16 | (40) | 16 | (43.24) | 17 | (23.94) | 82 | (35.8) | 0.89 | 0.77 | 0.05 |
|  | Mi | 15 | (36.58) | 16 | (40) | 15 | (37.5) | 14 | (37.83) | 14 | (19.71) | 74 | (32.31) | 0.75 | 0.97 | 0.05 |
|  | Mo | 6 | (14.63) | 1 | (2.5) | 2 | (5) | 4 | (10.81) | 3 | (4.22) | 16 | (6.98) | 0.05 | 0.34 | 0.05 |
|  | S | 0 | (0) | 0 | (0) | 0 | (0) | 0 | (0) | 1 | (1.4) | 1 | (0.43) |  |  | 0.44 |
| Ageusia | Mi | 0 | (0) | 0 | (0) | 1 | (2.5) | 0 | (0) | 0 | (0) | 1 | (0.43) |  | 0.33 |  |
| Anosmia | Mo | 1 | (2.43) | 0 | (0) | 0 | (0) | 0 | (0) | 0 | (0) | 1 | (0.43) | 0.32 |  | 0.18 |
| Headache | Mi | 14 | (34.14) | 14 | (35) | 15 | (37.5) | 14 | (37.83) | 12 | (16.9) | 69 | (30.13) | 0.93 | 0.97 | 0.03 |
| Headache | Mo | 5 | (12.19) | 1 | (2.5) | 1 | (2.5) | 4 | (10.81) | 3 | (4.22) | 14 | (6.11) | 0.09 | 0.13 | 0.11 |
| Bruise | Mi | 0 | (0) | 1 | (2.5) | 0 | (0) | 0 | (0) | 0 | (0) | 1 | (0.43) | 0.30 |  |  |
| Memory impairment | Mo | 1 | (2.43) | 0 | (0) | 0 | (0) | 0 | (0) | 0 | (0) | 1 | (0.43) | 0.32 |  | 0.18 |
| Dizziness | Mi | 0 | (0) | 3 | (7.5) | 0 | (0) | 1 | (2.7) | 1 | (1.4) | 5 | (2.18) | 0.07 | 0.29 | 0.44 |
| Migraine | Mo | 0 | (0) | 0 | (0) | 1 | (2.5) | 0 | (0) | 0 | (0) | 1 | (0.43) |  | 0.33 |  |
| Paresthesia | Mi | 2 | (4.87) | 0 | (0) | 2 | (5) | 1 | (2.7) | 0 | (0) | 5 | (2.18) | 0.15 | 0.60 | 0.06 |
| Loss of muscle strength | Mi | 0 | (0) | 1 | (2.5) | 0 | (0) | 1 | (2.7) | 0 | (0) | 2 | (0.87) | 0.30 | 0.29 |  |
| Guillain-Barré syndrome | S | 0 | (0) | 0 | (0) | 0 | (0) | 0 | (0) | 1 | (1.4) | 1 | (0.43) |  |  | 0.44 |
| Drowsiness | Mi | 0 | (0) | 0 | (0) | 0 | (0) | 0 | (0) | 1 | (1.4) | 1 | (0.43) |  |  | 0.44 |
| Vertigo | Mi | 0 | (0) | 1 | (2.5) | 0 | (0) | 0 | (0) | 0 | (0) | 1 | (0.43) | 0.30 |  |  |
| Endocrine disorders | Total | 0 | (0) | 0 | (0) | 1 | (2.5) | 0 | (0) | 1 | (1.4) | 2 | (0.87) |  | 0.33 | 0.44 |
|  | Mi | 0 | (0) | 0 | (0) | 1 | (2.5) | 0 | (0) | 1 | (1.4) | 2 | (0.87) |  | 0.33 | 0.44 |
|  | Mo | 0 | (0) | 0 | (0) | 0 | (0) | 0 | (0) | 0 | (0) | 0 | (0) |  |  |  |
|  | S | 0 | (0) | 0 | (0) | 0 | (0) | 0 | (0) | 0 | (0) | 0 | (0) |  |  |  |
| Hyperprolactinemia | Mi | 0 | (0) | 0 | (0) | 0 | (0) | 0 | (0) | 1 | (1.4) | 1 | (0.43) |  |  | 0.44 |
| Hypothyroidism | Mi | 0 | (0) | 0 | (0) | 1 | (2.5) | 0 | (0) | 0 | (0) | 1 | (0.43) |  | 0.33 |  |

| SOC<br>PT | SEV | IM<br>AVX | % | IM PL | % | IN<br>AVX | % | IN PL | % | IM AZ | % | Total | % | IM<br>AVX-PL<br>p-value | IN<br>AVX-PL<br>p-value | AVX-AZ<br>p-value |
| --- | --- | --- | --- | --- | --- | --- | --- | --- | --- | --- | --- | --- | --- | --- | --- | --- |
| Gastrointestinal disorders | Total | 13 | (31.7) | 7 | (17.5) | 16 | (40) | 7 | (18.91) | 15 | (21.12) | 58 | (25.32) | 0.13 | 0.04 | 0.21 |
|  | Mi | 11 | (26.82) | 7 | (17.5) | 15 | (37.5) | 7 | (18.91) | 12 | (16.9) | 52 | (22.7) | 0.31 | 0.07 | 0.21 |
|  | Mo | 3 | (7.31) | 0 | (0) | 1 | (2.5) | 0 | (0) | 4 | (5.63) | 8 | (3.49) | 0.08 | 0.33 | 0.72 |
|  | S | 0 | (0) | 0 | (0) | 1 | (2.5) | 0 | (0) | 0 | (0) | 1 | (0.43) |  | 0.33 |  |
| Dental caries | Mi | 0 | (0) | 0 | (0) | 1 | (2.5) | 0 | (0) | 0 | (0) | 1 | (0.43) |  | 0.33 |  |
| Colitis | Mi | 1 | (2.43) | 0 | (0) | 0 | (0) | 0 | (0) | 1 | (1.4) | 2 | (0.87) | 0.32 |  | 0.69 |
| Colitis | Mo | 0 | (0) | 0 | (0) | 0 | (0) | 0 | (0) | 1 | (1.4) | 1 | (0.43) |  |  | 0.44 |
| Diarrhea | Mi | 2 | (4.87) | 3 | (7.5) | 3 | (7.5) | 1 | (2.7) | 2 | (2.81) | 11 | (4.8) | 0.62 | 0.34 | 0.57 |
| Diarrhea | Mo | 2 | (4.87) | 0 | (0) | 0 | (0) | 0 | (0) | 1 | (1.4) | 3 | (1.31) | 0.15 |  | 0.27 |
| Dyspepsia | Mi | 0 | (0) | 1 | (2.5) | 1 | (2.5) | 1 | (2.7) | 1 | (1.4) | 4 | (1.74) | 0.30 | 0.95 | 0.44 |
| Dyspepsia | Mo | 0 | (0) | 0 | (0) | 0 | (0) | 0 | (0) | 1 | (1.4) | 1 | (0.43) |  |  | 0.44 |
| Abdominal distension | Mi | 0 | (0) | 0 | (0) | 0 | (0) | 2 | (5.4) | 0 | (0) | 2 | (0.87) |  | 0.13 |  |
| Abdominal pain | Mi | 0 | (0) | 1 | (2.5) | 1 | (2.5) | 0 | (0) | 0 | (0) | 2 | (0.87) | 0.30 | 0.33 |  |
| Lower abdominal pain | Mi | 0 | (0) | 0 | (0) | 0 | (0) | 0 | (0) | 1 | (1.4) | 1 | (0.43) |  |  | 0.44 |
| Periodontal disease | Mi | 1 | (2.43) | 0 | (0) | 0 | (0) | 0 | (0) | 0 | (0) | 1 | (0.43) | 0.32 |  | 0.18 |
| Gastroesophageal reflux disease | Mi | 0 | (0) | 0 | (0) | 0 | (0) | 0 | (0) | 1 | (1.4) | 1 | (0.43) |  |  | 0.44 |
| Gastroesophageal reflux disease | Mo | 0 | (0) | 0 | (0) | 0 | (0) | 0 | (0) | 1 | (1.4) | 1 | (0.43) |  |  | 0.44 |
| Gastritis | Mi | 1 | (2.43) | 0 | (0) | 1 | (2.5) | 0 | (0) | 0 | (0) | 2 | (0.87) | 0.32 | 0.33 | 0.18 |
| Glossitis | Mi | 0 | (0) | 0 | (0) | 0 | (0) | 0 | (0) | 1 | (1.4) | 1 | (0.43) |  |  | 0.44 |
| Inguinal hernia | Mi | 0 | (0) | 0 | (0) | 0 | (0) | 0 | (0) | 1 | (1.4) | 1 | (0.43) |  |  | 0.44 |
| Food poisoning | Mi | 0 | (0) | 0 | (0) | 0 | (0) | 0 | (0) | 1 | (1.4) | 1 | (0.43) |  |  | 0.44 |
| Nausea | Mi | 1 | (2.43) | 1 | (2.5) | 1 | (2.5) | 0 | (0) | 1 | (1.4) | 4 | (1.74) | 0.98 | 0.33 | 0.69 |
| Odynophagia | Mi | 4 | (9.75) | 1 | (2.5) | 7 | (17.5) | 2 | (5.4) | 3 | (4.22) | 17 | (7.42) | 0.17 | 0.09 | 0.24 |
| Odynophagia | Mo | 1 | (2.43) | 0 | (0) | 1 | (2.5) | 0 | (0) | 0 | (0) | 2 | (0.87) | 0.32 | 0.33 | 0.18 |
| Acute pancreatitis | S | 0 | (0) | 0 | (0) | 1 | (2.5) | 0 | (0) | 0 | (0) | 1 | (0.43) |  | 0.33 |  |
| Rectal polyp | Mi | 1 | (2.43) | 0 | (0) | 0 | (0) | 0 | (0) | 0 | (0) | 1 | (0.43) | 0.32 |  | 0.18 |
| Irritable bowel syndrome | Mi | 0 | (0) | 0 | (0) | 0 | (0) | 1 | (2.7) | 1 | (1.4) | 2 | (0.87) |  | 0.29 | 0.44 |

| SOC | PT | SEV | IM<br>AVX | % | IM PL | % | IN<br>AVX | % | IN PL | % | IM AZ | % | Total | % | IM<br>AVX-PL<br>p-value | IN<br>AVX-PL<br>p-value | AVX-AZ<br>p-value |
| --- | --- | --- | --- | --- | --- | --- | --- | --- | --- | --- | --- | --- | --- | --- | --- | --- | --- |
|  | Vomiting | Mi | 1 | (2.43) | 1 | (2.5) | 0 | (0) | 0 | (0) | 0 | (0) | 2 | (0.87) | 0.98 |  | 0.18 |
|  | Aphthous ulcer | Mi | 0 | (0) | 0 | (0) | 1 | (2.5) | 0 | (0) | 0 | (0) | 1 | (0.43) |  | 0.33 |  |
| General disorders and administration site conditions |  | Total | 25 | (60.97) | 8 | (20) | 14 | (35) | 8 | (21.62) | 18 | (25.35) | 73 | (31.87) | 0.0002 | 0.19 | 0.0002 |
|  |  | Mi | 24 | (58.53) | 8 | (20) | 14 | (35) | 7 | (18.91) | 17 | (23.94) | 70 | (30.56) | 0.0004 | 0.11 | 0.0003 |
|  |  | Mo | 2 | (4.87) | 0 | (0) | 0 | (0) | 1 | (2.7) | 1 | (1.4) | 4 | (1.74) | 0.15 | 0.29 | 0.27 |
|  |  | S | 0 | (0) | 0 | (0) | 0 | (0) | 0 | (0) | 0 | (0) | 0 | (0) |  |  |  |
|  | Asthenia | Mi | 1 | (2.43) | 2 | (5) | 3 | (7.5) | 4 | (10.81) | 3 | (4.22) | 13 | (5.67) | 0.54 | 0.61 | 0.62 |
|  | Axillary Pain | Mo | 1 | (2.43) | 0 | (0) | 0 | (0) | 0 | (0) | 0 | (0) | 1 | (0.43) | 0.32 |  | 0.18 |
|  | Injection Site Pain | Mi | 1 | (2.43) | 0 | (0) | 0 | (0) | 0 | (0) | 0 | (0) | 1 | (0.43) | 0.32 |  | 0.18 |
|  | Chest Pain | Mi | 16 | (39.02) | 5 | (12.5) | 0 | (0) | 0 | (0) | 10 | (14.08) | 31 | (13.53) | 0.006 |  | 0.002 |
|  | Flu-like Illness | Mo | 1 | (2.43) | 0 | (0) | 0 | (0) | 0 | (0) | 0 | (0) | 1 | (0.43) | 0.32 |  | 0.18 |
|  | Chill | Mo | 0 | (0) | 0 | (0) | 0 | (0) | 0 | (0) | 1 | (1.4) | 1 | (0.43) |  |  | 0.44 |
|  | Fatigue | Mi | 0 | (0) | 0 | (0) | 0 | (0) | 0 | (0) | 1 | (1.4) | 1 | (0.43) |  |  | 0.44 |
|  | General Discomfort | Mi | 1 | (2.43) | 0 | (0) | 0 | (0) | 0 | (0) | 0 | (0) | 1 | (0.43) | 0.32 |  | 0.18 |
|  | Chest Discomfort | Mi | 2 | (4.87) | 1 | (2.5) | 3 | (7.5) | 0 | (0) | 1 | (1.4) | 7 | (3.05) | 0.57 | 0.08 | 0.27 |
|  | Fever | Mi | 5 | (12.19) | 0 | (0) | 4 | (10) | 2 | (5.4) | 1 | (1.4) | 12 | (5.24) | 0.02 | 0.45 | 0.01 |
|  | Dry Skin | Mi | 3 | (7.31) | 0 | (0) | 4 | (10) | 1 | (2.7) | 4 | (5.63) | 12 | (5.24) | 0.08 | 0.19 | 0.72 |
|  | Asthenia | Mi | 0 | (0) | 0 | (0) | 1 | (2.5) | 0 | (0) | 0 | (0) | 1 | (0.43) |  | 0.33 |  |
|  | Axillary Pain | Mi | 5 | (12.19) | 1 | (2.5) | 2 | (5) | 2 | (5.4) | 6 | (8.45) | 16 | (6.98) | 0.09 | 0.93 | 0.52 |
|  | Injection Site Pain | Mo | 1 | (2.43) | 0 | (0) | 0 | (0) | 1 | (2.7) | 0 | (0) | 2 | (0.87) | 0.32 | 0.29 | 0.18 |
|  | Chest Pain | Mi | 0 | (0) | 0 | (0) | 0 | (0) | 1 | (2.7) | 0 | (0) | 1 | (0.43) |  | 0.29 |  |
| Musculoskeletal and connective tissue disorders |  | Total | 11 | (26.82) | 8 | (20) | 8 | (20) | 6 | (16.21) | 14 | (19.71) | 47 | (20.52) | 0.45 | 0.66 | 0.38 |
|  |  | Mi | 9 | (21.95) | 8 | (20) | 6 | (15) | 6 | (16.21) | 14 | (19.71) | 43 | (18.77) | 0.82 | 0.88 | 0.77 |
|  |  | Mo | 2 | (4.87) | 0 | (0) | 2 | (5) | 0 | (0) | 1 | (1.4) | 5 | (2.18) | 0.15 | 0.16 | 0.27 |
|  |  | S | 0 | (0) | 0 | (0) | 0 | (0) | 0 | (0) | 0 | (0) | 0 | (0) |  |  |  |
|  | Joint pain | Mi | 2 | (4.87) | 0 | (0) | 1 | (2.5) | 0 | (0) | 4 | (5.63) | 7 | (3.05) | 0.15 | 0.33 | 0.86 |
|  | Bruising | Mi | 1 | (2.43) | 1 | (2.5) | 0 | (0) | 1 | (2.7) | 2 | (2.81) | 5 | (2.18) | 0.98 | 0.29 | 0.90 |
|  | Costochondritis | Mo | 1 | (2.43) | 0 | (0) | 0 | (0) | 0 | (0) | 0 | (0) | 1 | (0.43) | 0.32 |  | 0.18 |
|  | Back pain | Mi | 1 | (2.43) | 1 | (2.5) | 2 | (5) | 0 | (0) | 2 | (2.81) | 6 | (2.62) | 0.98 | 0.16 | 0.90 |
|  | Back pain | Mo | 0 | (0) | 0 | (0) | 1 | (2.5) | 0 | (0) | 0 | (0) | 1 | (0.43) |  | 0.33 |  |
|  | Pain in a limb | Mi | 1 | (2.43) | 0 | (0) | 0 | (0) | 0 | (0) | 0 | (0) | 1 | (0.43) | 0.32 |  | 0.18 |
|  | Ligament sprain | Mi | 1 | (2.43) | 1 | (2.5) | 0 | (0) | 0 | (0) | 0 | (0) | 2 | (0.87) | 0.98 |  | 0.18 |
|  | Ligament sprain | Mo | 0 | (0) | 0 | (0) | 1 | (2.5) | 0 | (0) | 0 | (0) | 1 | (0.43) |  | 0.33 |  |
|  | Muscle spasms | Mi | 0 | (0) | 1 | (2.5) | 1 | (2.5) | 1 | (2.7) | 0 | (0) | 3 | (1.31) | 0.30 | 0.95 |  |
|  | Muscle spasms | Mo | 0 | (0) | 0 | (0) | 0 | (0) | 0 | (0) | 1 | (1.4) | 1 | (0.43) |  |  | 0.44 |
|  | Myalgia | Mi | 5 | (12.19) | 3 | (7.5) | 3 | (7.5) | 3 | (8.1) | 6 | (8.45) | 20 | (8.73) | 0.47 | 0.92 | 0.52 |
|  | Myalgia | Mo | 1 | (2.43) | 0 | (0) | 0 | (0) | 0 | (0) | 0 | (0) | 1 | (0.43) | 0.32 |  | 0.18 |
|  | Osteopenia | Mi | 0 | (0) | 0 | (0) | 0 | (0) | 0 | (0) | 1 | (1.4) | 1 | (0.43) |  |  | 0.44 |
|  | Tendonitis | Mi | 0 | (0) | 1 | (2.5) | 0 | (0) | 1 | (2.7) | 1 | (1.4) | 3 | (1.31) | 0.30 | 0.29 | 0.44 |
| Eye disorders |  | Total | 2 | (4.87) | 2 | (5) | 3 | (7.5) | 2 | (5.4) | 1 | (1.4) | 10 | (4.36) | 0.97 | 0.70 | 0.27 |
|  |  | Mi | 2 | (4.87) | 2 | (5) | 2 | (5) | 2 | (5.4) | 1 | (1.4) | 9 | (3.93) | 0.97 | 0.93 | 0.27 |
|  |  | Mo | 0 | (0) | 0 | (0) | 1 | (2.5) | 0 | (0) | 0 | (0) | 1 | (0.43) |  | 0.33 |  |
|  |  | S | 0 | (0) | 0 | (0) | 0 | (0) | 0 | (0) | 0 | (0) | 0 | (0) |  |  |  |
|  | Allergic Conjunctivitis | Mo | 0 | (0) | 0 | (0) | 1 | (2.5) | 0 | (0) | 0 | (0) | 1 | (0.43) |  | 0.33 |  |
|  | Conjunctivitis | Mi | 1 | (2.43) | 0 | (0) | 0 | (0) | 0 | (0) | 0 | (0) | 1 | (0.43) | 0.32 |  | 0.18 |
|  | Eye pain | Mi | 0 | (0) | 0 | (0) | 0 | (0) | 1 | (2.7) | 0 | (0) | 1 | (0.43) |  | 0.29 |  |
|  | Photopsia | Mi | 1 | (2.43) | 0 | (0) | 0 | (0) | 0 | (0) | 0 | (0) | 1 | (0.43) | 0.32 |  | 0.18 |
|  | Sty on the eyelid | Mi | 0 | (0) | 0 | (0) | 0 | (0) | 0 | (0) | 1 | (1.4) | 1 | (0.43) |  |  | 0.44 |
|  | Conjunctival hemorrhage | Mi | 0 | (0) | 0 | (0) | 1 | (2.5) | 0 | (0) | 0 | (0) | 1 | (0.43) |  | 0.33 |  |
|  | Iris hemorrhage | Mi | 0 | (0) | 0 | (0) | 1 | (2.5) | 0 | (0) | 0 | (0) | 1 | (0.43) |  | 0.33 |  |
|  | Eye discomfort | Mi | 0 | (0) | 1 | (2.5) | 0 | (0) | 0 | (0) | 0 | (0) | 1 | (0.43) | 0.30 |  |  |
|  | Abnormal sensation in the eye | Mi | 0 | (0) | 1 | (2.5) | 0 | (0) | 1 | (2.7) | 0 | (0) | 2 | (0.87) | 0.30 | 0.29 |  |
| Psychiatric Disorders |  | Total | 1 | (2.43) | 1 | (2.5) | 4 | (10) | 0 | (0) | 4 | (5.63) | 10 | (4.36) | 0.98 | 0.04 | 0.43 |
|  |  | Mi | 1 | (2.43) | 1 | (2.5) | 3 | (7.5) | 0 | (0) | 3 | (4.22) | 8 | (3.49) | 0.98 | 0.08 | 0.62 |
|  |  | Mo | 0 | (0) | 0 | (0) | 2 | (5) | 0 | (0) | 2 | (2.81) | 4 | (1.74) |  | 0.16 | 0.27 |
|  |  | S | 0 | (0) | 0 | (0) | 0 | (0) | 0 | (0) | 0 | (0) | 0 | (0) |  |  |  |
|  | Agitation | Mi | 0 | (0) | 0 | (0) | 0 | (0) | 0 | (0) | 1 | (1.4) | 1 | (0.43) |  |  | 0.44 |
|  | Attention Impairment | Mi | 0 | (0) | 1 | (2.5) | 0 | (0) | 0 | (0) | 1 | (1.4) | 2 | (0.87) | 0.30 |  | 0.44 |
|  | Anxiety | Mi | 1 | (2.43) | 0 | (0) | 1 | (2.5) | 0 | (0) | 1 | (1.4) | 3 | (1.31) | 0.32 | 0.33 | 0.69 |
|  | Depression | Mi | 0 | (0) | 0 | (0) | 1 | (2.5) | 0 | (0) | 0 | (0) | 1 | (0.43) |  | 0.33 |  |
|  | Stress | Mo | 0 | (0) | 0 | (0) | 0 | (0) | 0 | (0) | 1 | (1.4) | 1 | (0.43) |  |  | 0.44 |
|  | Insomnia | Mo | 0 | (0) | 0 | (0) | 1 | (2.5) | 0 | (0) | 0 | (0) | 1 | (0.43) |  | 0.33 |  |
|  | Panic Reaction | Mo | 0 | (0) | 0 | (0) | 0 | (0) | 0 | (0) | 1 | (1.4) | 1 | (0.43) |  |  | 0.44 |
|  | Mixed Anxiety and Depressive Disorder | Mo | 0 | (0) | 0 | (0) | 1 | (2.5) | 0 | (0) | 0 | (0) | 1 | (0.43) |  | 0.33 |  |
|  | Sleep Disorder | Mi | 0 | (0) | 0 | (0) | 1 | (2.5) | 0 | (0) | 0 | (0) | 1 | (0.43) |  | 0.33 |  |
| Renal and Urinary Disorders |  | Total | 1 | (2.43) | 0 | (0) | 0 | (0) | 0 | (0) | 1 | (1.4) | 2 | (0.87) | 0.32 |  | 0.69 |
|  |  | Mi | 0 | (0) | 0 | (0) | 0 | (0) | 0 | (0) | 1 | (1.4) | 1 | (0.43) |  |  | 0.44 |
|  |  | Mo | 0 | (0) | 0 | (0) | 0 | (0) | 0 | (0) | 0 | (0) | 0 | (0) |  |  |  |
|  |  | S | 1 | (2.43) | 0 | (0) | 0 | (0) | 0 | (0) | 0 | (0) | 1 | (0.43) | 0.32 |  | 0.18 |
|  | Renal Colic | S | 1 | (2.43) | 0 | (0) | 0 | (0) | 0 | (0) | 0 | (0) | 1 | (0.43) | 0.32 |  | 0.18 |
|  | Dysuria | Mi | 0 | (0) | 0 | (0) | 0 | (0) | 0 | (0) | 1 | (1.4) | 1 | (0.43) |  |  | 0.44 |
| Respiratory, thoracic, and mediastinal disorders |  | Total | 12 | (29.26) | 10 | (25) | 15 | (37.5) | 9 | (24.32) | 11 | (15.49) | 57 | (24.89) | 0.66 | 0.21 | 0.08 |
|  |  | Mi | 12 | (29.26) | 10 | (25) | 15 | (37.5) | 9 | (24.32) | 10 | (14.08) | 56 | (24.45) | 0.66 | 0.21 | 0.05 |
|  |  | Mo | 1 | (2.43) | 0 | (0) | 2 | (5) | 0 | (0) | 0 | (0) | 3 | (1.31) | 0.32 | 0.16 | 0.18 |
|  |  | S | 1 | (2.43) | 0 | (0) | 0 | (0) | 0 | (0) | 1 | (1.4) | 2 | (0.87) | 0.32 |  | 0.69 |
|  | Dyspnea | S | 0 | (0) | 0 | (0) | 0 | (0) | 0 | (0) | 1 | (1.4) | 1 | (0.43) |  |  | 0.44 |
|  | Pharyngolaryngeal pain | Mi | 0 | (0) | 0 | (0) | 0 | (0) | 2 | (5.4) | 0 | (0) | 2 | (0.87) |  | 0.13 |  |
|  | Pharyngolaryngeal pain | Mi | 0 | (0) | 1 | (2.5) | 0 | (0) | 0 | (0) | 0 | (0) | 1 | (0.43) | 0.30 |  |  |
|  | Pharyngeal erythema | Mi | 1 | (2.43) | 5 | (12.5) | 2 | (5) | 1 | (2.7) | 0 | (0) | 9 | (3.93) | 0.08 | 0.60 | 0.18 |
|  | Epistaxis | S | 1 | (2.43) | 0 | (0) | 0 | (0) | 0 | (0) | 0 | (0) | 1 | (0.43) | 0.32 |  | 0.18 |
|  | Pharyngotonsillitis | Mi | 0 | (0) | 0 | (0) | 0 | (0) | 3 | (8.1) | 2 | (2.81) | 5 | (2.18) |  | 0.06 | 0.27 |
|  | Pharyngotonsillitis | Mi | 1 | (2.43) | 0 | (0) | 3 | (7.5) | 1 | (2.7) | 0 | (0) | 5 | (2.18) | 0.32 | 0.34 | 0.18 |

| SOC<br>PT | SEV | IM<br>AVX | % | IM PL | % | IN<br>AVX | % | IN PL | % | IM AZ | % | Total | % | IM<br>AVX-PL<br>p-value | IN<br>AVX-PL<br>p-value | AVX-AZ<br>p-value |
| --- | --- | --- | --- | --- | --- | --- | --- | --- | --- | --- | --- | --- | --- | --- | --- | --- |
| Hypertrophy of nasal turbinates | Mi | 2 | (4.87) | 0 | (0) | 3 | (7.5) | 2 | (5.4) | 1 | (1.4) | 8 | (3.49) | 0.15 | 0.70 | 0.27 |
| Throat irritation | Mo | 1 | (2.43) | 0 | (0) | 1 | (2.5) | 0 | (0) | 0 | (0) | 2 | (0.87) | 0.32 | 0.33 | 0.18 |
| Nasal discomfort | Mi | 0 | (0) | 0 | (0) | 1 | (2.5) | 0 | (0) | 0 | (0) | 1 | (0.43) |  | 0.33 |  |
| Nasopharyngitis | Mi | 0 | (0) | 0 | (0) | 0 | (0) | 2 | (5.4) | 0 | (0) | 2 | (0.87) |  | 0.13 |  |
| Allergic rhinitis | Mi | 0 | (0) | 0 | (0) | 2 | (5) | 0 | (0) | 0 | (0) | 2 | (0.87) |  | 0.16 |  |
| Rhinorrhea | Mi | 1 | (2.43) | 0 | (0) | 0 | (0) | 0 | (0) | 0 | (0) | 1 | (0.43) | 0.32 |  | 0.18 |
| Nasal dryness | Mi | 0 | (0) | 0 | (0) | 1 | (2.5) | 0 | (0) | 0 | (0) | 1 | (0.43) |  | 0.33 |  |
| Cough | Mi | 6 | (14.63) | 4 | (10) | 6 | (15) | 5 | (13.51) | 3 | (4.22) | 24 | (10.48) | 0.52 | 0.85 | 0.05 |
| Cough | Mi | 0 | (0) | 0 | (0) | 2 | (5) | 0 | (0) | 0 | (0) | 2 | (0.87) |  | 0.16 |  |
| Laryngeal disorder | Mi | 4 | (9.75) | 1 | (2.5) | 2 | (5) | 3 | (8.1) | 5 | (7.04) | 15 | (6.55) | 0.17 | 0.58 | 0.61 |
| Dyspnea | Mo | 0 | (0) | 0 | (0) | 1 | (2.5) | 0 | (0) | 0 | (0) | 1 | (0.43) |  | 0.33 |  |
| Pharyngolaryngeal pain | Mi | 0 | (0) | 1 | (2.5) | 0 | (0) | 0 | (0) | 0 | (0) | 1 | (0.43) | 0.30 |  |  |
| Vascular Disorders | Total | 3 | (7.31) | 0 | (0) | 4 | (10) | 1 | (2.7) | 3 | (4.22) | 11 | (4.8) | 0.08 | 0.19 | 0.48 |
|  | Mi | 3 | (7.31) | 0 | (0) | 4 | (10) | 1 | (2.7) | 3 | (4.22) | 11 | (4.8) | 0.08 | 0.19 | 0.48 |
|  | Mo | 0 | (0) | 0 | (0) | 0 | (0) | 0 | (0) | 0 | (0) | 0 | (0) |  |  |  |
|  | S | 0 | (0) | 0 | (0) | 0 | (0) | 0 | (0) | 0 | (0) | 0 | (0) |  |  |  |
| Hypertensive crisis | Mi | 0 | (0) | 0 | (0) | 1 | (2.5) | 0 | (0) | 0 | (0) | 1 | (0.43) |  | 0.33 |  |
| Peripheral Vascular Disease | Mi | 0 | (0) | 0 | (0) | 0 | (0) | 0 | (0) | 1 | (1.4) | 1 | (0.43) |  |  | 0.44 |
| Hematoma | Mi | 1 | (2.43) | 0 | (0) | 0 | (0) | 0 | (0) | 1 | (1.4) | 2 | (0.87) | 0.32 |  | 0.69 |
| Hemorrhoids | Mi | 0 | (0) | 0 | (0) | 0 | (0) | 0 | (0) | 1 | (1.4) | 1 | (0.43) |  |  | 0.44 |
| Hypertension | Mi | 1 | (2.43) | 0 | (0) | 3 | (7.5) | 1 | (2.7) | 0 | (0) | 5 | (2.18) | 0.32 | 0.34 | 0.18 |
| Hypotension | Mi | 1 | (2.43) | 0 | (0) | 1 | (2.5) | 0 | (0) | 0 | (0) | 2 | (0.87) | 0.32 | 0.33 | 0.18 |

SOC: System Organ Class; PT: Preferred Term; AE: adverse event; SEV: severity; Mi: mild; Mo: moderate; S: severe; AVX: AVX/COVID-12; AZ: AZ/ChAdOx-1-S; IM: intramuscular; IN: intranasal; PL: placebo. p-value calculated from the comparison of proportions (Z-test).

**Table 2S.**
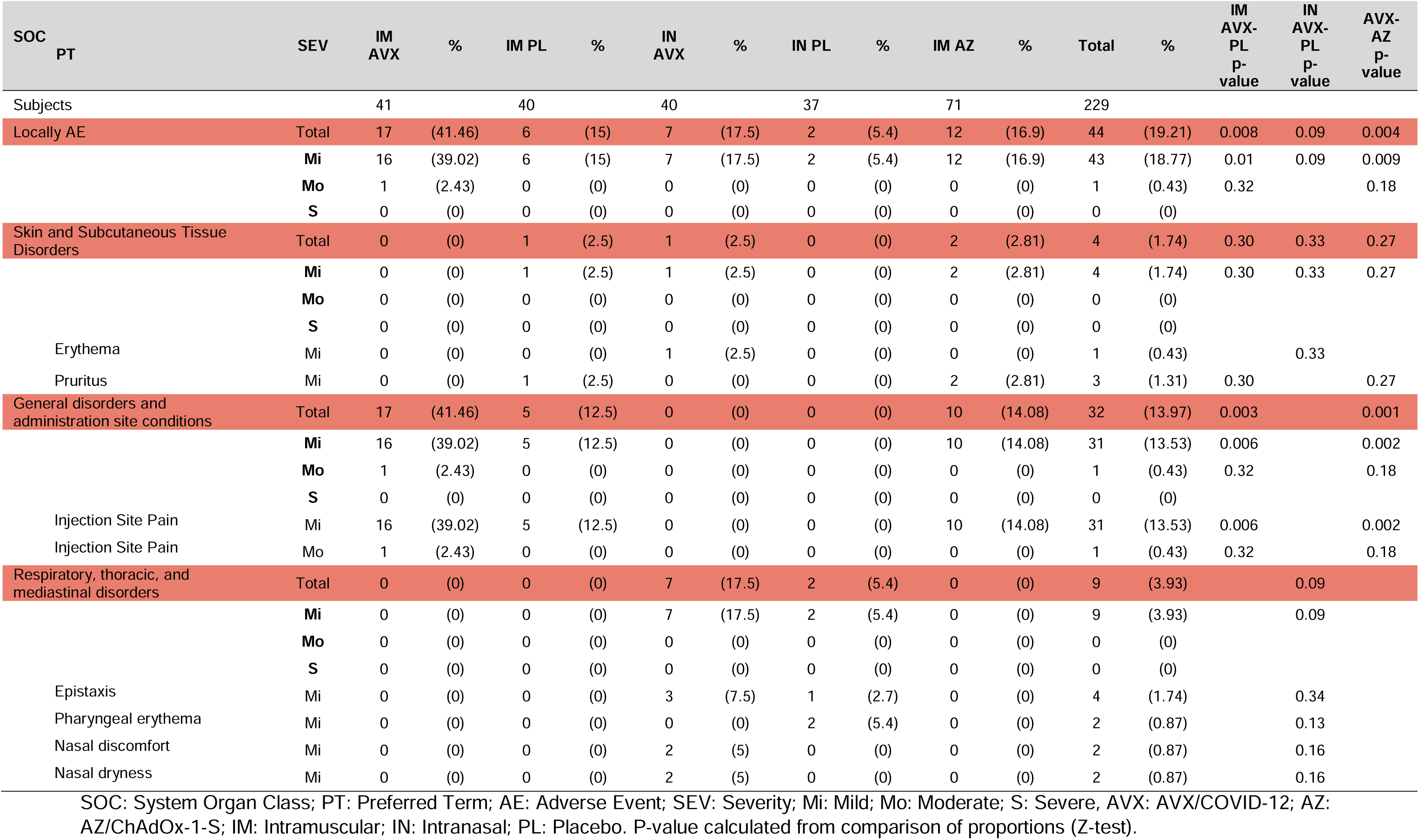
Proportion of affected subjects by local adverse events of special interest at 7 days post-immunization.

| SOC<br>PT | SEV | IM<br>AVX | % | IM PL | % | IN<br>AVX | % | IN PL | % | IM AZ | % | Total | % | IM<br>AVX-PL<br>p-value | IN<br>AVX-PL<br>p-value | AVX-AZ<br>p-value |
| --- | --- | --- | --- | --- | --- | --- | --- | --- | --- | --- | --- | --- | --- | --- | --- | --- |
| Subjects |  | 41 |  | 40 |  | 40 |  | 37 |  | 71 |  | 229 |  |  |  |  |
| Locally AE | Total | 17 | (41.46) | 6 | (15) | 7 | (17.5) | 2 | (5.4) | 12 | (16.9) | 44 | (19.21) | 0.008 | 0.09 | 0.004 |
|  | Mi | 16 | (39.02) | 6 | (15) | 7 | (17.5) | 2 | (5.4) | 12 | (16.9) | 43 | (18.77) | 0.01 | 0.09 | 0.009 |
|  | Mo | 1 | (2.43) | 0 | (0) | 0 | (0) | 0 | (0) | 0 | (0) | 1 | (0.43) | 0.32 |  | 0.18 |
|  | S | 0 | (0) | 0 | (0) | 0 | (0) | 0 | (0) | 0 | (0) | 0 | (0) |  |  |  |
| Skin and Subcutaneous Tissue Disorders | Total | 0 | (0) | 1 | (2.5) | 1 | (2.5) | 0 | (0) | 2 | (2.81) | 4 | (1.74) | 0.30 | 0.33 | 0.27 |
|  | Mi | 0 | (0) | 1 | (2.5) | 1 | (2.5) | 0 | (0) | 2 | (2.81) | 4 | (1.74) | 0.30 | 0.33 | 0.27 |
|  | Mo | 0 | (0) | 0 | (0) | 0 | (0) | 0 | (0) | 0 | (0) | 0 | (0) |  |  |  |
|  | S | 0 | (0) | 0 | (0) | 0 | (0) | 0 | (0) | 0 | (0) | 0 | (0) |  |  |  |
| Erythema | Mi | 0 | (0) | 0 | (0) | 1 | (2.5) | 0 | (0) | 0 | (0) | 1 | (0.43) |  | 0.33 |  |
| Pruritus | Mi | 0 | (0) | 1 | (2.5) | 0 | (0) | 0 | (0) | 2 | (2.81) | 3 | (1.31) | 0.30 |  | 0.27 |
| General disorders and administration site conditions | Total | 17 | (41.46) | 5 | (12.5) | 0 | (0) | 0 | (0) | 10 | (14.08) | 32 | (13.97) | 0.003 |  | 0.001 |
|  | Mi | 16 | (39.02) | 5 | (12.5) | 0 | (0) | 0 | (0) | 10 | (14.08) | 31 | (13.53) | 0.006 |  | 0.002 |
|  | Mo | 1 | (2.43) | 0 | (0) | 0 | (0) | 0 | (0) | 0 | (0) | 1 | (0.43) | 0.32 |  | 0.18 |
|  | S | 0 | (0) | 0 | (0) | 0 | (0) | 0 | (0) | 0 | (0) | 0 | (0) |  |  |  |
| Injection Site Pain | Mi | 16 | (39.02) | 5 | (12.5) | 0 | (0) | 0 | (0) | 10 | (14.08) | 31 | (13.53) | 0.006 |  | 0.002 |
| Injection Site Pain | Mo | 1 | (2.43) | 0 | (0) | 0 | (0) | 0 | (0) | 0 | (0) | 1 | (0.43) | 0.32 |  | 0.18 |
| Respiratory, thoracic, and mediastinal disorders | Total | 0 | (0) | 0 | (0) | 7 | (17.5) | 2 | (5.4) | 0 | (0) | 9 | (3.93) |  | 0.09 |  |
|  | Mi | 0 | (0) | 0 | (0) | 7 | (17.5) | 2 | (5.4) | 0 | (0) | 9 | (3.93) |  | 0.09 |  |
|  | Mo | 0 | (0) | 0 | (0) | 0 | (0) | 0 | (0) | 0 | (0) | 0 | (0) |  |  |  |
|  | S | 0 | (0) | 0 | (0) | 0 | (0) | 0 | (0) | 0 | (0) | 0 | (0) |  |  |  |
| Epistaxis | Mi | 0 | (0) | 0 | (0) | 3 | (7.5) | 1 | (2.7) | 0 | (0) | 4 | (1.74) |  | 0.34 |  |
| Pharyngeal erythema | Mi | 0 | (0) | 0 | (0) | 0 | (0) | 2 | (5.4) | 0 | (0) | 2 | (0.87) |  | 0.13 |  |
| Nasal discomfort | Mi | 0 | (0) | 0 | (0) | 2 | (5) | 0 | (0) | 0 | (0) | 2 | (0.87) |  | 0.16 |  |
| Nasal dryness | Mi | 0 | (0) | 0 | (0) | 2 | (5) | 0 | (0) | 0 | (0) | 2 | (0.87) |  | 0.16 |  |

SOC: System Organ Class; PT: Preferred Term; AE: Adverse Event; SEV: Severity; Mi: Mild; Mo: Moderate; S: Severe, AVX: AVX/COVID-12; AZ: AZ/ChAdOx-1-S; IM: Intramuscular; IN: Intranasal; PL: Placebo. P-value calculated from comparison of proportions (Z-test).

**Table 3S.**
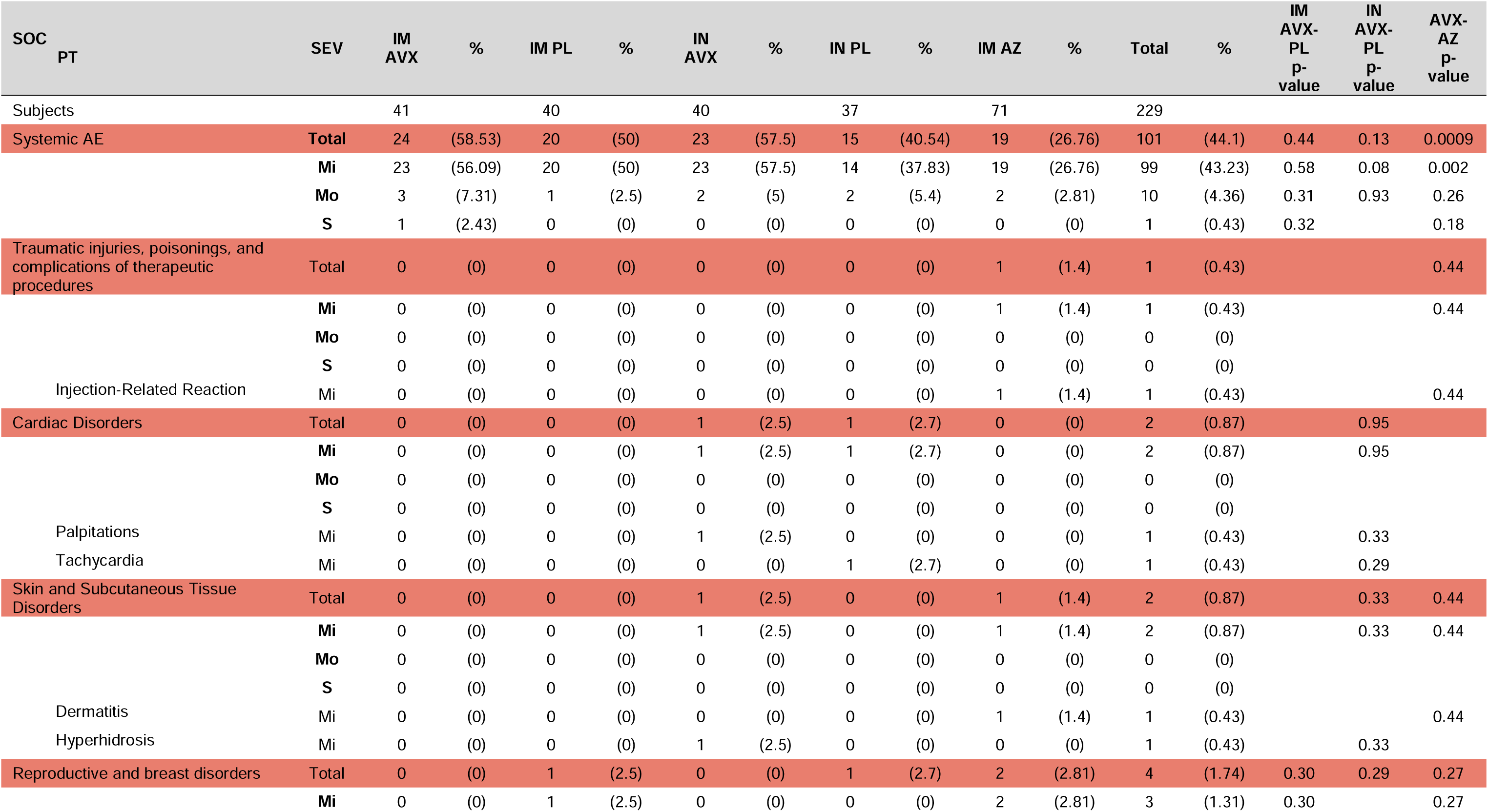
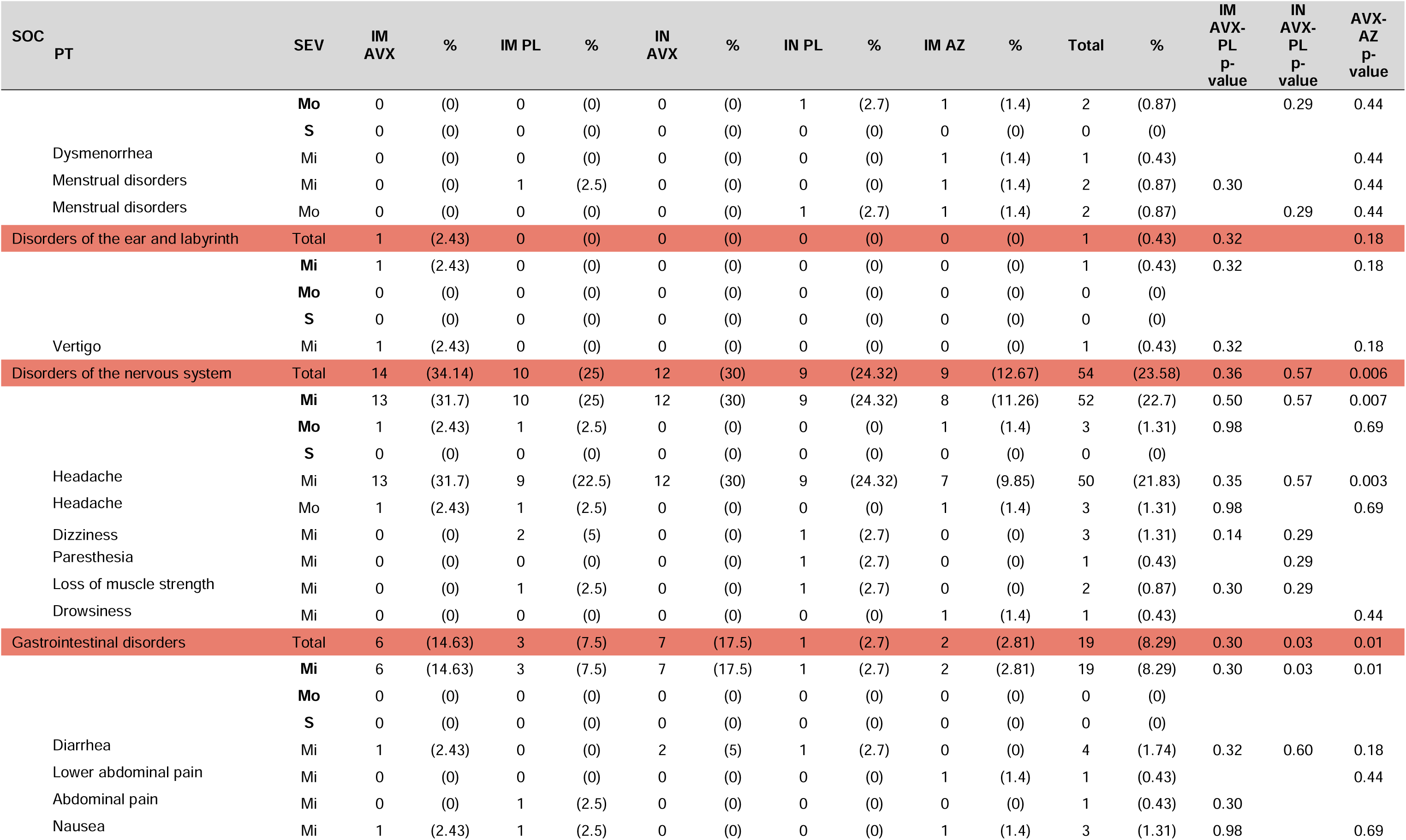
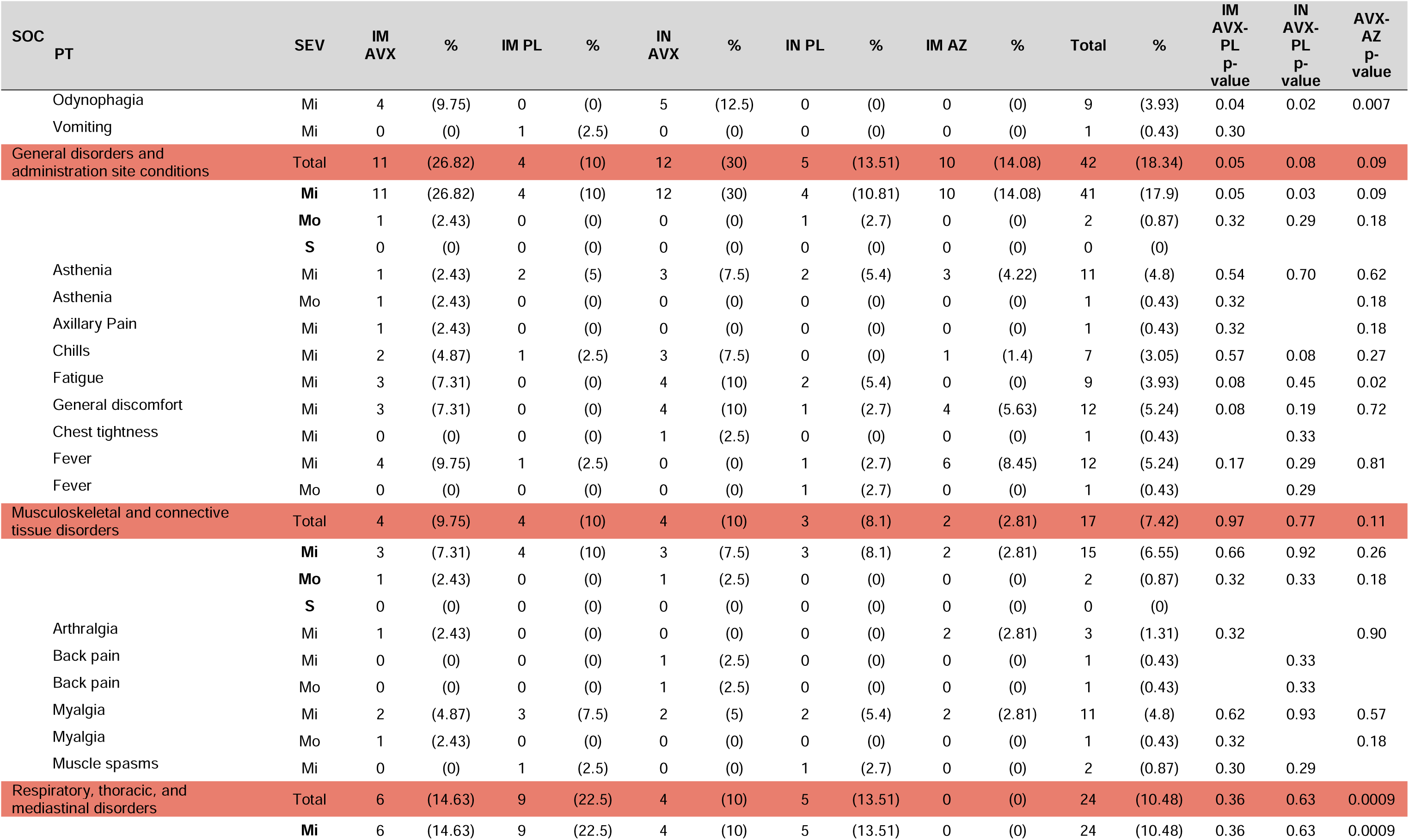
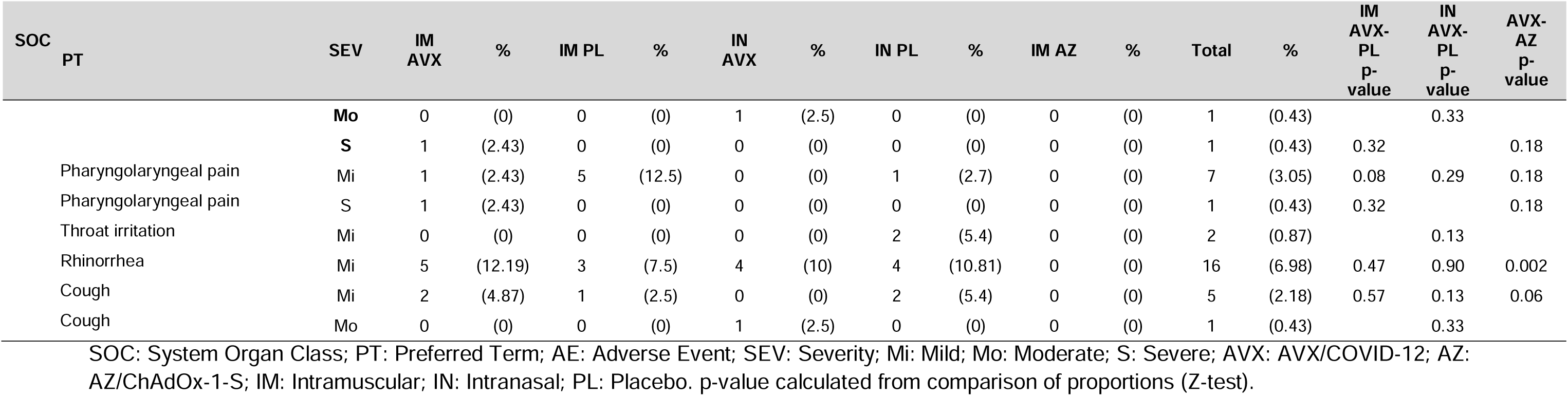
Proportion of affected subjects by systemic adverse events of special interest at 7 days post-immunization.

**Table 4S.**
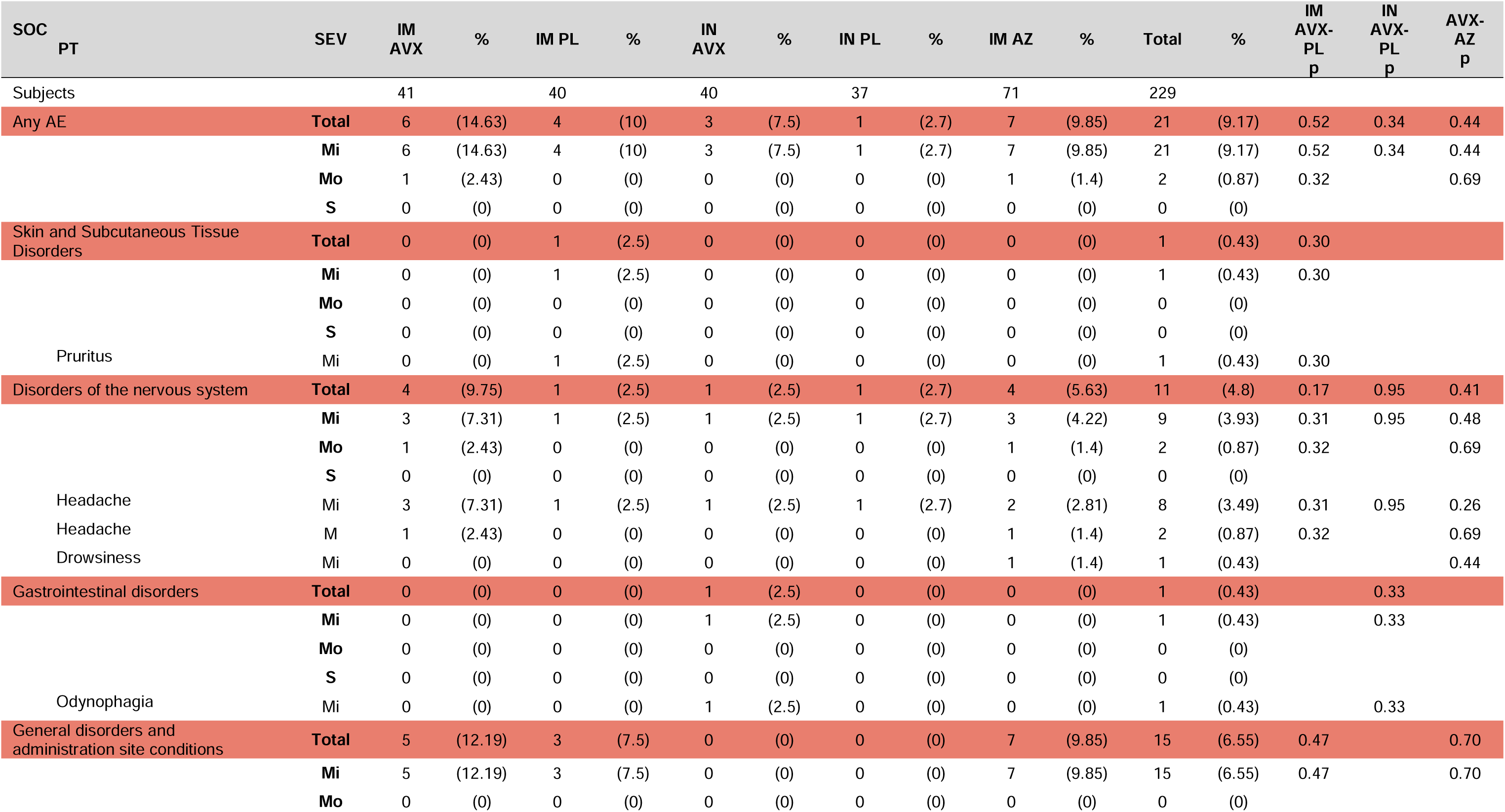
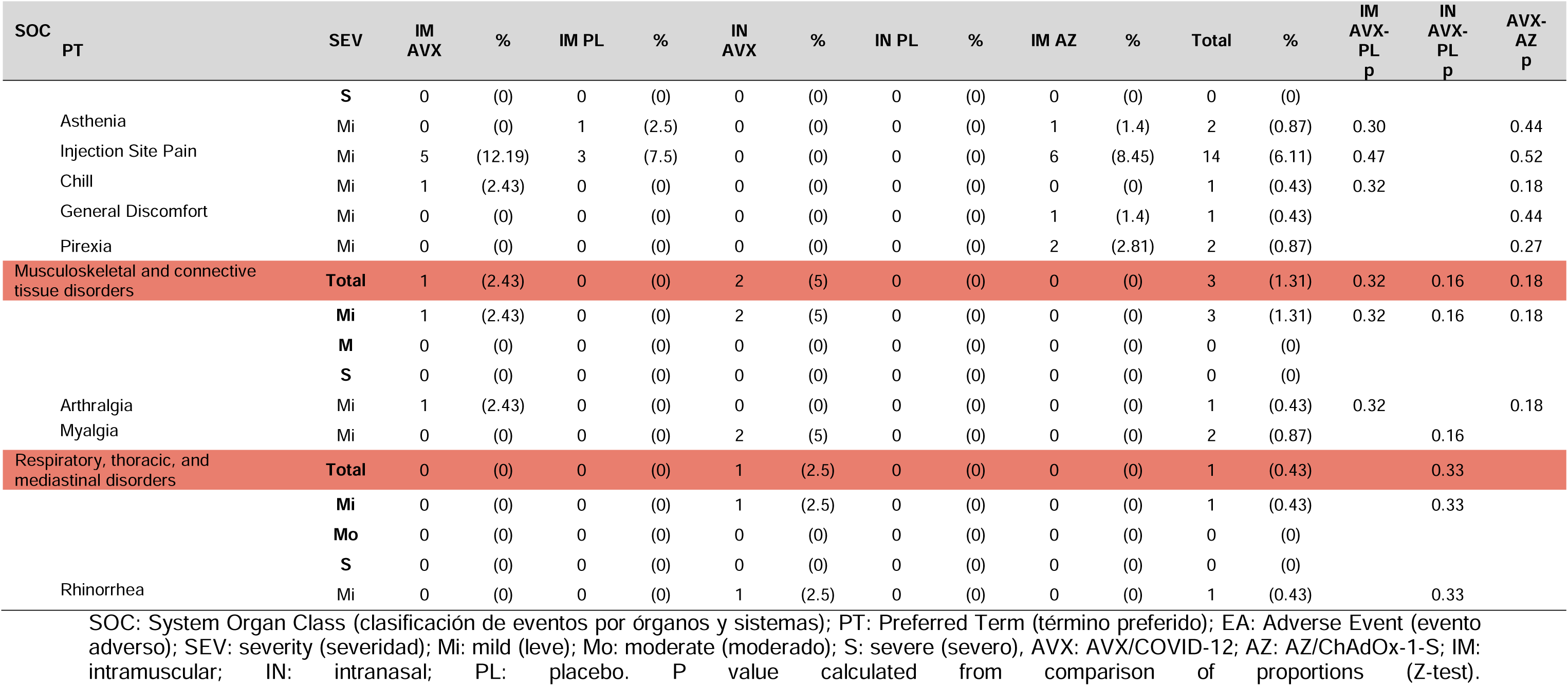
Proportion of subjects affected by adverse events associated with vaccination at 7 days post-immunization.

**Table 5S.** Antibody titers throughout the study in subjects vaccinated with AVX/COVID-12 via intramuscular route.

| IM AVX/COVID-12 |  |  |  |  |  |  |
| --- | --- | --- | --- | --- | --- | --- |
|  | Baseline | Day 14 | Day 42 | Day 90 | Day 180 | Day 365 |
| **Volunteers** | 41 | 38 | 38 | 39 | 39 | 36 |
| **Antibody titers IgG against spike protein** |  |  |  |  |  |  |
| GM | 227.06 | 2171.4 | 1489 | 1399.7 | 1013.4 | 1514.2 |
| CI | 166 - 310.5 | 1609.2 - 2930 | 1083.8 - 2045.9 | 977.4 - 2004.4 | 651.4 - 1576.4 | 1054.2 - 2174.8 |
| p-Value |  | <0.001 | <0.001 | <0.001 | <0.001 | <0.001 |
| **Wuhan-1 neutralizing titers** |  |  |  |  |  |  |
| GM | 37.2 | 308.7 | 509.7 | 372.5 | 382.8 | 513.9 |
| CI | 28.8 - 48.2 | 205.5 - 463.6 | 347.4 - 747.8 | 226.2 - 613.6 | 217.2 - 674.7 | 332 - 795.6 |
| p-Value |  | <0.001 | <0.001 | <0.001 | <0.001 | <0.001 |
| **Delta neutralizing titers** |  |  |  |  |  |  |
| GM | 20.2 | 68.3 | 87.4 | 118.2 | 150.1 | 211.5 |
| CI | 17.8 - 23.1 | 45.9 - 101.6 | 55.3 - 138.2 | 71.2 - 196.1 | 91.5 - 246.1 | 132.3 - 337.9 |
| p-Value |  | <0.001 | <0.001 | <0.001 | <0.001 | <0.001 |
| **Alpha neutralizing titers** |  |  |  |  |  |  |
| GM | 39.5 | 292.68 | 742.6 | 452.2 | 437.3 | 372.7 |
| CI | 30 - 52 | 209.1 - 409.7 | 443.3 - 1244 | 281.3 - 726.8 | 255.7 - 747.7 | 234.5 - 592.5 |
| p-Value |  | <0.001 | <0.001 | <0.001 | <0.001 | <0.001 |
| **Beta neutralizing titers** |  |  |  |  |  |  |
| GM | 24.8 | 62.4 | 89.1 | 103.4 | 110.4 | 244.3 |
| CI | 20.8 - 29.7 | 42.5 - 91.7 | 55 - 144.2 | 62.7 - 170.5 | 67.1 - 181.9 | 151 - 395.4 |
| p-Value |  | 0.002 | <0.001 | <0.001 | <0.001 | <0.001 |
| **BA.2 neutralizing titers\*** |  |  |  |  |  |  |
| GM | 23.6 | 88.2 | ND | ND | ND | ND |
| CI | 20.3 - 27.5 | 55.6 - 139.7 |  |  |  |  |
| p-Value |  | <0.001 |  |  |  |  |
| **BA.5 neutralizing titers\*** |  |  |  |  |  |  |
| GM | 23 | 143 | ND | ND | ND | ND |
| CI | 19.7 - 26.9 | 94.5 - 216.3 |  |  |  |  |
| p-Value |  | <0.001 |  |  |  |  |

\*Sample size n=39, AVX: AVX/COVID-12; IM: intramuscular; ND: not determined; GM: geometric mean; CI: confidence interval. Comparison of GM tested with Wilcoxon signed-rank test (paired), variable on logarithmic scale.

**Table 6S.** Antibody titers throughout the study in subjects vaccinated with AVX/COVID-12 intramuscularly who did not experience COVID-19 during the study.

| AVX/COVID-12 IM |  |  |  |  |  |
| --- | --- | --- | --- | --- | --- |
|  | Day 14 | Day 42 | Day 90 | Day 180 | Day 365 |
| **Sujetos** | 34 | 28 | 25 | 15 | 12 |
| **Antibody titers IgG against spike protein** |  |  |  |  |  |
| GM | 2056.2 | 1226.6 | 1022 | 388.6 | 1133.9 |
| CI | 1496.4 - 2825.6 | 844.3 - 1782.1 | 651.5 - 1603 | 197.8 - 763.4 | 463 - 2776.8 |
| **Wuhan-1 neutralizing titers** |  |  |  |  |  |
| GM | 289 | 404.3 | 286.3 | 142.4 | 466.9 |
| CI | 185.9 - 449.1 | 258.1 - 633.4 | 158.2 - 518 | 63.9 - 317.6 | 164.7 - 1323.4 |
| **Delta neutralizing titers** |  |  |  |  |  |
| GM | 60.6 | 57.8 | 70.4 | 57.6 | 201.8 |
| CI | 39.7 - 92.3 | 35.9 - 93.1 | 40.1 - 123.7 | 34.3 - 96.8 | 70.3 - 579.1 |
| **Alpha neutralizing titers** |  |  |  |  |  |
| GM | 286.4 | 603.1 | 346.9 | 192.1 | 338.7 |
| CI | 197.2 - 416.1 | 328.2 - 1107.8 | 188.9 - 636.9 | 79.1 - 466.4 | 108.7 - 1055.4 |
| **Beta neutralizing titers** |  |  |  |  |  |
| GM | 55.3 | 70.5 | 58.9 | 44.7 | 226.9 |
| CI | 37.3 - 82.1 | 40.4 - 122.8 | 33.3 - 104 | 25 - 80 | 71.3 - 721.3 |
| **BA.2 neutralizing titers** |  |  |  |  |  |
| GM | 76.6 | ND | ND | ND | ND |
| CI | 47.3 - 124 |  |  |  |  |
| **BA.5 neutralizing titers** |  |  |  |  |  |
| GM | 130.1 | ND | ND | ND | ND |
| CI | 83.8 - 202.2 |  |  |  |  |

AVX: AVX/COVID-12; IM: intramuscular; ND: not determined; GM: geometric mean; CI: confidence interval. Comparison of GM using Wilcoxon signed-rank test (paired), variable on a logarithmic scale.

**Table 7S.**
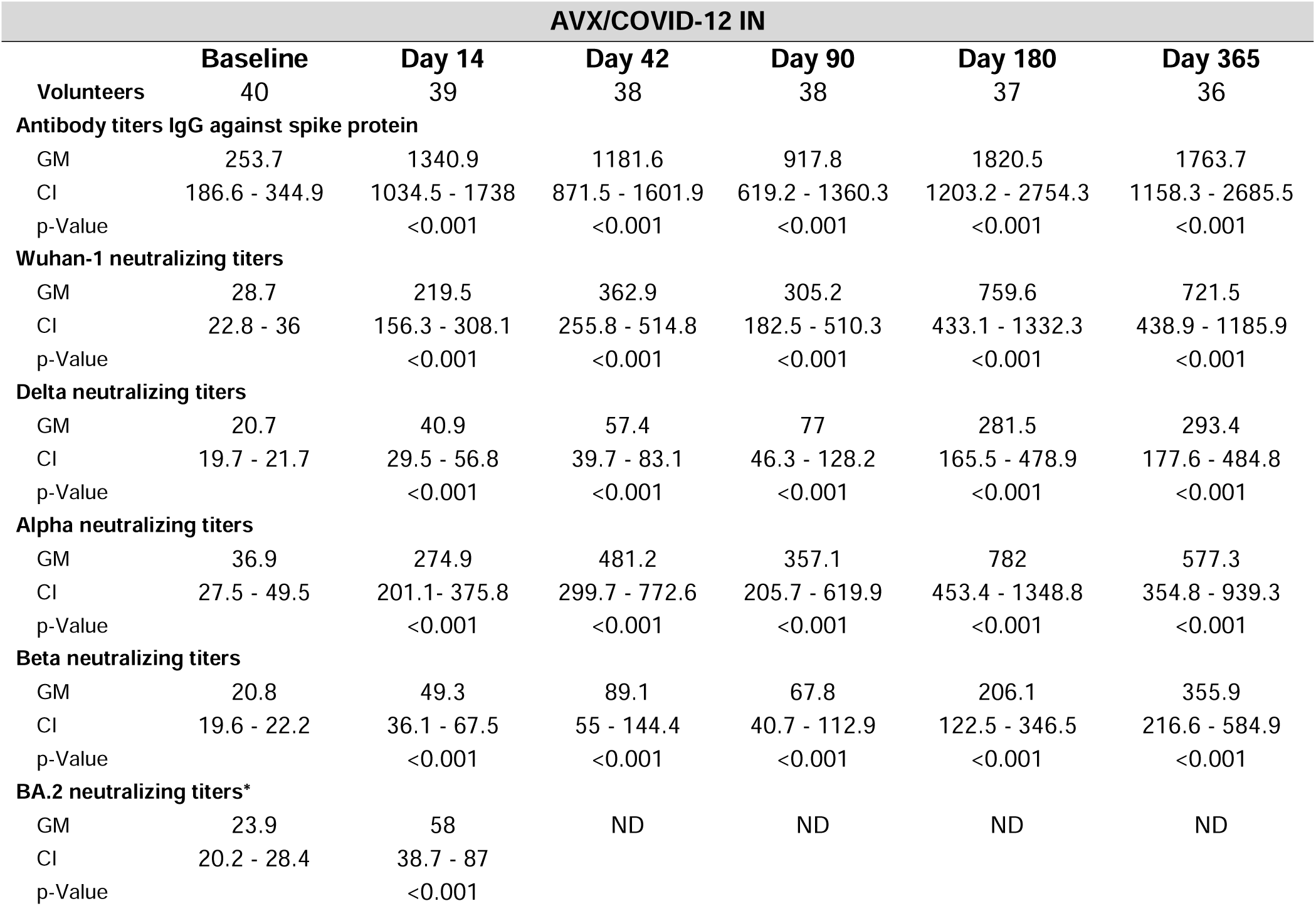
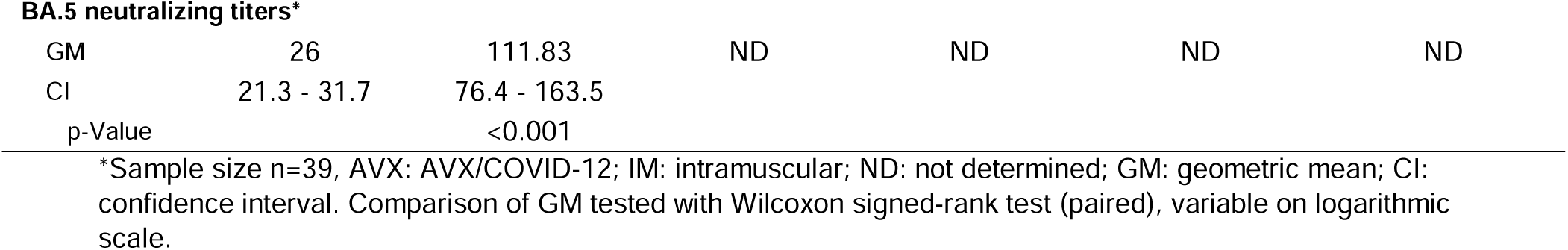
Antibody titers over the study duration in subjects vaccinated with AVX/COVID-12 via the intranasal route.

| AVX/COVID-12 IN |  |  |  |  |  |  |
| --- | --- | --- | --- | --- | --- | --- |
|  | Baseline | Day 14 | Day 42 | Day 90 | Day 180 | Day 365 |
| Volunteers | 40 | 39 | 38 | 38 | 37 | 36 |
| **Antibody titers IgG against spike protein** |  |  |  |  |  |  |
| GM | 253.7 | 1340.9 | 1181.6 | 917.8 | 1820.5 | 1763.7 |
| CI | 186.6 - 344.9 | 1034.5 - 1738 | 871.5 - 1601.9 | 619.2 - 1360.3 | 1203.2 - 2754.3 | 1158.3 - 2685.5 |
| p-Value |  | <0.001 | <0.001 | <0.001 | <0.001 | <0.001 |
| **Wuhan-1 neutralizing titers** |  |  |  |  |  |  |
| GM | 28.7 | 219.5 | 362.9 | 305.2 | 759.6 | 721.5 |
| CI | 22.8 - 36 | 156.3 - 308.1 | 255.8 - 514.8 | 182.5 - 510.3 | 433.1 - 1332.3 | 438.9 - 1185.9 |
| p-Value |  | <0.001 | <0.001 | <0.001 | <0.001 | <0.001 |
| **Delta neutralizing titers** |  |  |  |  |  |  |
| GM | 20.7 | 40.9 | 57.4 | 77 | 281.5 | 293.4 |
| CI | 19.7 - 21.7 | 29.5 - 56.8 | 39.7 - 83.1 | 46.3 - 128.2 | 165.5 - 478.9 | 177.6 - 484.8 |
| p-Value |  | <0.001 | <0.001 | <0.001 | <0.001 | <0.001 |
| **Alpha neutralizing titers** |  |  |  |  |  |  |
| GM | 36.9 | 274.9 | 481.2 | 357.1 | 782 | 577.3 |
| CI | 27.5 - 49.5 | 201.1 - 375.8 | 299.7 - 772.6 | 205.7 - 619.9 | 453.4 - 1348.8 | 354.8 - 939.3 |
| p-Value |  | <0.001 | <0.001 | <0.001 | <0.001 | <0.001 |
| **Beta neutralizing titers** |  |  |  |  |  |  |
| GM | 20.8 | 49.3 | 89.1 | 67.8 | 206.1 | 355.9 |
| CI | 19.6 - 22.2 | 36.1 - 67.5 | 55 - 144.4 | 40.7 - 112.9 | 122.5 - 346.5 | 216.6 - 584.9 |
| p-Value |  | <0.001 | <0.001 | <0.001 | <0.001 | <0.001 |
| **BA.2 neutralizing titers\*** |  |  |  |  |  |  |
| GM | 23.9 | 58 | ND | ND | ND | ND |
| CI | 20.2 - 28.4 | 38.7 - 87 |  |  |  |  |
| p-Value |  | <0.001 |  |  |  |  |
| **BA.5 neutralizing titers\*** |  |  |  |  |  |  |
| GM | 26 | 111.83 | ND | ND | ND | ND |
| CI | 21.3 - 31.7 | 76.4 - 163.5 |  |  |  |  |
| p-Value |  | <0.001 |  |  |  |  |

\*Sample size n=39, AVX: AVX/COVID-12; IM: intramuscular; ND: not determined; GM: geometric mean; CI: confidence interval. Comparison of GM tested with Wilcoxon signed-rank test (paired), variable on logarithmic scale.

**Table 8S.**
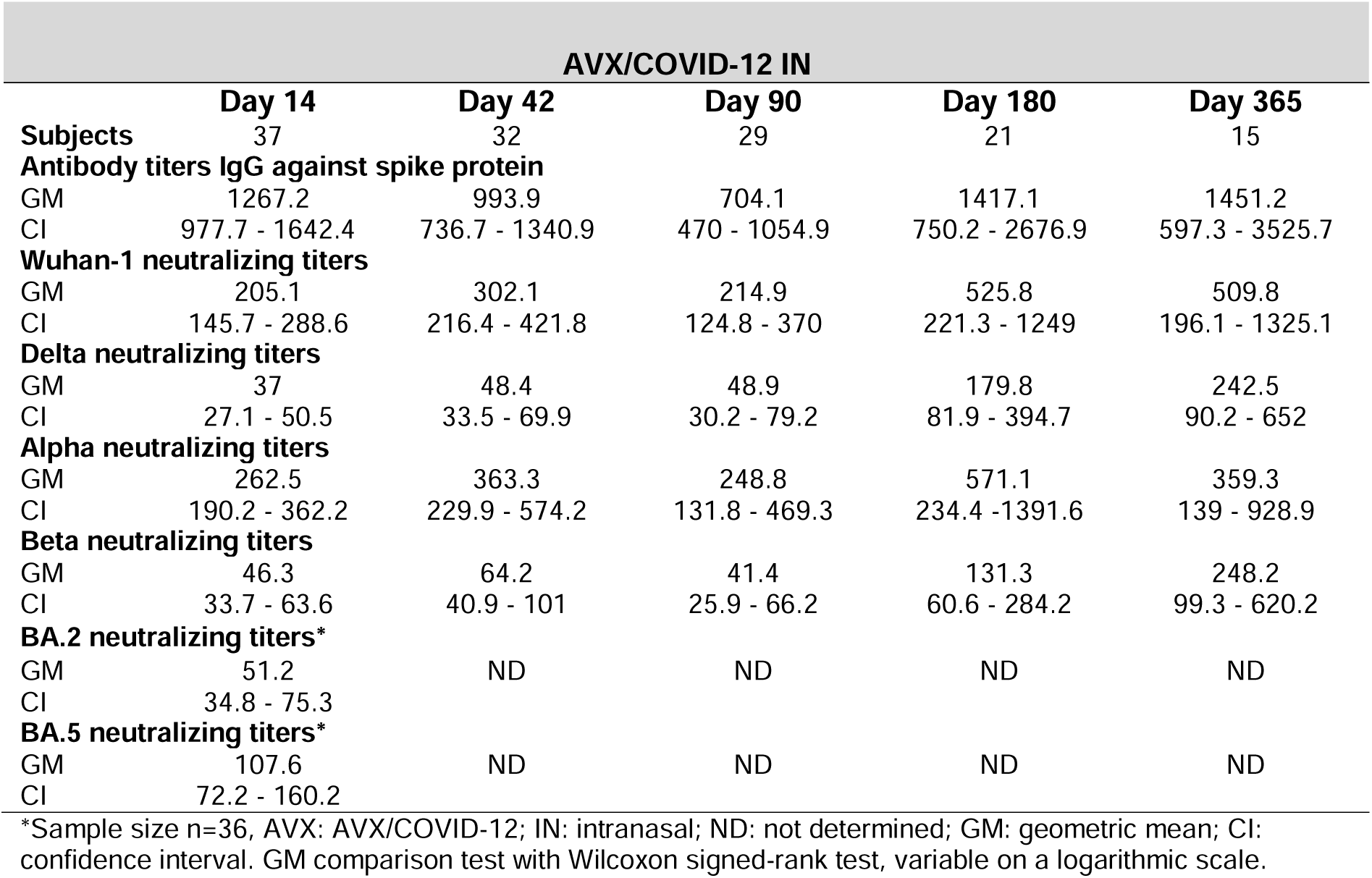
Antibody titers throughout the study in subjects vaccinated with AVX/COVID-12 intranasally who did not experience COVID-19 during the study.

